# PANDORA: Population Archive of Neuroimaging Data Organized for Rapid Analysis

**DOI:** 10.64898/2026.01.05.26343425

**Authors:** Aslan Abivardi, Matthew Webster, Paul McCarthy, Fidel Alfaro-Almagro, Lav Radosavljevic, Karla L. Miller, Saad Jbabdi, Mark W. Woolrich, Weikang Gong, Christian F. Beckmann, Lloyd T. Elliott, Thomas E. Nichols, Stephen M. Smith

## Abstract

Population-scale neuroimaging allows for novel biological discovery, but voxelwise analyses are computationally paralyzing and noisy, whereas imaging-derived phenotypes discard crucial spatial detail. We introduce PANDORA (Population Archive of Neuroimaging Data Organized for Rapid Analysis), a data-adaptive modelling platform designed to resolve this trade-off. PANDORA has encoded brain MRI data comprising 98 sub-modalities from over 80,000 UK Biobank participants in a highly efficient supervoxel representation. By performing statistical regressions directly within this compressed embedding, PANDORA reduces storage by up to 99% and accelerates computation 10-fold, while acting as a spatial denoiser to enhance statistical power. PANDORA also includes the full-resolution voxelwise ground-truth data, curated imaging confound variables, and a fast analysis tool achieving whole brain, voxelwise population-level regression in seconds to minutes. We showcase PANDORA’s ability to reproduce known patterns and reveal new associations including trauma, anxiety/depression, autism polygenic scores, and *EPHA3*.

---

With multimodal MRI from over 80,000 participants, UK Biobank (UKB) brain imaging has advanced the field of population neuroimaging to an unprecedented scale^1^. With increasing dataset size, however, come major computational challenges. From the six structural and functional imaging modalities, the core UK Biobank processing pipeline^2^ derives 98 sub-modalities (e.g., fractional anisotropy and mean diffusivity in white matter tracts from diffusion data, and grey-matter density maps and raw intensities from the T1-weighted images), greatly expanding the size, complexity and potential of the resource.

This has introduced a critical tension. On one hand, voxelwise analyses preserve high-resolution spatial information but are increasingly computationally paralyzing at the scale of 80,000 subjects and plagued by high noise. On the other hand, imaging-derived phenotypes (IDPs, pre-calculated summary measures from pre-specified brain regions^2^) offer manageable data dimensionality, but at the cost of discarding the rich, fine-grained spatial patterns that often characterize individual differences and disease pathology.

To resolve this trade-off, we introduce PANDORA (Population Archive of Neuroimaging Data Organized for Rapid Analysis). PANDORA is a data-adaptive modelling platform that bridges the gap between the raw complexity of voxels and the radical simplification of IDPs. By projecting multimodal brain MRI data into an optimized supervoxel embedding and performing statistical regressions directly within this space, PANDORA achieves massive data compression (up to 99% storage reduction) and a 10-fold acceleration in statistical computation. The PANDORA supervoxels are linear embeddings using very high-dimensional independent component analysis; a supervoxel can be thought of as a “soft clustering” of multiple voxels, where the voxels co-vary across subjects. The use of high-dimensional independent component analysis (ICA^4^) ensures that the supervoxels are spatially focal and biologically interpretable.

Previously, voxel-level image analysis with UKB data faced major practical barriers requiring expert intervention. For a single modality, users needed to retrieve one ZIP file per subject (i.e., at least 82,000 ZIP files), unpack them, and extract or derive the relevant sub-modality image. Creating analysis-ready data then required substantial new data curation, including modality-specific preprocessing, quality control to identify and robustly handle outlier subjects and voxels (identified in the context of the whole population dataset), normalization, and spatial transformation into common co-ordinate space. Even after this curation, population-level voxelwise modelling at UKB scale remains computationally challenging in I/O, memory and runtime. PANDORA removes these barriers by collating each sub-modality into a single HDF5 file and providing carefully curated voxel and supervoxel data. A PANDORA file can be fed directly into multi-subject regression against non-imaging variables or into deep neural network training. Regression is carried out using a new analysis tool that leverages the huge computational efficiency provided via the supervoxel embedding, while still giving voxelwise statistical outputs.

In this paper, we first describe the PANDORA UKBv1 resource, including the advanced general and modality-specific preprocessing pipeline used to generate analysis-ready data, with variance-based masking, data standardization, multi-stage outlier removal, and subsequent data-adaptive compression into supervoxels. We then validate the supervoxel representations by quantifying information retention in terms of full-resolution variance explained compared to the existing multimodal IDPs. Building on this, we describe our supervoxel-based regression framework, in which model estimation is performed in the encoded supervoxel space, with voxel- and grayordinate-level statistics recovered via supervoxel-to-voxel mappings. The mathematical details are given in Methods, and we summarize the implementation in our *fsl_glm* software tool, benchmarking its statistical power, runtime and memory use against conventional full-resolution mass-univariate regression.

Finally, we conduct four multimodal imaging experiments designed to showcase the utility of PANDORA across distinct neuroscientific domains: (1) lifetime trauma, as an environmental exposure with widespread multimodal effects; (2) anxiety and depressive symptom dimensions, probing sensitivity to overlapping but distinct affective phenotypes; (3) a single-nucleotide polymorphism (SNP) in the *EPHA3* gene, providing a case study in high-resolution voxelwise characterization of a developmental axon-guidance-related variant; and (4) polygenic scores for early-versus late-diagnosis autism^3^, testing whether these distinct genetic architectures have dissociable whole-brain signatures in UKB. Further analyses on smoking, alcohol and coffee are reported in Supplementary Results.

Crucially, PANDORA opens up entirely new avenues of investigation to researchers without deep imaging analysis expertise. Previously, these researchers could only use IDPs that capture relatively crude anatomical regions, yet voxelwise analyses have repeatedly demonstrated insight that goes far beyond IDPs by segmenting substructures and linking different brain regions^1, 4^. The combination of data curation, ease-of-use and efficiency offered by PANDORA means that any researcher with a knowledge of anatomy can now probe the richness of UKB brain imaging with fine spatial precision. Taken together, these advances position PANDORA as a transformative resource for fast, voxelwise discovery across environmental, symptomatic and genetic dimensions of brain variation.

## Results

### Resource description

PANDORA UKBv1 is a large-scale, analysis-ready neuroimaging archive built from data encompassing N=81,939 UKB participants and aggregating 98 sub-modalities (e.g., diffusion tensor imaging (DTI) metrics) spanning six MRI modalities across structural, diffusion, and functional imaging (Supplementary Table 1).

For each sub-modality, we provide full-resolution data as a subject × voxels matrix, by vectorizing (unwrapping) each participant’s 3D image into a single row. We also provide two reduced representations to enable scalable analysis: subject × supervoxel matrices at 1K and 10K supervoxels, with accompanying supervoxel-to-voxel spatial maps for visualization and interpretation.

All outputs are stored in HDF5 for broad compatibility (Python, MATLAB, C/C++) and fast I/O, with array layouts optimized for high-throughput streaming and parallel reads using the fast (stride-1) dimension.

Supervoxels deliver major storage reductions relative to full-resolution matrices across sub-modalities: a reduction factor of 35–79 for 1K supervoxels (reducing 22–765 GB to 0.6–9.7 GB) and 4–8 for 10K (to 5.8– 96.4 GB)—equivalently, disk usage drops by as much as 99% and 87%, respectively.

PANDORA also includes two preprocessed imaging confound matrices (for use in deconfounding later analyses, spanning effects like head size, sex, age and head motion): a larger covariate set (222 covariates capturing technical and subject-level variability) and a smaller 20-covariate set spanning the most important confounds, for analyses in smaller UKB subject subsets.

### Supervoxels preserve substantially more variance than IDPs

We quantified information retention for 1K- and 10K-supervoxel encodings and benchmarked them against modality-specific IDPs (and also the full set of >5K multimodal IDPs) by computing the percent of full-resolution voxel-level variance explained by each representation.

On average, 10K-supervoxels captured 95.9% ± 6.2% (range 63.0–100.0) of full-resolution variance, 1K-supervoxels 58.4% ± 10.2% (range 32.5–95.3), modality-specific IDPs 13.2% ± 7.0% (range 0.8–47.1), and multimodal IDPs 19.1% ± 9.9% (range 7.6–54.3). Thus, both supervoxel encodings substantially outperform IDPs, with the 10K representation closely approximating full-resolution data. Fig. 1A illustrates results across modalities.

**Figure 1.**
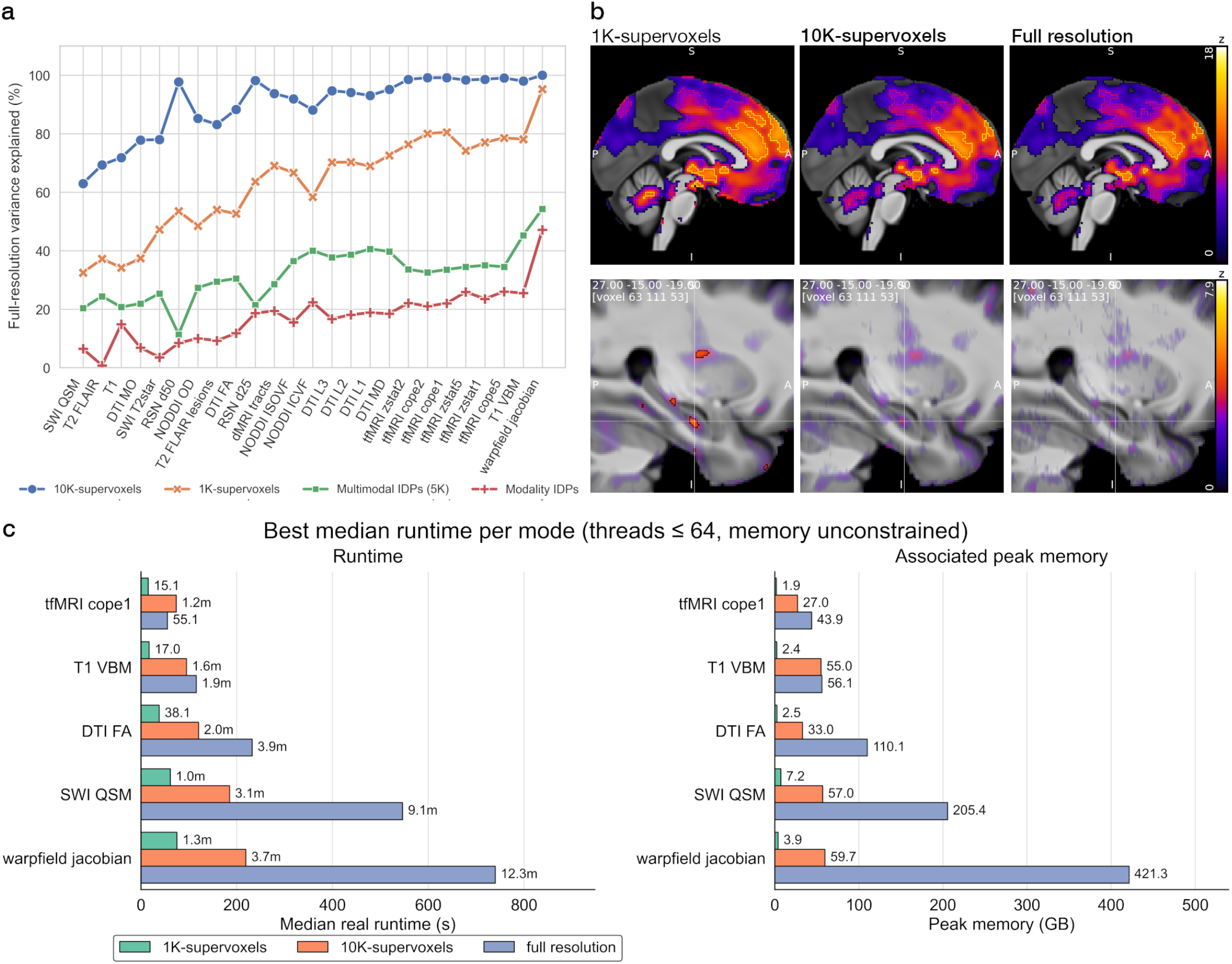
PANDORA supervoxels retain high full-resolution information while improving sensitivity and computational efficiency. **a. Image variance explained by supervoxels vs. IDPs across modalities.** For each of the 98 imaging sub-modalities, we show the percentage of voxel/grayordinate-level variance captured by 1K-supervoxels, 10K-supervoxels, modality-specific IDPs and multimodal (i.e., pooling all) IDPs. Across modalities, supervoxel encodings capture substantially more variance than IDPs, with 10K-supervoxels closely approximating full-resolution data, and 1K retaining a majority of the signal. Variance explained is modality-dependent, with inherently noisier sub-modalities (e.g., QSM, DTI MO) showing lower values and smoother or lower-resolution maps (e.g., VBM, warpfield Jacobian, CIFTI fMRI) showing the highest. **b. Example association maps and spatial fidelity**. <u>Upper panels</u>: Statistical maps (z-statistic) of association of past tobacco smoking (UKB ID: 1249) with decreased VBM signal, thresholded at hierarchical FDR levels 0.023 (colored overlay with black outline), 2.9 x 10^-7^ (red outline) and 7.6 x 10^-24^ (yellow outline); hierarchical FDR accounts for inference over 98 sub-modality maps. Supervoxels, especially 10K (Fig. 2), show high spatial convergence with the full data, while power increases from full-resolution to 10K-to 1K-supervoxels. Peak z-values for 1K-, 10K-supervoxels and full-resolution were at -17.6, -16.2 and -16.0 in left putamen, while z-statistic for left putamen gray matter volume IDP (UKB ID: 25882) was at -11.7. <u>Lower panels</u>: Denoising and data-adaptive compression enables detection of weaker effects. A positive association between neuroticism score and anterior hippocampal QSM, shown here, is only detectable using 1K-supervoxels (mapFDR-corrected p < 0.05, black outline), with a subthreshold denoising gain already apparent from voxels to 10K-supervoxels. **c. Runtime and memory benchmarking**. 1K-supervoxel analysis is much faster and more memory efficient than the other options. The advantage of 10K-supervoxels over full voxels becomes evident for sub-modalities with higher voxel counts. (See full benchmarks across parameter settings and when using more limited computational resources in Supplementary Materials).

**Figure 2.**
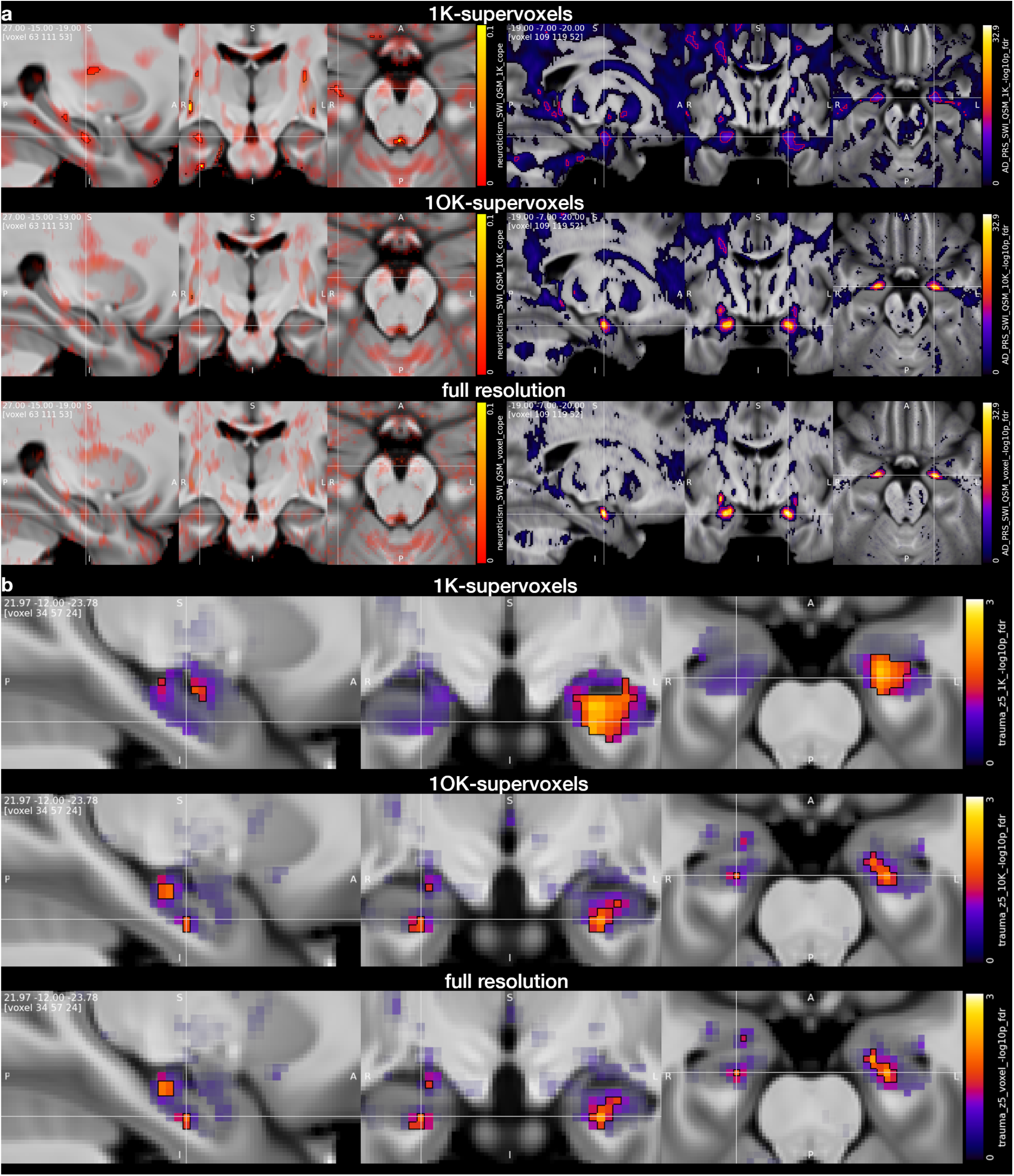
1K- or 10K-supervoxels: which to choose? **a.** The higher statistical power of the 1K-supervoxels can detect subtle effects, which may otherwise not reach significance, as seen on the left for an increase in right anterior hippocampus QSM associated with neuroticism score (UKB ID: 20217). Highly localized effects, however, as seen on the right for an Alzheimer’s disease polygenic risk score (UKB ID: 26206) association with centromedial amygdala QSM, may become weaker than when using 10K- or full resolution analyses. **b**. 10K-supervoxels exhibit high spatial accuracy in comparison with full-resolution analyses, illustrated here for the effect of cumulative traumatic life-events in an emotion task (faces–shapes contrast in the functional MRI data). <u>Visualization and significance thresholds</u>: Voxel color-coding reflects β-maps (2A left) or -log_10_(p) maps (2A right, 2B), with non-significant voxels showing color strength modulated by the Z-statistic map (in order to be able to visualize sub-threshold effects). Statistical significance is signified by contour outlines; 2A left and 2B are thresholded at map FDR corrected p < 0.05 (black) and p < 0.001 (yellow). 2A right is thresholded at hierarchical FDR (hFDR)-corrected p < 0.023 (black), 2.9 x 10^-7^ (red) and 7.6 x 10^-24^ (yellow) with hFDR accounting for inference over 9 sub-modality maps.

Consistent with expectations, modalities with higher voxel counts and/or greater inherent noise (e.g., quantitative susceptibility mapping (QSM), DTI tensor mode MO) showed lower explained variance. In contrast, lower-resolution CIFTI data and smoother maps (VBM, warpfield Jacobian) resulted in higher variance explained.

Analysis of shared explained variance between modality-specific IDPs and supervoxels indicated that, for most modalities, the voxel-space variance explained by IDPs largely lies within the subspace captured by supervoxels, with higher overlap for 10K than 1K supervoxels (median overlap: 92.3% for 10K vs. 85.3% for 1K; Figs. S6.1–S6.2). Conversely, supervoxels, especially 10K, explained substantial voxel-space variance not captured by IDPs, consistent with generally low IDP-specific explained variance.

### Supervoxel-regression: runtime and memory benchmarks

PANDORA includes a new supervoxel regression tool (packaged as an expansion of FSL’s *fsl_glm* univariate regression tool) that fits models in the encoded subspace, optimizing speed and memory efficiency, while returning full-resolution statistical maps that are equivalent to having reconstructed the full subjects × voxels data matrix from the supervoxel representation followed by voxelwise regression.

In five representative sub-modalities spanning a wide range of voxel/grayordinate counts, 1K-supervoxel regression reduced wall-clock (real) runtime by up to 10× relative to full-resolution voxelwise regression, on a 64-core node under unconstrained memory (threads ≤ 64; Fig. 1C). 10K-supervoxels also provided substantial wall-clock speedups, with the largest gains in higher-resolution modalities (3×), where voxelwise regression incurs substantially more voxel-space computation and data movement.

Looking beyond wall-clock time, user CPU time (core-seconds; total user-space CPU time summed across threads) showed even larger differences: 1K-supervoxels reduced core-seconds by more than two orders of magnitude relative to voxelwise regression, consistent with substantially less total compute performed in voxel space (Supplementary Fig. S7). This advantage persisted under resource-limited, cloud instance-like cost-effective settings (for instance 4 cores, 32 GB RAM; Supplementary Fig. S8), where the largest sub-modality completed in <1.5 minutes with 1K-supervoxels, compared with 80 minutes at full resolution (and 25 minutes for 10K-supervoxels).

Finally, peak memory usage was substantially reduced for 1K-supervoxels and remained lower for 10K-supervoxels than for voxelwise regression across modalities and settings (Fig. 1C; Supplementary Figs. S7–S9). In practice, this means analyses can be run quickly on smaller (and therefore less costly) compute instances while remaining performant. Peak memory depends on runtime settings, particularly thread count and chunk size.

### Supervoxel-regression: spatial fidelity and statistical power

Across 1K- and 10K-supervoxel and full-resolution GLM experiments (using *fsl_glm*), 10K-supervoxels provided a spatially highly accurate substitute for full-resolution regression, whereas 1K-supervoxels delivered greater power to detect or confidently exclude weaker, distributed effects, in many cases retaining most of the detailed spatial structure observed at 10K. This power gain is consistent with supervoxels imposing a data-adaptive low-rank representation that suppresses noise while preserving dominant spatial signal.

Figure 2A highlights the benefits and limitations of 1K-supervoxels: a weak effect in anterior hippocampal iron deposition associated with increased neuroticism score is detected only with 1K-supervoxels, whereas a spatially specific strong association in the central amygdala does not reach the significance levels observed in 10K or full-resolution analyses. On the other hand, Figure 2B shows the full-resolution results have near-perfect spatial agreement with the 10K-supervoxels, but inferior spatial agreement with the 1K-supervoxels.

To quantify sensitivity more broadly, we compared statistical power across all 98 imaging sub-modalities for the effect of past tobacco smoking (UKB ID: 1249); we measure power with squared z-statistics, aggregated with either a sum or average (see Fig 3). Across modalities, 1K-supervoxels yielded the highest power, followed by 10K-supervoxels, with full-resolution analyses typically lowest. These differences were highly significant at the omnibus level (Friedman test, 3 paired methods across 98 sub-modalities: statistic = 147.57, p = 9.02 × 10^−33^). Post-hoc paired Wilcoxon signed-rank tests (Pratt; Holm-corrected) confirmed consistent ordering across modalities: 1K-supervoxels outperformed 10K-supervoxels in 95/98 sub-modalities (W = 13.0, *p*_Holm_ = 3.73 × 10^−17^ ; median Δ = 1.34 × 10^4^), and 1K-supervoxels outperformed full-resolution in 93/98 (W = 192.0, *p*_Holm_ = 4.96 × 10^−15^; median Δ = 1.21 × 10^4^). 10K-supervoxels outperformed full-resolution in 84/98 sub-modalities (W = 323.0, *p*_Holm_ = 9.31 × 10^−14^; median Δ = 380.6). Together, these analyses establish a robust power advantage of supervoxel regression, especially 1K, across a broad range of imaging modalities.

**Figure 3.**
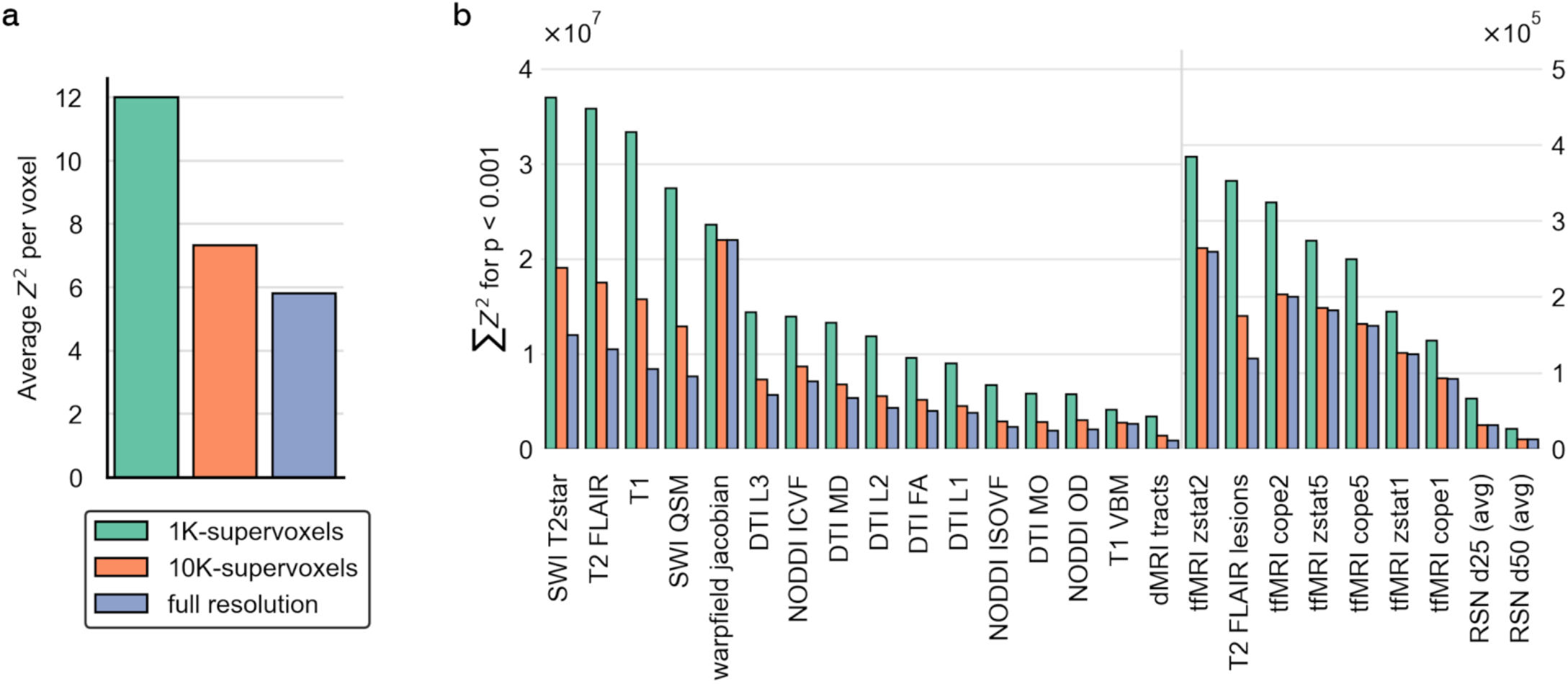
Statistical power across 98 imaging sub-modalities (associations with past tobacco use). **a.** Power across sub-modalities, as measured by average squared z-statistic per voxel/grayordinate (unthresholded) was highest for 1K-supervoxels, followed by 10K supervoxels and full-resolution. **b**. Individual sub-modality comparisons (summed Z^2^ across voxels/grayordinates with uncorrected p < 0.001) showed a consistent and pronounced advantage for 1K-supervoxels: 1K outperformed 10K in 95/98 modalities and full resolution in 93/98, indicating a near-universal power gain from stronger data-adaptive compression/denoising. Improvements were especially marked for noisier and higher-resolution sub-modalities (e.g., SWI and T1), where the gap between 10K and full resolution was also larger. Differences between methods were significant with Friedman’s test (p < 10^−32^; see body text). For visualization, panel B uses a split x-axis to show lower- and higher-power sub-modalities in the same plot.

Figure 1B illustrates this effect for smoking in VBM and neuroticism in QSM. For both modalities, significance increased from full-resolution to 10K, and further to 1K. The denoising benefit is particularly apparent for QSM, while spatial agreement is near-perfect between 10K-supervoxels and full-resolution maps.

### Experimental results

To illustrate PANDORA’s utility across diverse studies, we report four representative analyses spanning environmental exposure, symptom dimensions, single-variant association, and polygenic scores, each across multiple imaging sub-modalities. Unless otherwise noted, results are from 10K-supervoxel analyses, because for the present study we prioritized high spatial accuracy and closer approximation to full-resolution maps; full statistical maps and peak coordinates are provided in the Supplementary Results, which also include a fifth analysis of lifestyle exposures (alcohol, smoking, and coffee). All analyses adjusted for 222 imaging confounds plus experiment-specific covariates (Table 1; Methods). For each experiment, sub-modalities were selected *a priori* to balance biological coverage, interpretability, and multiple-testing burden (see Methods: Applications of PANDORA).

**Table 1.** Experimental characteristics for the five GLM experiments.

| *Experiment* | **N subjects** | **Sex (% f.)** | **Age (mean ± SD)** | **Confounds** | **N sub-modalities/ contrasts** |
| --- | --- | --- | --- | --- | --- |
| 1: Cumulative Lifetime Trauma | 60,345 | 54.7 | 66.5 ± 7.8 | Townsend deprivation, confounds_all | 13 / 2 |
| 2: Anxiety vs. Depressive Symptom Load | 67,009 | 54.2 | 66.4 ± 7.8 | Townsend deprivation, confounds_all | 12 / 8 |
| 3: *EPHA3* (rs987748) | 67,192 | 52.5 | 66.8 ± 7.9 | 40 genetic PCs, confounds_all | 16 / 2 |
| 4: Early- vs. late-autism polygenic scores | 67,192 | 52.5 | 66.8 ± 7.9 | 40 genetic PCs, confounds_all | 10 / 8 |
| S1: Alcohol, Coffee and Smoking | 78,277 | 53.4 | 66.4 ± 7.9 | Townsend deprivation, ranked education, confounds_all | 13 / 14 |

#### Experiment 1: Effect of cumulative trauma on brain

Cumulative trauma, defined as a composite of multiple adverse-life-event items and adjusted for deprivation, was significantly associated with 12/13 tested sub-modalities. We report 10K-supervoxel results unless stated otherwise and classify evidence as strong for associations surviving hierarchical FDR correction over all tested sub-modalities at *p*_*hFDR*_ < 0.001, and moderate *p*_*hFDR*_ < 0.01, with minimum corrected significance threshold being *p*_*hFDR*_ < 0.05. Key modality-specific findings are summarized below, with selected highlights in Fig. 4 and full anatomical results and peak statistics in the Supplementary Results (Figs. S1.1–S1.2; Supplementary Tables).

**Figure 4.**
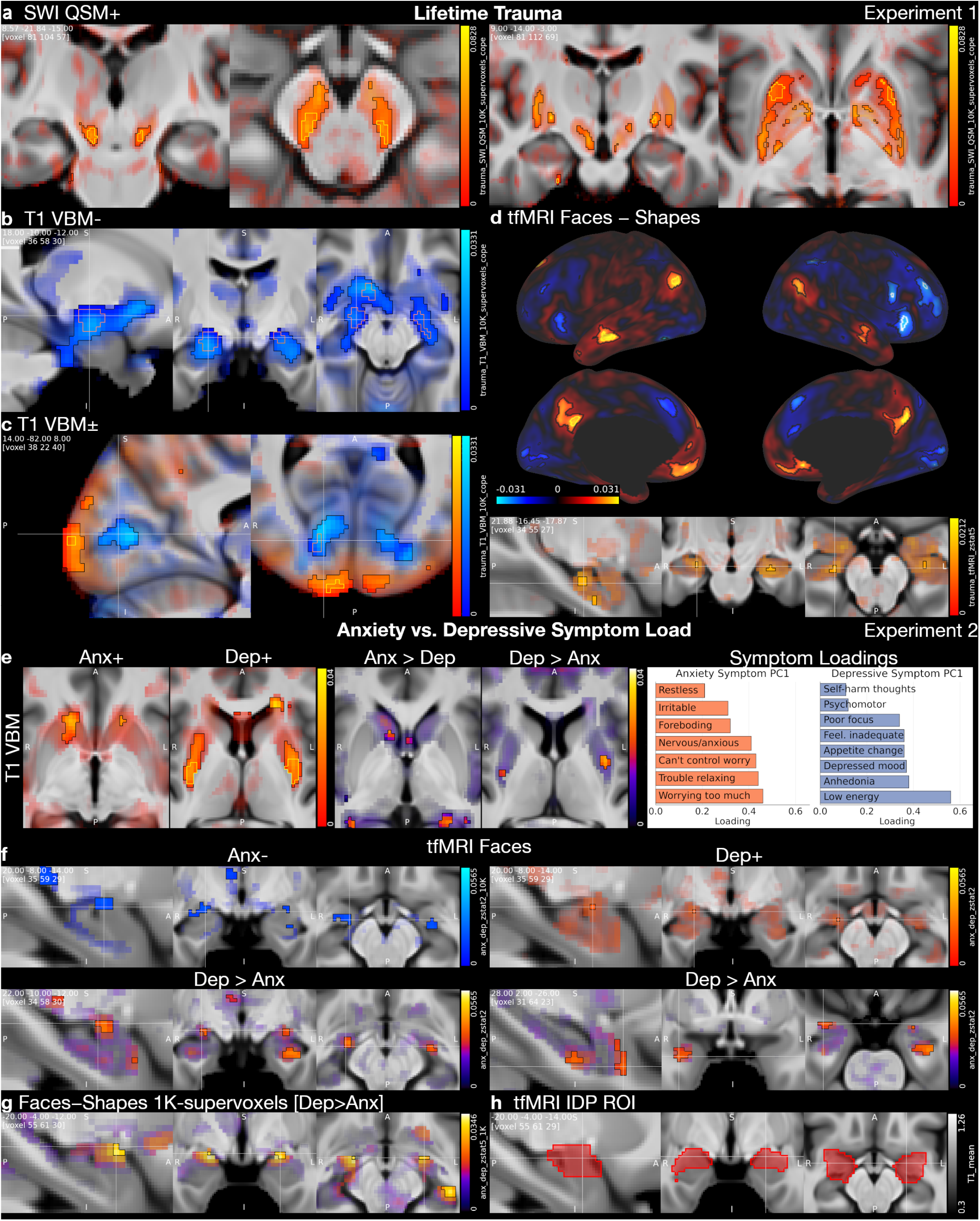
Experimental highlights 1 and 2 (10K-supervoxels unless stated otherwise). **Experiment 1 (a–d). a.** Lifetime cumulative trauma tracked higher quantitative susceptibility mapping (QSM) signal in the substantia nigra and striatum, indicative of iron accumulation in dopaminergic circuitry, a pattern consistent with reports of increased Parkinson’s disease risk in PTSD^5^. **b**. In grey matter, the strongest association localized to the amygdalo-hippocampal complex (peak right dorsocaudal amygdala: Z = −7.1; right amygdala IDP [UKB ID 25889] for comparison: Z = −4.1), underscoring a robust, spatially specific amygdala effect despite heterogeneity in prior clinical reports^**6**^. **c**. Calcarine effects followed the V1 retinotopic axis: greater VBM in posterior V1 (central visual field) but reduced along more anterior calcarine cortex (peripheral visual field), consistent with PTSD symptomatology in which hyperarousal biases visual processing towards threat monitoring and attentional narrowing^7^. **d**. During the emotion task, higher trauma load was associated with increased default mode network (DMN) and anterior hippocampus activation (and right vl-/dlPFC deactivation) in the faces−shapes contrast, counter to the typical DMN deactivation reported in emotional-face viewing^8^ and consistent with anterior hippocampal involvement in avoidance behavior^9^—a core PTSD symptom dimension. **Experiment 2 (e–h)**. Anxiety and depressive symptom load dissociated along striatal-amygdala axes. **e**. Positive striatal VBM associations separated topographically (depression in posterior putamen; anxiety in anterior caudo-putamen), with direct comparison analyses recapitulating this spatial dissociation. **f**. Amygdala task responses diverged in sign, with anxiety linked to reduced amygdala reactivity and depression to increased reactivity to faces and shapes (Supplementary Results); depression–anxiety and anxiety–depression contrasts further localized subregional amygdala effects. **g**. A 1K-supervoxel example illustrates the gain in power, revealing subtle faces–shapes effects that were not detected in the 10K analysis. **h**. For comparison with these localized effects, we show a group-level amygdala activation ROI used to derive UKB IDPs (mask5a.nii.gz; e.g., UKB ID 25052). <u>Visualization convention (Figs. 4 and 5)</u>: β-maps were thresholded using hierarchical FDR (hFDR) across tested sub-modalities within each experiment (see body text). Positive associations are shown in red and negative associations in blue. Outlines indicate corrected significance at two levels: black for p_hFDR_ < 0.05, and yellow (positive) or orange (negative) for p_hFDR_ < 0.001 (white for cortical surface plots). For non-significant effects, color intensity is modulated by the Z-statistic.

**Figure 5.**
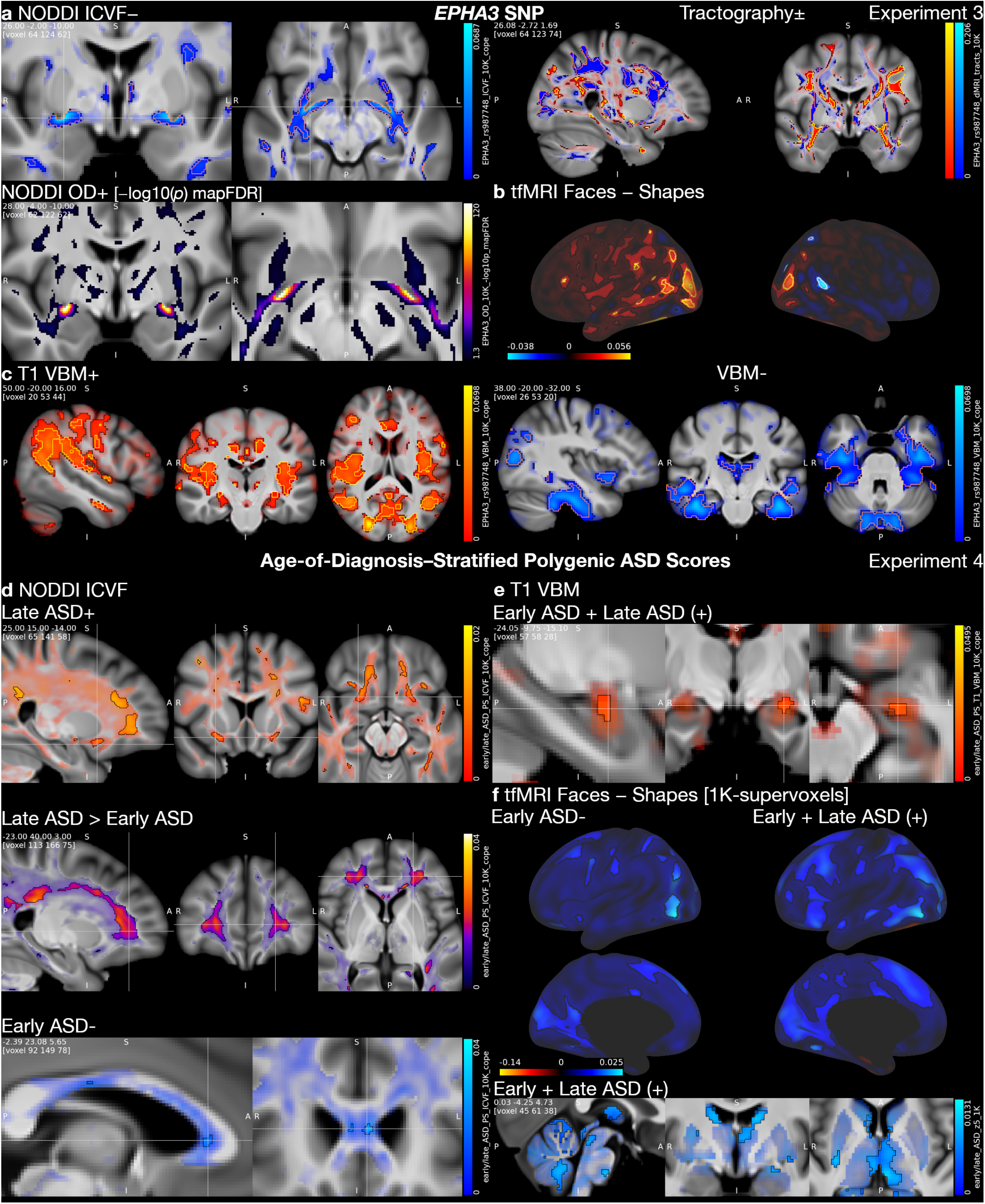
Experimental highlights 3 and 4 (10K-supervoxels unless stated otherwise). **Experiment 3 (a–c). a.** Carriers of the minor variant for SNP rs987748 in the axon-guiding gene EPHA3 exhibited reduced microstructural integrity in the anterior commissure across imaging markers (e.g., ICVF and OD) with exceptionally robust evidence (OD; mapFDR-corrected −log_10_(p) > 100). This focal effect was accompanied by fine-scale variation in pathway organization–apparent here on tractography and mirrored across diffusion maps–with closely offset positive and negative segments consistent with marked shifts in tract course/positioning and local fiber architecture. **b**. Consistent with these structural alterations, the emotion task (faces–shapes contrast) showed a striking left-lateralization of temporo-parietal and subcortical (Supplementary Results) responses within the face-perception network. **c**. VBM, in turn, revealed distributed grey-matter differences, with increases spanning fronto-parieto-occipital cortex and decreases weighted toward temporal lobe, especially fusiform and middle temporal cortex. **Experiment 4 (d–f). d**. In our main analysis, early- and late-diagnosis ASD polygenic scores dissociated in white-matter microstructure, with late-ASD showing increased frontal neurite density (ICVF) and the late-ASD > early-ASD contrast reinforcing the difference, whereas higher early-ASD scores were characterized by reduced ICVF in anterior corpus callosum. **e**. Interestingly, the two uncorrelated scores exhibited an additive effect on increased amygdala volume. **f**. In a final illustration of the supervoxel sensitivity trade-off, the higher-power 1K-supervoxel analysis additionally picked up a distributed reduction in faces–shapes responses for early-ASD spanning cortex and subcortex that was not apparent in the standard 10K maps. <u>Visualization convention</u>: see Fig. 4.

##### SWI

QSM showed strong evidence for positive trauma association in bilateral substantia nigra, striatum, amygdalo-striatal transition area, left medial forebrain bundle, and right entorhinal cortex, while strong-evidence decreases were seen in lateral geniculate nuclei, left thalamus (VLpv), right lateral hypothalamus/substantia innominata, internal/external capsule, temporal cortices, and cerebellum. T2^*^ showed widespread strong-evidence positive associations in parieto-occipital white matter (WM), cingulum, along dmPFC, peaking at left fusiform GM/WM boundary. Negative associations were strongly supported in cerebellum, right anterior insula, and left superior temporal gyrus.

##### VBM

Trauma associated with decreased local grey matter (GM) volume in bilateral amygdalo-hippocampal complex, right substantia innominata, nucleus accumbens and left thalamus (VLp/VLa), and calcarine sulcus (anterior V1) with strong evidence. We found moderate-to-strong evidence for positive association in posterior V1 and ventromedial prefrontal cortices (vmPFC).

##### Task functional MRI (tfMRI zstat)

For the faces−shapes contrast, there was strong evidence for a positive association in the default mode network (DMN) including anterior hippocampus and left middle temporal gyrus (MTG). Specifically, relative to the group-level contrast, higher trauma load associated with stronger faces>shapes responsiveness in precuneus, MTG, and left mPFC, and attenuated shapes>faces effect in right anterior subgenual ACC (BA25). Increased shapes>faces activations were seen in right vl-/dlPFC, anterior insula, and angular gyrus. Faces main effect associated with reduced deactivation in right precuneus (moderate-evidence) and significantly increased deactivation in right supramarginal gyrus.

##### DTI and neurite orientation and density imaging (NODDI)

ICVF showed positive association limited to left putamen (moderate-evidence) but robustly supported a global negative shift throughout supra- and infratentorial WM (peak centrum semiovale: −log_10_(*p*)_mapFDR_ > 15). Evidence strongly supported corresponding MD increases and FA decreases across major tracts.

##### Resting-state networks (RSNs)

In the salience-like RSN d25-9 (SN) component, we found strong-evidence negative associations in left paracentral lobule, bilateral cuneus, and right dmPFC (reduced coupling), and left MTG (increased anticorrelation). In somatomotor-like RSN d25-13 (SMN), trauma associated negatively in the right precentral/opercular region (increased anticorrelation; moderate-evidence). Within RSN d25-2 (DMN), dmPFC and left precuneus exhibited reduced coupling, with no significant positive associations.

#### Experiment 2: Dissociating anxiety and depressive symptom dimensions

Anxiety and depressive symptom load (first principal components of symptom items from two online questionnaires) showed dissociable effects across 12/12 sub-modalities. We summarize key associations and contrasts for these two symptom dimensions. We also revisited prior UKB reports of absent Hariri-task amygdala effects^10^ by testing amygdala voxel-level reactivity rather than region-of-interest/IDP summaries. Fig. 4 shows selected highlights; full results are in S2.1–S2.6.

##### VBM

Anxiety score was linked to greater grey-matter volume in right cerebellum (moderate-evidence), with significant positive associations in bilateral anterior caudate (also contrasting against depression in right caudate). Depression score showed moderate-to-strong evidence for positive associations in bilateral putamen, left dlPFC, right frontal operculum/anterior insula, optic chiasm/hypothalamic region, left V1, and bilateral cerebellum. Putaminal volumes were positively associated with the depression−anxiety load difference.

Anxiety score showed moderate-to-strong evidence for negative associations along vm-/dmPFC and cingulate sulci. Depression score showed widespread GM strong-evidence negative associations in bilateral amygdalo-hippocampal complex, straight gyri, subgenual ACC, posterior insula, and pulvinar.

##### tfMRI (zstat)

Depression score was associated with higher right amygdala responses to both shapes (basolateral; reduced deactivation) and faces (centromedial; increased activation). In contrast, anxiety was associated with lower amygdala responses, i.e., greater right basolateral deactivation to shapes, and reduced responses to faces in centromedial and right lateral amygdala alongside increased basolateral deactivation. Depression−anxiety load difference was related to increased basolateral responses to shapes (decreased deactivation) and centromedial responses to faces (increased activation). No effect emerged for faces−shapes in the primary 10K-supervoxel analysis; however, 1K-supervoxel analysis detected a positive association in the left (anterior) centromedial amygdala, indicating increased faces>shapes responsiveness with higher depression–anxiety load difference. Anxiety score further exhibited strong-evidence distributed negative associations (e.g., temporo-parietal junction and PFC) with faces and shapes main effects, while depression score negatively associated in sensorimotor cortices, striatum and cerebellum.

##### SWI

Positive depression score associated with QSM in bihemispheric posterior insula, Heschl’s gyrus and left cingulate sulcus (strong-evidence), while negative associations were seen in left bed nucleus of the stria terminalis (BNST)/anterior commissure.

Anxiety score showed strong-evidence positive association with T2^*^ in bilateral dlPFC extending into the GM/cerebrospinal fluid (CSF) boundary region. Depression positively associated with T2^*^ in subcallosal area, inferior temporal gyri and right parieto-occipital WM/GM boundary. Depression scores were negatively associated with T2^*^ across widespread cortical regions and along sulci (peak left dlPFC: −log(*p*)_mapFDR_ > 10). Strong-evidence negative association of depression score was seen in anterior hippocampus. Note that brain/CSF boundary effects should be interpreted cautiously as they may reflect residual geometric effects (misalignment).

##### RSNs

Within RSN d25-2 (DMN), anxiety score showed moderate-evidence positive association with right precuneus (increased coupling) and negative association with right dmPFC and supramarginal gyrus (increased anticorrelation). We observed no association for depressive symptoms with DMN. Within RSN d25-9 (SN), depression score exhibited strong-evidence positive association with left anterior precuneus (increased coupling), and moderate-evidence negative association with left premotor/motor cortex and cuneus (reduced coupling).

#### Experiment 3: EPHA3 gene (rs987748)

Robust rs987748 effects were detected across all 17/17 tested sub-modalities, with distributed patterns along major white-matter pathways and highly localized signals in anterior commissure (AC), plus additional cortical/subcortical associations. Below we report key associations for the minor allele (Fig. 5, selected highlights; S4.1–S4.5, full results).

##### VBM

The minor allele showed strongest positive associations with GM in midline AC and temporal AC/uncinate fascicle (UF) region. Broad positive associations (strong-evidence) appeared across frontal, and most prominently parieto-occipital cortices. Negative associations were most robust for temporal lobes, especially fusiform cortices and middle temporal gyri.

##### DTI and NODDI

Microstructure associations were widespread and highly significant. Most consistent effects involved AC, bilateral LGN, arcuate and longitudinal fasciculi and temporo-occipital U-fibers. Across DTI measures, MD and FA showed region-specific bidirectional shifts, including increased MD and decreased FA in AC and LGN, with prominent effects in temporal-occipital WM, posterior callosal fibers, and cerebellar WM. NODDI metrics provided especially strong evidence for commissural effects, marked by very robust positive OD association in lateral AC (−log_10_(*p*)_mapFDR_ > 100) with co-localized negative associations in neurite density and isotropic fraction, and distributed ICVF/ISOVF alterations across temporo-occipital U-fibers, major association tracts, thalamic/optic radiations, and cerebellar/mesencephalic WM; and additional negative OD effects in select regions posterior to AC and posterior callosum.

##### tfMRI (zstat)

Robust activation differences were seen across shapes, faces, and the faces−shapes contrast, consistently involving fusiform and temporo-parietal regions (supramarginal/angular; parieto-occipital transition). Negative associations in all three z-statistic maps tended to cluster in middle and superior temporal gyri. The faces–shapes contrast was notably left-lateralized, with strongest positive effects in left occipito-temporal cortex and additional left-dominant effects extending into vl-/dmPFC and left putamen.

##### SWI

QSM was most robustly associated in AC, followed by left putaminal surface, superior temporal and fusiform cortices, right anterior hippocampus, Heschl’s gyrus, middle longitudinal fascicles, and occipital lobe WM. Negative associations were seen with highest evidence in internal capsule and periventricular white matter, in between cingulum bundle and corticospinal tracts.

##### RSNs

rs987748 was associated with coupling changes in DMN (RSN d25-2), a fronto-parietal network (RSN d25-6), and a language-like network (RSN d25-18). Most prominent effects localized to posterior midline and temporo-parietal hubs (e.g., PCC/precuneus, TPJ), with broader contributions from PFC and subcortex. Increases and decreases often appeared side-by-side, yielding paired “up/down” configurations rather than uniform shifts.

#### Experiment 4: Multimodal associations of early-vs late-diagnosis autism polygenic scores

Early- and late-diagnosis autism (ASD) polygenic scores (PS), derived from a recently published autism GWAS that separated genetic effects linked to earlier versus later age-at-diagnosis^3^, were found in 6/10 tested sub-modalities, revealing dissociable white-matter signatures and additive effects in amygdala (Fig. 5, selected highlights, Figs. S5.1–S5.3, full results).

##### DTI and NODDI

Results showed higher neurite density (ICVF) for late-ASD PS, with effects most apparent in frontal and fronto-basal white matter but also extending to forceps major, superior temporal white matter, and fusiform pathways. The late>early contrast was consistent with this pattern and expressed it more strongly, with a modest frontal/fronto-basal emphasis. Early-ASD PS instead showed negative ICVF associations in anterior callosal white matter and nearby longitudinal tracts. OD effects were smaller and more focal (e.g., positive association with late-PS in right uncinate fascicle). FA showed moderate negative associations with early-PS in anterior callosum, left cingulate bundle and across corticospinal tracts; Early-PS exhibited higher MD in left FAT with a broader early>late pattern centered on frontal lobe extending into parieto-occipital regions.

##### T1 VBM

The combined early+late PS were associated with greater volume in left amygdala and bilateral cerebellum. Moderate evidence indicated negative associations in left cerebellar GM for early-ASD PS; contralateral reduction was also significant. Left dlPFC and right putamen volumes were higher for late-ASD than early-ASD.

##### tfMRI (zstat)

Moderate evidence indicated increased deactivation of right precuneus to emotional faces for combined early+late PS (negative association), with significant effect also seen in left hemisphere. No significant associations were seen for shapes or faces−shapes using 10K supervoxels. Exploratory 1K analysis of faces−shapes supported widespread negative cortical and subcortical associations for early PS, i.e., a general shift towards shapes>faces responsiveness. More extensive associations were seen for combined early+late PS, while late PS alone was not significantly associated.

##### SWI

SWI was not associated with autism scores using 10K-supervoxels. Exploratory 1K-supervoxel analysis detected left mid/anterior hippocampus T2^*^ association with early-ASD PS (moderate evidence), and to a lesser extent with early+late PS. Early-ASD PS was associated with moderate-evidence QSM decrease in right posterior insula, while late-ASD PS was associated with QSM decrease in AC and left substantia nigra.

## Discussion

With PANDORA, we provide a major resource and computationally efficient framework for multimodal population neuroimaging using UK Biobank data. Alongside streamlined general and modality-specific processing and a convenient data layout, we have efficient compressed versions of the MRI maps, based on supervoxels. A core component is a regression formulation that naturally bridges encoding to full-resolution space analysis, thus leveraging memory and speed advantages of supervoxels. Quantitative and qualitative comparisons show that supervoxels are much faster, and often more powerful, than full-resolution univariate analyses, with distinct statistical and spatial behavior for 1K- and 10K-variants. The tool is implemented as an expansion of FSL’s *fsl_glm* in C++, prioritizing speed.

Across five experiments, we demonstrate real-world utility and potential of the resource, including: widespread brain changes associated with cumulative trauma; dissociable effects of anxiety vs. depressive symptoms; differential associations with smoking, coffee and alcohol (Supplementary Results); voxel-level mapping of *EPHA3* effects; and the first brain mapping contrasting polygenic scores derived for early-vs. late-diagnosed autism. Key findings are discussed below.

Comparing full-resolution (original data) variance explained by 1K-/10K-supervoxels vs. modality-specific and multimodal IDPs (∼5K), we found that supervoxels explain more variance than IDPs in every sub-modality. On average, 10K-supervoxels explained over 95%, 1K-supervoxels around 58%, and multimodal IDPs less than 20% of the variance. Importantly, variance explained does not account for different levels of noise in the data, where modelling 100% of the variance would therefore not be desirable. Inherently noisy sub-modalities such as SWI QSM or DTI MO (mode of anisotropy), for instance, exhibited among the lowest levels of explained variance.

We compared power in finding associations with smoking, for supervoxel and full-resolution data representations in all sub-modalities (Fig. 3). 1K-supervoxels outperformed 10K-supervoxels in 95 of 98 sub-modalities and full-resolution in 93 of 98; 10K-supervoxels outperformed full-resolution analyses in 84 of 98 sub-modalities. In noisy sub-modalities or when dealing with noise in the target phenotype, such an increase in power can result in identification of otherwise hidden effects (Fig. 2A). Direct comparison revealed a high spatial overlap between 10K-supervoxels and full-resolution results, which was not completely matched by the 1K-supervoxels. 1K-supervoxels may lack sensitivity for some of the most highly localized effects (Fig. 2AB).

Cloud storage and computing incur high costs on research projects and may impede full use of valuable resources such as UK Biobank. Analysis speed, which directly drives computing costs, adds further friction. Large datasets also raise memory demands and often require specialized strategies even on high-RAM machines.

PANDORA addresses these constraints in several ways. First, the supervoxel versions of the data achieve massive disk-space reduction of up to 98.7% (1K) and 87.2% (10K). Next, supervoxel-regression capitalizes on this advantage by estimating its model in encoding space, simultaneously reducing the amount of computation and avoiding build-up of the full-resolution matrix in memory. Benchmark analyses thus revealed a substantial speed-up across modalities of all sizes. Third, we reduce memory footprint via online computation and chunking, implemented for both supervoxel and full-resolution modes. To achieve fast read times and efficient downstream regression, we chose the disk layout (stride-1) to match the dominant access pattern during streaming and computation. Supervoxel modes stream only the spatial maps, sharply reducing I/O. To maximize speed, all modes are parallelized using the C++ standard library. We chose C++ over Python for computational efficiency and explicit control of memory and parallelization.

To demonstrate PANDORA’s broad value, we carried out five exemplar neuroscientific regression studies, and discuss here key findings from four (full results, including for the fifth study, are described in Supplementary Materials):

Lifetime trauma was robustly associated with increased tissue magnetic susceptibility (using quantitative susceptibility mapping, QSM) in substantia nigra, consistent with higher iron content (Fig. 4). Strikingly, posttraumatic stress disorder (PTSD) has been shown to increase Parkinson’s disease (PD) risk with reported hazard ratios between 1.48 and 3.46^5, 11^. While chronic restraint stress can trigger dopaminergic loss in substantia nigra in rats^12^ and nigral QSM is increased in PD^13^, to our knowledge, a direct anatomical link in humans has not been reported. Beyond substantia nigra, we observed strong evidence for increased QSM in bilateral putamen, a downstream site of PD pathology. A rat model of PTSD exhibited neuronal injury in the striatum caused by iron deposition^14^. Intriguingly, we also found decreased QSM in a thalamic region linked to PD tremor^15^, compatible with greater myelination or calcification.

With grey-matter local volume (VBM), we found strong evidence for grey-matter reduction in bilateral amygdalo-hippocampal complex. While hippocampal volume loss is consistently reported in PTSD^16^, findings for amygdala have been mixed^6^. Amygdala volumes have, e.g., been reported to be enlarged in war-veterans with PTSD, whereas greater combat exposure associated with smaller amygdala in the same study^17^. Another study from the same year, conducted in veterans with and without PTSD, found smaller amygdala volumes in PTSD and no association with trauma load^18^. Notably, smaller amygdala volume has been associated with PTSD severity in veterans and traumatized youths^19, 20^.

We further observed a retinotopic calcarine signature, with posterior V1 expansion alongside more anterior calcarine reductions, suggesting a “tunneled” visual emphasis on central information. This aligns with threat-monitoring-related attentional narrowing, in which processing is biased toward salient central cues at the expense of peripheral contextual detail^7^. Additional signatures implicating default-mode and approach– avoidance circuitry during negative face viewing are presented in Figure 4.

Recent work examined amygdala responses to negative faces (contrasting against shapes) in depression in UKB, but found no association^10^. We revisited this question of missing (ROI-based) amygdala activation in the Hariri emotion task^21^, using anxiety and depressive symptom principal components and voxel-level analysis, across all three z-statistic tfMRI functional contrasts. We found significantly increased reactivity for greater depressive symptom load in right amygdala to both shapes and faces, while anxiety related negatively to amygdala activation for both. Correspondingly, the contrast of depressive–anxiety symptoms was positively associated with basolateral amygdala activation to shapes and centromedial activation to faces.

Unsurprisingly, given that depressive load associated with both shapes and faces, we observed no association with the faces–shapes contrast. This, however, provided an illustration of higher sensitivity using 1K-supervoxels, which did detect a significant positive relation between depressive–anxiety load difference and centromedial amygdala responsiveness to faces>shapes.

VBM revealed a striking striatal dissociation between symptom dimensions: depressive load mapped to localized increase in posterior putamen, whereas anxiety load mapped to increase in anterior caudate. This pattern admits a circuit-level interpretation, with posterior putamen being preferentially embedded in somatomotor circuitry and anterior caudate aligning with frontoparietal circuitry (the main anxiety effect localized to the central head of the caudate, which preferentially couples with frontoparietal control circuitry; the anxiety>depression effect localized more medially, where it couples with medial prefrontal/default-network circuitry).^22^ Notably, clinical studies in major depression have been inconsistent, with the most recent large-scale meta-analysis of basal ganglia volumes^23^ failing to replicate earlier reports of striatal volume loss^24^.

In a third main experiment, we investigated the effect of the *EPHA3* SNP rs987748 on the brain. Ephrin signaling plays core roles in axon guidance, cell migration^25^, and tissue patterning^26^; *EPHA3* gene mutations have been implicated in glioblastoma^27^ and autism^28^.

Prior study of rs987748 has been limited: in our work using IDPs, expected minor-allele dosage was associated with occipital-lobe resting-state connectivity with the left hemisphere and an ICA-derived whole-brain connectivity measure (see S18.3)^29^, while a separate report described weak association of the major allele with fluid intelligence^30^.

Across diffusion sub-modalities and QSM, we observed the most robust evidence. Minor-allele count was associated with reduced microstructural integrity of anterior commissure (e.g., increased fiber dispersion; −log_10_(*p*)_mapFDR_ > 100) and temporal lobe connectivity via anterior commissure/uncinate fascicle. Conversely, tract integrity increased in mid-corpus callosum and in long-range temporo-parietal and parieto-occipital tracts. Notably, within the same pathways, positive and negative associations often appeared in nearby but shifted locations, consistent with fine-grained control of tract geometry and local fiber architecture.

These microstructural effects were accompanied by strong evidence for volumetric differences: the minor allele was linked to reduced temporal lobe and dorsal thalamus volumes, and increased volumes in fronto-parieto-occipital lobes. Functional analyses exhibited striking left-lateralization during face perception across cortical and subcortex, while resting-state networks showed distinct anatomical shifts in activity. Left-lateralized face processing has been reported in left-handed men^31^, while a nearby *EPHA3* SNP has been associated with hemispheric differences in language-network connectivity^32^. *EphA3* is highly expressed in the developing dorsal thalamus of rats^33^ and in mouse anterior commissure and corpus callosum^34^. While EphA3 has been implicated in mouse callosal axon segregation^35^ and *EphA4* is required for anterior commissure formation^36^, evidence connecting EphA3 to anterior commissure disruption (e.g., in *Olig2* knockout mice) has remained inconclusive^37^.

In a final experiment, we analyzed the effect of early- and late-diagnosis autism polygenic scores on the brain.

Zhang et al.^3^ reported that autism genetics differs by age-at-diagnosis, with partially distinct polygenic architectures and only modest genetic correlation between early- and late-diagnosed groups. An early-diagnosis factor was associated with early social/communication problems, whereas a late-diagnosis factor related to greater socioemotional difficulties and showed positive genetic correlations with ADHD and other psychiatric disorders. Using the age-of-diagnosis–stratified GWAS summary statistics, we constructed separate polygenic scores in UKB.

Our key finding was increased neurite density (ICVF) specific to late-autism polygenic score (PS), most pronounced in frontal white matter, and divergent from early-autism score. Early-autism PS instead showed reduced neurite density in corpus callosum. Altered frontal lobe connectivity has been repeatedly implicated in autism^38, 39^. In a previous mixed-age cohort (3–36 years), corpus callosum measures showed greater variability in midsagittal area in autism versus controls, and within autism, larger callosal area correlated with higher intelligence and faster processing speed^40^.

Prior work on amygdala volume in ASD generally reports early childhood overgrowth^41, 42^ and normal or reduced volumes in adolescence and adulthood^43, 44^. Curiously, only the combination of high early-*plus* late-autism scores was associated with greater left amygdala volume. A recent Hariri-paradigm study comparing 72 autistic participants and controls found no difference in mean amygdala activity during emotional face processing (specifically faces–shapes)^45^; we similarly observed no significant associations in the primary 10K-supervoxel analysis. 1K-supervoxels, by contrast, revealed a distributed negative cortico-subcortical signature for higher early-ASD polygenic load, consistent with a general shift toward reduced face-selective responding. Associations with combined early+late PS were even more extensive including amygdala, whereas late PS alone was not significantly associated. Taken together, these patterns illustrate how 1K-supervoxels can recover spatially diffuse effects that otherwise remain subthreshold.

### PANDORA in practice: recommendations and future directions

Given their high spatial fidelity, lower computational burden, and increased statistical power in most cases, 10K-supervoxels can act as replacement and arguably be given preference over full-resolution analyses. The case for 1K-supervoxel analyses is more complex: frequent power gains may come at the cost of localization. Further, the huge gain in computational efficiency allows to quickly run a 1K-supervoxel analysis across all 98 sub-modalities, which makes exploratory analyses highly efficient and effective, but of course it is important to be aware of the dangers of data-trawling and circularity when making the final inferences (e.g., with 10K-supervoxels).

Future release plans include modality-specific tuning of supervoxel component number, cross-modal encodings, revisiting intensity normalization for T1/T2, adding grey-matter diffusion maps, and expanding the PANDORA toolset to include machine-learning workflows.

**In summary**, PANDORA delivers a scalable, RAP-friendly framework that standardizes voxel/grayordinate access across 98 UKB sub-modalities, introduces information-preserving supervoxels, and implements a new supervoxel regression approach that operates in encoding space while returning full-resolution statistics. Benchmarks show major I/O, memory, and runtime gains; 10K-supervoxels closely match full-voxel results and can serve as a drop-in replacement, while 1K-supervoxels boost power for low-SNR effects. Across five experiments spanning exposure, neuropsychiatric, and genetic domains, we replicated known findings and uncovered new multimodal associations, demonstrating that PANDORA will accelerate discovery and maximize sensitivity.

## Online Methods

### Dataset

PANDORA UKBv1 encompasses brain MRI data for N=81,939 UKB subjects (53.2% female; mean age ± SD at study baseline: 55.0 ± 7.5 y, range: 40.1 – 70.4 y; mean age ± SD at first imaging session: 66.6 ± 7.9 y, range: 44.6 – 87.1 y), with 98 sub-modalities (distinct image processing output types) derived from 6 imaging modalities across structural, diffusion and functional MRI (supplementary table 1). Functional MRI images use the HCP CIFTI representation^46^ as grayordinates (cortical surface vertices and subcortical voxels), and all other modalities use voxel representation, in MNI152 standard space.

For each sub-modality, full-resolution data are collated into a large subject × voxels (or grayordinates) 2D matrix, with the voxels/grayordinates from a single subject unwrapped into a single row in the matrix. Additionally, for every modality we provide two compressed versions, i.e., subject × supervoxel matrices for 1K- and 10K-supervoxels. The supervoxel matrices come with supervoxel to voxel (/grayordinates) mappings for spatial interpretation.

To ensure high compatibility (Python, MATLAB, C/C++) and fast disk-access for analyses, all data files use the Hierarchical Data Format (HDF5) format^47^, which among other things allows for contiguous disk space allocation. Furthermore, full-resolution matrices and supervoxel-to-voxel mappings are oriented such that the larger voxel dimension lies along the faster-varying (stride-1) dimension, which makes streaming them during analysis a viable option.

As our data processing pipeline already removes most redundant datapoints (e.g., voxels outside of the brain), standard file compression of the full-resolution data matrix did not add a large benefit in storage reduction (<15%), and we thus opted to prioritize simplicity and data access speed, and not add file compression (while achieving disk space reduction using the supervoxel approach).

### Data processing

To create the full-resolution voxel-level datasets, pre-processed UKB NIfTI / CIFTI images as described in Alfaro-Almagro et al.^2^, were first loaded into a subject × voxel (or grayordinates) matrix; 3D NIfTI images (in standard MNI space) were either masked using a mean fractional anisotropy image-derived white matter mask (for dMRI-based sub-modalities) or using a liberal T1-derived whole-brain mask (for all other sub-modalities). The much smaller surface-based CIFTI images were loaded in completely, as there are no “background” grayordinates.

To remove low-variance voxels, we computed across-subject standard deviations for each voxel, discarding those with values < 5% of the 90^th^ percentile (robust maximum) across voxels. This variance-derived mask, a subset of the loading mask, is available for every PANDORA sub-modality.

Next, outlier participants (with respect to mean image intensity and/or number of outlier voxels) were fully removed, after which individual outlier voxels were imputed in the remaining subjects using the column median value. This process is described in detail below.

Finally, the columns of the remaining modality matrix are demeaned, and then the whole matrix is divided by its global standard deviation. Crucially, we do not standardize individual columns, as this would remove signal variance differences across voxels that may be important. Both column- and row-mean are saved (as mean modality image and mean subject-course) as part of this processing.

Due to the large memory requirement of holding the entire full-resolution matrix in RAM as required for encoding, processing steps had to be ensured to work with in-place operations, exclusively necessitating careful selection of library functions (e.g., NumPy’s median function copies its input, while nanmedian does not) and custom code for non-contiguous column/row removal. A compute node with 3TB RAM was required to create PANDORA for the largest of the sub-modalities.

All data processing was performed in Python 3.10.4 using NumPy and scikit-learn (versions 2.2.3 and 1.15.2, respectively).^48, 49^

### Modality-specific processing

T1, T2-FLAIR and voxel-based morphometry (VBM) maps required specific processing steps applied before loading images into the initial subject × voxel matrices.

Due to their non-quantitative nature, bias-field corrected T1 and T2-FLAIR images were normalized using the image median white matter value. This normalization will not always be perfect, e.g., in cases where biological signal of interest may bleed into the normalization factor. For example, for T2-FLAIR images, the median may be shifted in subjects with very high lesion load, and also partially correlates with age. Future work will attempt to address this with more sophisticated normalization.

Standard processing was applied for VBM maps. Specifically, we applied a smoothing with a Gaussian kernel with sigma 2.12 mm (5 mm full width at half maximum). Additionally, voxels with median gray matter probability < 5% were discarded after removal of low-variance voxels in the main pipeline (median probability per voxel was estimated using a random subset of 10,000 subjects).

### Outlier removal

While the UKB data has undergone extensive quality control as part of the core image processing pipeline, the data still contains some outlier samples, which were either biological outliers (large-scale anatomical/pathological anomalies) or caused by image artifacts. We developed a two-step outlier removal pipeline, primarily aimed at removing outliers of the latter kind with exception of very extreme cases (e.g., one sample with extensive tissue loss). This was followed by imputation of outlier voxels.

To find a good balance between removing subjects with image artifacts and not removing those with biological pathology (which some researchers may wish to study), thresholds had to be manually determined for each modality. This was done by visual inspection of representative numbers of samples at different cut-off values, comparing average valued samples to cases with image artifacts, as well as those showing plausible biological and pathological variation.

In step 1, we removed outliers with respect to mean image intensity (i.e., the row-wise mean of the full matrix). Similar to Alfaro-Almagro et al.^50^, we subtract the population median image intensity from the subject’s mean image intensities and calculate the median absolute deviation (MAD) of these values (i.e., the median of the absolute value of the mean intensities). The demedianed mean intensities are then scaled by MAD. For most modalities a reasonable outlier threshold could be set on the absolute value of the MAD-scaled intensities, whereas for a few (notably T1 and FA), using different thresholds on negative and positive outliers gave a better demarcation of outliers with image artifacts. The thresholds for this first step ranged between 6-12 MAD.

Step 2 was aimed at removing outlier subjects with a large number of extreme-valued voxels or grayordinates. Here, differently to step 1, we performed MAD scaling on the columns of the full matrix rather than on the row-wise means. Specifically, we first subtracted the column-wise median from each entry of the matrix and then estimated column-wise MAD by taking, for each column, the median of the absolute values of the demedianed entries. The full matrix was then scaled using these column-wise MAD values. Because MAD can be zero, values below a numerical precision threshold were replaced by the corresponding column standard deviation divided by 1.48 (using the approximation SD ≈ 1.48 × MAD) before scaling. To reduce computational cost, column-wise medians and MADs were estimated on a random subset of 10,000 subjects.

Using this MAD-scaled matrix, we defined, for each participant, the number of extreme-valued voxels as the number of voxels exceeding a threshold (fixed at 6 for most sub-modalities). Having obtained these subject-level counts, we could then perform the same procedure on them as for the subject-level mean intensities, i.e., apply another round of MAD-scaling and set a second, sub-modality specific, threshold, to these MAD-scaled counts, defining the outlier participants for step 2.

The first threshold of step 2 was set at a MAD-score of 6 for all sub-modalities except T1 and the Jacobian warp, where a higher initial threshold (MAD-score 20) resulted in a better separation of subjects with atrophy and subjects with warping errors. For the second threshold, optimal values per sub-modality varied more widely than for step 1, ranging from MAD-scores of 6 (e.g., for T2-FLAIR and SWI) up to 2000 for one of the task fMRI z-statistic maps.

In step 3, individual outlier voxels (or grayordinates) were imputed in the remaining subject matrix after outlier removal. That is, voxels above another sub-modality specific threshold in the previously demedianed and MAD-scaled matrix were set to 0, corresponding to median imputation. Manually determined thresholds for step 3 spanned a wide range, from MAD-scores of 8 (e.g., SWI T2^*^) up to 1000, with the overall aim of reducing noise- or artifact-related outliers without removing normal or pathological variation (e.g., large ventricles).

Step 1 was skipped for the Jacobian warp, where no satisfactory balance between sensitivity and specificity could be found. The outlier procedure was also not applied to the T2-FLAIR lesion map, as the binary lesion data rendered these steps unsuitable.

The manually determined thresholds are available within the PANDORA processing pipeline (https://git.fmrib.ox.ac.uk/PANDORA/pandora_proc).

### Supervoxel encoding

All subject × voxel (or grayordinates) full-resolution matrices were separately encoded into a subject × 1K-supervoxel matrix and a second, more detailed, subject × 10K-supervoxel matrix, using independent component analysis (ICA). One supervoxel is one ICA component—a soft clustering of voxels (Fig. S7) which have highly similar subject variability to each other. This has the advantage of giving an embedding with simple, focal spatial interpretability, and without losing spatial detail (e.g., because of pre-smoothing^51^). We used the FastICA algorithm of Hyvärinen and Oja ^52^, as implemented in scikit-learn^49^, with the scikit-learn’s randomized SVD function for whitening^53^.

Let *Y* ∈ ℝ^*N*×*V*^ denote the full subject × voxel matrix. Consider the SVD

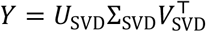

where *U*_*SVD*_ ∈ ℝ^*N*×*K*^ are subject eigenvectors, *V*_*SVD*_ ∈ ℝ^*V*×*K*^ are spatial eigenvectors, and Σ_*SVD*_ is diagonal containing the non-negative singular values. The number of eigenvectors is *K* < min(*N, V*).

ICA is then applied to the first *K* spatial eigenvectors, finding a *K* × *K* mixing matrix *A*_ICA_ and *K* statistically independent components collected in *S* ∈ ℝ^*K*×*V*^ such that

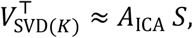

where the (*K*) subscript denotes selecting first *K* dimensions. These decompositions are combined into the *K*-dimensional approximation

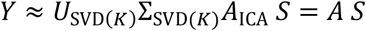

where the subject-weight matrix

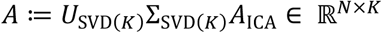

contains the subject weights for the supervoxels, and *S* ∈ ℝ^*K*×*V*^ contains the spatial maps (one supervoxel map per ICA component). Hence *A* can be used instead of the full voxel data *Y* for downstream analyses such as regressing imaging against other variables and is indeed the main variable used in PANDORA analyses.

*S* can be used to interpret between-subject analyses carried out using *A*, or (as below) utilized in efficient ways *within* analyses in order that they may become equivalent to voxelwise analyses, but without the high-memory requirements of working with the full data (and with denoising, controlled by *K*).

Note that the subject weights *A* can be connected back to the original data with the definition of *U*,

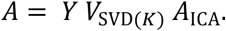

This shows that the subject weights are simply linear combinations of the voxelwise data *Y*. Therefore, if the rows of *Y* (i.e., subjects) are regarded as independent, then the rows of *A* can likewise be treated as independent.

### Supervoxel GLM

In order to efficiently utilize the supervoxels and importantly bypass the reconstruction of a costly (huge) full-resolution *N* × *V* matrix, we use an adjusted general linear model (GLM) regression approach.

### Setup

In addition to data *Y*, subject weights *A* ∈ ℝ^*N*×*K*^, and supervoxel maps *S* ∈ ℝ^*K*×*V*^, we have an *N* × *P* design matrix *X*, and either a contrast vector *c*^⊤^ ∈ ℝ^1×P^ or a contrast matrix *C* ∈ ℝ^M×P^. *X* can include variables of interest and confounds (PANDORA is supplied with a pre-estimated set of imaging confounds, and the user can optionally add others, such as genetic principal components). This is a challenging setting because there are up to millions of voxels (*V*) and 82K subjects (*N*).

While the original, full data mass-univariate model is

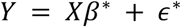

we are instead interested in a mass-univariate model of the ICA-reduced supervoxel data,

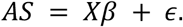

The core assumption of the mass-univariate linear model is independent errors (across subjects) for each voxel *i*. As noted above, the traditional assumption of independent subjects in *Y* likewise justifies the independence, over rows, for each column of data of *A*, and this independence is maintained after spatial projection by *S*. This justifies an assumption of independent errors *ϵ*.

### Retaining the mean in the denoised model

Most statistical analyses treat the overall mean or intercept as a nuisance, and it is also the case that the ICA decomposition is performed on centered data. However, the mean subject-course can be a feature of interest (e.g. in task fMRI), and thus we need to retain this information. Let 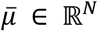 denote the mean subject-course (voxelwise mean across space),

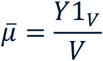

(with 1_V_ being the *V* × 1 column vector of ones) so that the centered ICA approximation satisfies

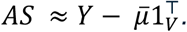

We integrate 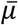 into the decomposition by augmenting

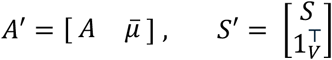

so that

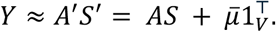

For simplicity, we drop the prime superscript below and treat *AS* as including the mean subject-course. Note that the mean *image* has also been subtracted from the data (at an even earlier processing step) but does not need to reappear for the purposes of ICA or regression. The mean image is, however, included in the PANDORA distribution, because it can be useful for later visualizations (for example, for visualizing the average image of a subset of subjects *A*_*subset*_*S*, the mean image should be added on).

### Model estimation

We estimate as Equation (1) with ordinary least squares:

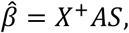

where *X*^+^ denotes the Moore-Penrose pseudoinverse, equal to (*X*^⊤^*X*)^-9^*X*^⊤^ when *X* is full rank. Define the reduced-space coefficients as

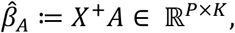

which combine the regression and subject weights so that the full voxelwise coefficient image can be written as

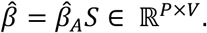

An essential insight is that while 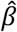 is a large *P* × *V* matrix, a sufficient statistic for the regression is 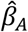, a much smaller *P* × *K* matrix. However, we do not want to make inference on the *K*-dimensional regression coefficients 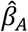, but in voxel space. Writing the *i*th voxel estimate as 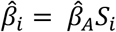, where *S*_;_ is the *i*th column of *S*, we now develop inference procedures in the voxel domain using 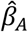.

### Sample variance of estimates

As mentioned above, the traditional assumption of independent subjects justifies the row-wise independence of each *ϵ*_;_. Thus, the sampling variance for the estimate at the *i*th voxel is

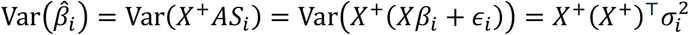

where 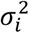 is the residual variance at voxel *i* under the denoised model.

### Contrast inference

For a contrast vector *c*^⊤^ ∈ ℝ^1×P^, we wish to test the null hypothesis *H*_0_: *c*^⊤^*β*_i_ = 0 for each voxel *i*. For this the contrast estimate is 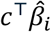 and the sampling variance

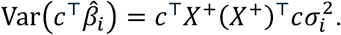

For a contrast matrix *C* ∈ ℝ^M×P^ we likewise test *H*_0_: *Cβ*_i_ = 0 with the sampling covariance of 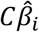

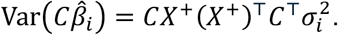

F*-Test*

For an *M* × *P* contrast matrix *C*, define the *M* × *M* matrix

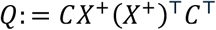

At voxel *i*, noting that 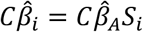, the F-statistic is

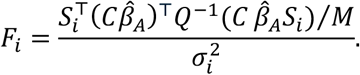

To compute this for all voxels simultaneously, consider the computation in two steps, first whitening (technically, overwhitening) the *M* × *V* matrix of contrast estimates

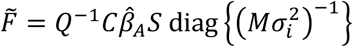

where postmultiplication by diag{·} specifies a column-wise scaling, and then the final 1 × *V* F-statistic image is the column-wise sum of the Hadamard product

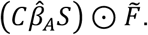

Of course, in practice, one can do this all together, generating the F-image as

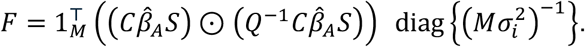

### Residual error calculation

For the voxel *i*, its residual error is

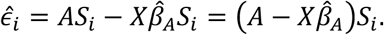

Naïve calculation of the sum of squared errors, 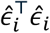 does not exploit the low-dimensional structure, nor is easy to vectorize. Instead, consider the *N* × *K* reduced-dimension residual matrix *R*

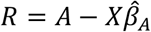

Then for voxel *i*,

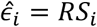

and the sum of squared errors can be written directly as

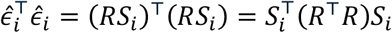

Let

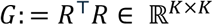

be the Gram (cross-product) matrix of the reduced residuals. Then

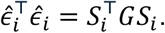

The residual variance at voxel *i* is

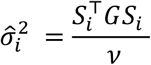

where *ν*: = *N* − rank(*X*) is the residual degrees of freedom (typically *ν* = *N* − *P* when *X* is full rank).

### Efficient evaluation

To make the computational benefit explicit, consider the voxel maps as a single matrix *S* = [*S*_1_, …, *S*_V_] ∈ ℝ^*K*×*V*^. The vector of voxelwise sums of squared errors is the diagonal of *S*^⊤^*GS*:

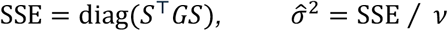

We do not compute the *V* × *V* matrix *S*^⊤^*GS*. Instead, we compute

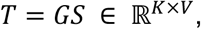

and then evaluate the diagonal using column-wise dot-products

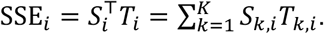

This yields an efficient, vectorized computation requiring only a *K* × *K* Gram matrix and matrix-matrix products in the reduced space. Forming *G* costs *O*(*NK*^2^) once, and the voxelwise step costs *O*(*K*^2^*V*) (plus *O*(*KV*) for the final dot-products), avoiding explicit computation of the voxel-space residuals *RS*, which would cost *O*(*NKV*) and require handling an *N* × *V* residual matrix.

### fsl_glm PANDORA expansion

The new supervoxel GLM approach for PANDORA data is accessible via an expansion of the *fsl_glm* tool, which is part of the MELODIC toolbox in FSL^54^. The software, written in C++, has been adapted to process HDF5 data in separate voxel/grayordinate and supervoxel pipelines. Both pipelines can be run in single- or multi-threaded mode. The pipelines have been optimized for memory efficiency and speed. In particular, the full-resolution pipeline reads data from disk in chunks along the (stride-1) voxel axis, while the supervoxel pipeline does the same for the component-to-voxel mappings (if needed). All of the supervoxel GLM operations described above are implemented in this chunked, streaming fashion on the spatial maps, which keeps memory usage effectively constant with respect to the number of voxels/components. Supervoxel analyses are currently available for 1K and 10K components. Using the modality mask from the processing stage, the final statistical maps are output as images (NIFTI or CIFTI, and hence more convenient for display than HDF5). Because of the high-N setting, we output p-values as -log_10_(*p*).

The expanded *fsl_glm* also provides new processing capability for CIFTI data. Moreover, the user can select one of two sets of pre-processed imaging confounds to include in the model.

Subject subsets of interest are selected simply by including only the relevant subjects in the user-supplied subject ID file, alongside the design matrix file (which must have matching subject rows). This allows for sex-specific analyses, discovery/replication splits, or cross-validation, without modifying the pipeline. The supervoxel (ICA) decomposition is pre-computed once per modality on the full cohort and then held fixed for all such analyses; the GLM stage (including confound modeling and normalization) is then estimated on the chosen subset. Because the ICA step is fully unsupervised and independent of outcomes or group labels, reusing this basis does not compromise cross-validation.

We benchmarked runtimes (along with diskspace use) for small, medium and large modalities using the voxel vs. supervoxel pipeline.

### Confound processing

PANDORA comes with two sets of preprocessed imaging confounds (*confounds_all* and *confounds_small*) available for the full sample. The full set is 222 confounds, reflecting many technical and subject variabilities such as head motion and head size, and the reduced set just covers the 20 most important confounds (sex, head size, etc.). The confounds were processed using a new pipeline described in detail in a recent preprint^55^ and following Alfaro-Almagro et al.^50^, which retains confounds that pass statistical tests of being relevant to the imaging data.

The small set of most important imaging confounds can be useful for example when running analyses on a small subset of UKB subjects. This set was chosen by keeping confounds for which the maximum absolute correlation across all IDPs was greater than 0.5, or where the median absolute correlation was greater than 0.04.

### Encoding quality

To quantify the information content retained by the 1K/10K-supervoxel encoding, and, in comparison, by the modality-specific and multimodal IDPs (i.e., pooling all IDPs from all modalities), we estimated percent of variance explained for each modality by the respective encoding. This is the amount of variance in the full data captured by the subspace spanned by its encoded representation.

Let *Y* ∈ ℝ^*N*×*V*^ denote the full-resolution matrix and *A* ∈ ℝ^*N*×*K*^ the encoded basis (supervoxels or IDPs). *Y* is projected onto the column space of *A* via its pseudoinverse:

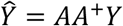

We then compute the proportion of variance explained as the ratio of the squared Frobenius norms:

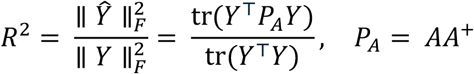

We report 100 × *R*^2^ as % variance explained.

To investigate the overlap between supervoxels and IDPs, we further computed 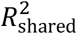 as the variance explained of *Y* projected onto the subspace spanned by IDPs by *Y* projected onto the span of the supervoxels times the full-resolution variance captured by the IDPs:

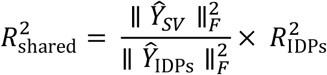

Because there are no modality-specific IDPs for warpfield Jacobian and tractography, we conservatively used all T1-derived IDPs and all diffusion-derived IDPs for the respective modalities.

### Benchmarking

We benchmarked the supervoxel regression against full-resolution analysis across five representative sub-modalities for a typical analysis with one regressor of interest and the full set of imaging confounds. Benchmarks were run in parallel, sweeping number of threads {4, 8, 16, 32, 64} and chunk sizes {1000, 2000, 4000, 8000}. Each configuration was repeated three times and summarized by the median. We recorded wall-clock (“real”) time, user CPU time (core-seconds; summed across threads), system CPU time, and peak resident memory. Cloud instance-like runtimes were derived from the same sweep by filtering configurations to those within the instances’ CPU core limit and discarding runs whose peak memory exceeded instance RAM.

### Applications of PANDORA

To assess the analytic potential of PANDORA and the supervoxel analysis pipeline, we performed a series of experiments of varying complexity, constructing regression design matrices from UKB variables and regressing against multimodal imaging data. All analyses were run with our expanded *fsl_glm*, which takes a given PANDORA sub-modality folder, together with simple text files specifying subject list, design matrix and regression contrasts as input, and can straightforwardly switch between full-resolution and supervoxel analyses. Pre-computed imaging confounds are by default automatically concatenated onto the input design within *fsl_glm*.

For single-variable designs, we supplied a text file containing the variable of interest and a matching subject list; *fsl_glm* then constructed the full design matrix internally by appending the selected imaging confounds and applying column-wise demeaning. Rows (subjects) with missing values in any design column are automatically removed. Contrasts for positive and negative main effects were specified as simple two-row text files (one row per contrast, one column per design matrix regressor). More complex designs (e.g., multiple behavioral or genetic regressors and subtractive terms) were handled in the same way, with multi-column design and contrast matrices. From the user’s perspective, the interface is identical for voxel and supervoxel analyses.

All main analyses used the full set of imaging confounds (confounds_all; see above) and demeaned designs. We first ran simple single-variable experiments (neuroticism score, Alzheimer’s disease polygenic risk score (AD PRS), and effect of smoking) to verify the behavior of the pipeline and to compare 1K/10K-supervoxel with full-resolution regression in a straightforward setting. The core results reported in this paper are based on five more complex experiments (1–4, S1), each designed to address a distinct scientific question: the impact of cumulative trauma, differential anxiety–depression effects, a specific *EPHA3* variant, early-versus late-diagnosis autism polygenic scores, and a supplementary experiment on multimodal signatures of common substances. Table 1 summarizes experimental characteristics, confounds and number of performed tests. In general, the number of subjects is determined by the amount of non-missing data in the non-imaging regressors.

Regressors of interest for the five experiments were constructed as follows:

Cumulative trauma was defined as a composite score from multiple adverse-life-event items (similar to Künzi et al.^56^). Specifically, the score was calculated as the mean (after mapping to common scale) of the following semi-quantitative items from the adverse life events section of the mental well-being online follow-up: Experienced a life-threatening injury or illness (ID: 29087), experienced a violent or sexual assault (ID: 29086), experienced the death of a close friend of family member due to suicide (ID: 29090), experienced the death of a spouse or partner (ID: 29089), felt hated by family member as a child (ID: 29078), physical violence by partner or ex-partner as an adult (ID: 29083), physically abused by family as a child (ID: 29077), sexual intercourse by partner or ex-partner without consent as an adult (ID: 29085), sexual interference by partner or ex-partner without consent as an adult (ID: 29084), sexually molested as a child (ID: 29079), and stopped from seeing friends or family by partner or ex-partner as an adult (ID: 29081). Townsend deprivation index (ID: 22189) was added as confound to the design.

Anxiety and depressive symptom load were estimated as the first principal component of anxiety and depressive symptoms averaged across two online follow up questionnaires.

Items for anxiety comprised, in descending order of loading on the first principal component: recent worrying too much about different things (loading = 0.46; IDs: 20520, 29060), recent trouble relaxing (loading = 0.44; IDs: 20515, 29061), recent inability to stop or control worrying (loading = 0.43; IDs: 20509, 29059), recent feelings of nervousness or anxiety (loading = 0.41; IDs: 20506, 29058), recent feelings of foreboding (loading = 0.32; IDs: 20512, 29064), recent easy annoyance or irritability (loading = 0.31; IDs: 20505, 29063), and recent restlessness (loading = 0.21; IDs: 20516, 29062).

Items for depression comprised, in descending order of loading: recent feelings of tiredness or low energy (loading = 0.56; IDs: 20519, 29005), recent lack of interest or pleasure in doing things (loading = 0.38; IDs: 20514, 29002), recent feelings of depression (loading = 0.37; IDs: 20510, 29003), recent feelings of inadequacy (loading = 0.36; IDs: 20507, 29007), recent poor appetite or overeating (loading = 0.36; IDs: 20511, 29006), recent trouble concentrating on things (loading = 0.34; IDs: 20508, 29008), recent changes in speed/amount of moving or speaking (loading = 0.12; IDs: 20518, 29009), and recent thoughts of suicide or self-harm (loading = 0.11; IDs: 20513, 29010).

The first principal component explained 70.0% and 58.3% of answers to anxiety and depression questions, respectively. Townsend deprivation index (ID: 22189) was added as a confound.

For the *EPHA3* experiment, the regressor of interest was the additive dosage of SNP rs987748, coded as the expected minor-allele count per subject, with the minor allele frequency (MAF) being 0.372 in the analyzed sample (N = 67,192). Dosages were extracted from the UK Biobank imputed genotype data. 40 genetic principal components (ID: 22009) were added as confounds to the design matrix. As in all analyses, the design was demeaned within *fsl_glm*, and subjects with missing genotype or genetic principal component values were excluded.

A recent age-of-diagnosis–stratified autism GWAS^3^ reported two partially separable common-variant components corresponding to earlier vs later autism diagnosis. Using the public summary statistics for these components, we constructed two polygenic scores ^57^ in UK Biobank by harmonizing alleles, performing linkage disequilibrium (LD) based clumping with PLINK^58^ (p_1_ = 0.001, r^2^=0.5, 250 kb) using the UK Biobank imputed genotype panel as the LD reference; and summing dosage x β across clumped SNPs. The clumped SNP sets for the two scores did not overlap, and the resulting scores were (as expected) uncorrelated. Unlike typical polygenic risk score workflows, we did not tune clumping parameters or the p-value threshold, since our goal was to compare the general effects of predefined early vs. late factors rather than optimize prediction. The two scores were standardized and used as regressors of interest along with 40 genetic principal components (ID: 22009) as confounds.

Prior to clumping, the set of SNPs was restricted to those with minor allele frequency greater than 0.001, and Hardy-Weinberg equilibrium p-value less than 1E-7. Imputed SNPs with INFO score < 0.3 were excluded, leading to 20,329,180 SNPs retained. The 67,192 participants included in this experiment represent a maximal unrelated subset of all UK Biobank participants with scans available in Summer 2025, with recent white British ancestry (as determined by the UK Biobank variable *in*.*white*.*British*.*ancestry*.*subset*), with euploidy, and with no missing confounds. A random two-thirds subset was used for clumping; associations were assessed in the full sample (N = 67,192).

For smoking, coffee and alcohol we used items from the lifestyle and environment questionnaire, i.e., past tobacco smoking (ID: 1249, semi-quantitative: never smoked, tried once or twice, smoked occasionally and smoked on most or all days), coffee intake (ID: 1498, quantitative: cups per day), and alcohol intake frequency (ID: 1558, semi-quantitative: never, special occasions only, one to three times a month, once or twice a week, three or four times a week, daily or almost daily). As for all analyses, the design matrix was demeaned and standardized within *fsl_glm*. Maximum level of education obtained from the qualifications item (ID: 6138) in the sociodemographics touchscreen questionnaire, and Townsend deprivation index (ID: 22189), were added as confounds.

Across the five experiments, we selected a limited set of imaging sub-modalities *a priori* based on biological relevance to the target phenotype and to provide complementary coverage across structural, diffusion, and functional measures. We avoided testing an exhaustive set of sub-modalities to keep the multimodal results interpretable and to limit an overly large multiple-testing burden, particularly for weaker effects. In the autism polygenic-score analysis specifically, we did not extend testing to resting-state network (RSN) sub-modalities after weak task-fMRI effects in the primary 10K-supervoxel analysis, in order to limit additional multiple comparisons.

### Regression experiment statistics

As in Gillis et al.^59^, we corrected for multiple comparisons using the hierarchical false-discovery rate (FDR) approach described by Benjamini and Bogomolov ^60^.

While classical FDR correction^61^ controls for the expected false discovery rate in a flat family of hypotheses, it does not account for data-driven selection across levels of a hierarchy. For example, if map-wise FDR was used for 100 sub-modalities and there were no true effects anywhere, on average 5 sub-modalities would have 1 or more detections; considering just those 5 significant maps the average FDR within these maps would be 100% (i.e., all reported post-threshold voxels are actually null). The hierarchical FDR approach accounts for this selection and ensures average FDR control on the selected maps (in both fully null and non-null scenarios).

We used hierarchical FDR over the three levels of sub-modalities, contrasts and voxels, following a bottom-up then top-down scheme. For reference, the standard Benjamini Hochberg FDR method is as follows. Writing *N*_test_ ordered p-values 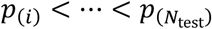, the level *α*_FDR_ FDR p-value threshold is *p*_(*i*∗)_, where *i*^*^ is the largest index *i* such that 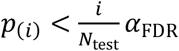; the FDR adjusted p-values are based on 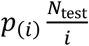, but capped at 1 and transformed to ensure monotonicity: 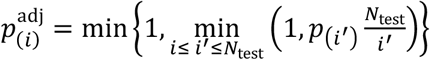.

#### Bottom-up

For each sub-modality *k* = 1, …, *K*, for each contrast *l* = 1, …, *L*, we computed “single map” FDR-adjusted *p-*values within the contrast (i.e., at every voxel/grayordinate). We summarized each contrast as the minimum FDR-adjusted *p-*value, and submitted these resulting *L* contrast summaries to a second FDR correction to obtain across-contrasts adjusted *p-*values (within each sub-modality), We then summarized each sub-modality by its minimum across-contrasts value, and submitted the resulting *K* sub-modality summary p-values to obtain *K* sub-modality-level FDR-adjusted *p-*values.

#### Top-down

We compared the *K* modality-level FDR-adjusted *p-*values to 0.05, and hence identified *K*_hit_ significant modalities. Within each of these *K*_hit_ sub-modalities, a further adjustment to the *L* FDR-adjusted p-values is made by scaling by *K* / *K*_hit_, and thresholding at 0.05 yielding *L*_*k*, hit_ significant contrasts in sub-modality *k*. Within each of the ∑_*k*_ *L*_*k*, hit_ maps, the final adjustment is made by scaling the single map FDR-adjusted *p*-values by (*KL*)/(∑_*k*_ *L*_*k*, hit_), which are then thresholded at 0.05.

### Anatomical description of experimental results

Anatomical localization for all experiments was primarily performed using the fourth edition of *Atlas of the human brain*^62^. Prefrontal cortex (PFC) definitions follow Carlén^63^, while thalamic subregions are reported with reference to a voxelized version of the Morel atlas^64-66^. For white matter tracts, the XTRACT HCP Probabilistic Tract Atlas^67^ and the JHU White-Matter Atlas^68^ were used for cross-checking.

## Supporting information

Supplementary Results and Analyses

Peak Statistics

Full Benchmarks

## Data Availability

All population-level summary statistic maps for all tests and all sub-modalities (z-statistics, regression coefficients and -log10(p) images) are openly available from: https://zenodo.org/records/19004272; PANDORA modality folders can be accessed (by all researchers approved for this data access by UK Biobank) on the UKB RAP cloud platform using the procedure described here: https://open.oxcin.ox.ac.uk/pages/pandora/web/; Code for the PANDORA processing pipeline is available at: https://git.fmrib.ox.ac.uk/PANDORA/pandora_proc; The fsl_glm source code is available within the MELODIC package: https://git.fmrib.ox.ac.uk/fsl/melodic; We have created a Docker image that is easy to get running inside a RAP compute node, and which includes: a graphical desktop (noVNC), all FSL brain image analysis tools (including fsl_glm), FSLeyes and HCP Workbench for visualization, Matlab (user must provide their own license), R and RStudio. The Docker image takes less than 10 minutes to install on a modest RAP instance and immediately allows to run analyses and visualisation of (e.g.) brain imaging data. Instructions are available at: https://docs.google.com/document/d/1QC3IaACVHS4MJQF1J71L_fNHLR4OkKJCB-Cs6FBEzGY.

## Ethics approval

The UK Biobank study was approved by the North West Multi-centre Research Ethics Committee (reference 11/NW/0382). The present project was authorized by UK Biobank under application ID 8107. All procedures were conducted in accordance with the principles of the Declaration of Helsinki.

## Data and Code Availability

- All population-level summary statistic maps for all tests and all sub-modalities (z-statistics, regression coefficients and –log10(*p*) images) are openly available from: https://zenodo.org/records/19004272.
- PANDORA modality folders can be accessed (by all researchers approved for this data access by UK Biobank) on the UKB RAP cloud platform using the procedure described here: https://open.oxcin.ox.ac.uk/pages/pandora/web/.
- Code for the PANDORA processing pipeline is available at: https://git.fmrib.ox.ac.uk/PANDORA/pandora_proc.
- The *fsl_glm* source code is available within the MELODIC package: https://git.fmrib.ox.ac.uk/fsl/melodic.
- We have created a Docker image that is easy to get running inside a RAP compute node, and which includes: a graphical desktop (noVNC); all FSL brain image analysis tools (including fsl_glm); FSLeyes and HCP Workbench for visualization; Matlab (user must provide their own license); R and RStudio. The Docker image takes less than 10 minutes to install on a modest RAP instance and immediately allows to run analyses and visualisation of (e.g.) brain imaging data. Instructions are available at: https://docs.google.com/document/d/1QC3IaACVHS4MJQF1J71L_fNHLR4OkKJCB-Cs6FBEzGY.

## Author Contributions

Conceptualization: A.A., K.L.M, S.J., M.W.W., W.G., C.F.B., T.E.N., S.M.S.

Methodology: all authors

Software: A.A. and M.W. (core implementation); F.A.-A., L.R., P.M., C.F.B., S.M.S.

Formal analysis: A.A., S.M.S.

Investigation: A.A., S.M.S.

Data curation: A.A., P.M., S.M.S.

Writing – original draft: A.A., T.E.N., S.M.S.

Writing – review & editing: all authors

## Supplementary Material

Full experimental results and discussion.

Full benchmarks.

Peak evidence tables for experiments.

