## Supplementary Results and Analyses for "PANDORA: Population Archive of Neuroimaging Data Organized for Rapid Analysis"

**Supplementary Results for GLM Experiments 1 – 4 and S1**

### **Experiment 1:** Association of lifetime trauma with the brain

Cumulative lifetime trauma scores corrected for deprivation score showed significant effects in twelve of thirteen tested structural and functional modalities (figures S1.1 and S1.2). Results are reported from the 10K-supervoxel analysis, except where otherwise noted. We classify evidence as strong for associations surviving hierarchical FDR correction at $p\_hFDR < 0.001$, and as moderate $p\_hFDR < 0.01$; the overall significance threshold is $p\_hFDR < 0.05$. Peak statistics for all experiments are summarized in supplementary tables.

Overall calibration and the relative sensitivity of the voxelwise, 10K- and 1K-supervoxel pipelines are summarized in S1.3, which contrasts observed −log10(p) distributions against permutation-derived and uniform-null expectations across sub-modalities. Below we summarize the main spatial patterns of association for each modality.

Anatomical localization for all experiments was primarily performed using the fourth edition of *Atlas of the human brain* (Mai et al., 2015). Prefrontal cortex (PFC) definitions follow Carlén (2017), while thalamic subregions are reported with reference to a voxelized version of the Morel atlas (Jakab et al., 2012; Krauth et al., 2010; Morel, 2007). For white matter tracts, the XTRACT HCP Probabilistic Tract Atlas (Warrington et al., 2020) and the JHU White-Matter Atlas (Hua et al., 2008) were used for cross-checking.

**Voxel-based morphometry (VBM):** Trauma was positively correlated with increased grey matter in right visual cortex (V1) and left ventromedial prefrontal cortex (vmPFC) with moderate to strong evidence. Moderate evidence was found for left V1, right vm- and dorsolateral (dl)PFC, across orbitofrontal cortices, left posterior fusiform gyrus, and in right paracentral lobule.

Strong evidence for grey matter decrease associated with trauma score was found for bilateral amygdalo-hippocampal complex, right substantia innominata, nucleus accumbens and left thalamus (VLp/VLa), and right calcarine sulcus. Moderate evidence was found for decrease in bilateral parietal operculum.

**Susceptibility weighted imaging (SWI) – quantitative susceptibility mapping (QSM):** Increase in QSM was seen with strong evidence in bilateral substantia nigra, dorsal striatum, amygdalo-striatal transition area, left medial forebrain bundle and in the right entorhinal cortex. QSM was decreased with strong evidence in lateral geniculate nuclei (LGN) with adjacent white matter, left ventrolateral thalamus (VLpv), right lateral hypothalamic area / substantia innominata, internal and external capsule white matter (extending to frontal operculum), temporal cortices (entorhinal, inferior temporal gyrus), and cerebellum.

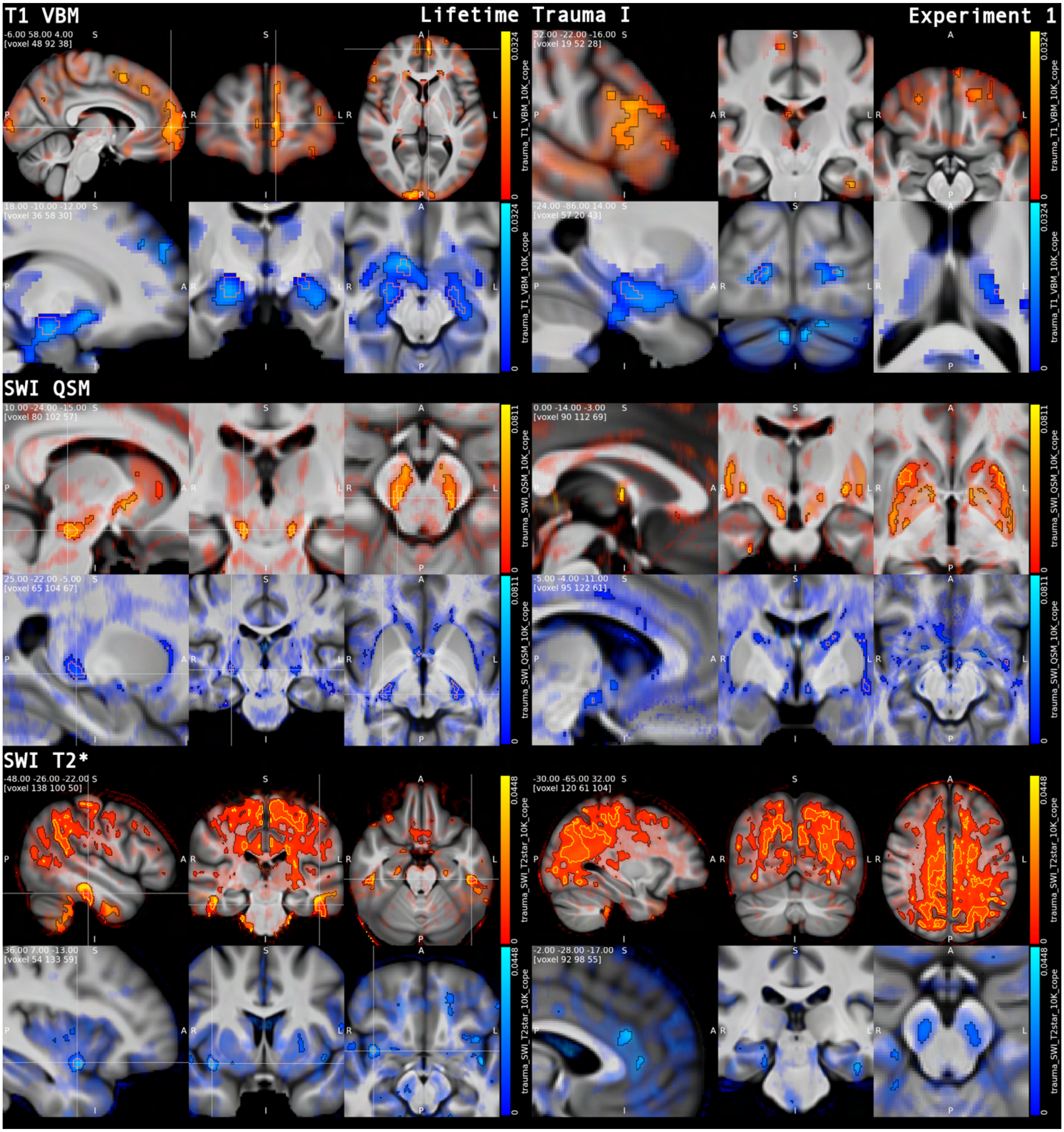


***Figure S1.1. Multimodal associations with cumulative trauma I: voxel-based morphometry (VBM) and susceptibility weighted imaging (SWI).*** *Life-time traumatic events showed distinct and distributed effects across modalities. Grey matter volume (VBM) was increased in prefrontal and visual cortices and decreased in the amygdalo-hippocampal complex (peak in right dorsocaudal amygdala: Z = -7.1; right amygdala IDP (UKB ID: 25889) for comparison: Z = -4.2). Quantitative susceptibility mapping (QSM) exhibited signs of iron accumulation in dopaminergic circuitry, supported by converging T2* effects in substantia nigra, while T2* further revealed widespread alterations in parieto-occipital white matter. <u>Common visualization conventions (Figs. S1.1-1.2)</u>: β-maps thresholded at hierarchical FDR corrected p < 0.05 (black outline) overlaid on non-significant effects modulated by Z-statistic (transparency without modulation for surface views), over mean T1-weighted image. Positive associations in red with yellow outline demarcating p_hFDR < 0.001, negative associations in blue with orange outline for p_hFDR < 0.001.*

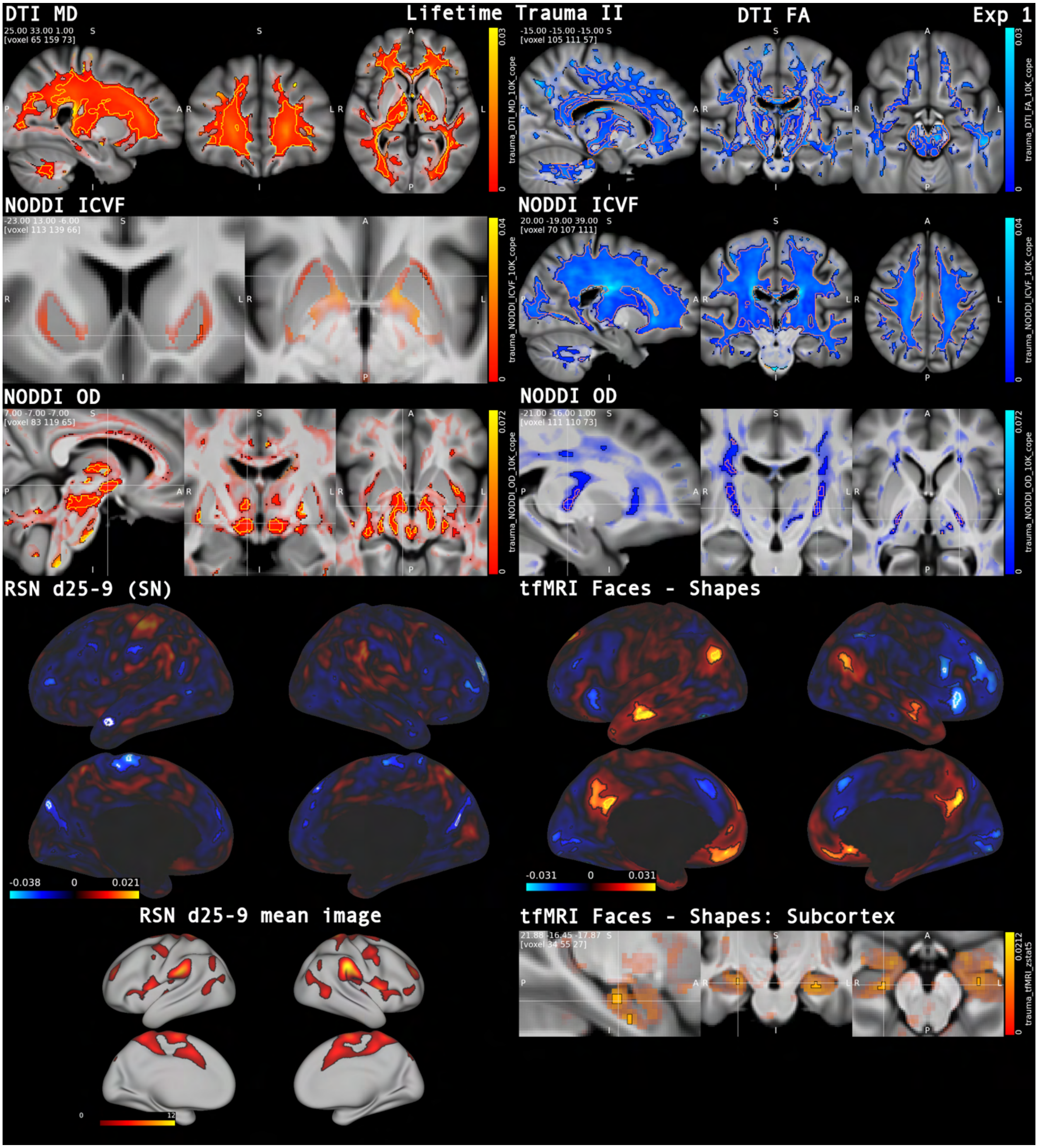


***Figure S1.2. Multimodal associations with cumulative trauma II: diffusion tensor imaging (DTI), neurite orientation dispersion and density imaging (NODDI) and functional MRI.*** *Global shifts in DTI- and NODDI-metrics (MD↑, FA/ICVF ↓) point towards reduced white matter integrity with higher load of traumatic events. Peak positive association with orientation dispersion (OD) was found in hypothalamus (which also exhibited a decrease in QSM) Functionally, trauma load was associated with increased activity of areas resembling the default mode network Hariri et al. (2002). (Visualization conventions as in Fig. S1.1).*

**SWI T2*:** Trauma exhibited widespread, strong evidence, positive associations with cerebral white matter in the parieto-occipital lobe, cingulum and along bilateral dorsomedial (dm)PFC. Peak evidence was found at the grey matter/white matter boundary of the left fusiform gyrus. T2* effects near middle fusiform gyrus should, however, be interpreted with caution because of possible susceptibility artifact from nearby mastoid air cells. Negative associations were strongly supported for right anterior insula, left superior temporal gyrus and bilateral cerebellum.

**Neurite orientation and density imaging (NODDI) – intracellular volume fraction (ICVF):** Neurite density related marker ICVF was positively associated in left putamen only (moderate evidence), while strong evidence for a negative shift in ICVF signal was seen across the entire supra- and infratentorial white matter with a robust peak in the right centrum semiovale ($-\log_{10}(p)$_mapFDR > 15).

**NODDI orientation dispersion (OD):** We observed strongest support ($-\log_{10}(p)$_mapFDR > 10) for increased OD in right hypothalamus and white matter between lateral substantia nigra and red nuclei. Further areas of strong evidence were left hypothalamus, bilateral thalamus, general substantia nigra area, and white-matter tracts along bilateral striatum and distinct cortical regions such as bilateral entorhinal and cingulate cortex.

Strongly supported negative associations were most prominent along bilateral corticospinal tract.

**DTI mean diffusivity (MD):** Evidence was strong for a global positive shift of MD, markedly, in fronto-parietal white matter with peak in the right anterior corona radiata. There were no negative associations with MD.

**DTI fractional anisotropy (FA):** FA was reduced throughout major supra- and infratentorial tracts with robust evidence for the left cerebral peduncle ($-\log_{10}(p)$_mapFDR > 15). There were no significant positive associations with FA.

**Task functional MRI (tfMRI) z-statistic (zstat) (effect of negative faces):** The main effect of negative faces from the emotion task showed moderate evidence for a positive association with right precuneus, i.e., reduced deactivation with respect to the group-level mean response, and significant negative association with right supramarginal gyrus, i.e., increased deactivation.

**tfMRI zstat (negative faces − neutral shapes):** For the faces−shapes contrast, we saw strong evidence for a positive association with left middle temporal gyrus, and the default mode network (specifically, bilateral precuneus, medial (m)PFC and left angular gyrus with significant activation of anterior hippocampus). Strong evidence for negative association was seen with right ventrolateral (vl)- and dlPFC, right anterior insula and angular gyrus.

**RSN d25-2 (DMN):** Within RSN d25-2 (DMN), evidence supported reduced coupling of dmPFC and left precuneus with higher trauma load, with no significant positive associations.

**RSN d25-9 (SN):** In the salience-like RSN d25-9 (SN) component, there was strong evidence for negative associations in left paracentral lobule, bilateral cuneus, and right dmPFC (reduced coupling), and left MTG (increased anticorrelation).

**RSN d25-13 (SMN):** Moderate evidence was found for a negative association in the sensorimotor network in the right precentral/opercular region (increased anticorrelation relative to group-level image).

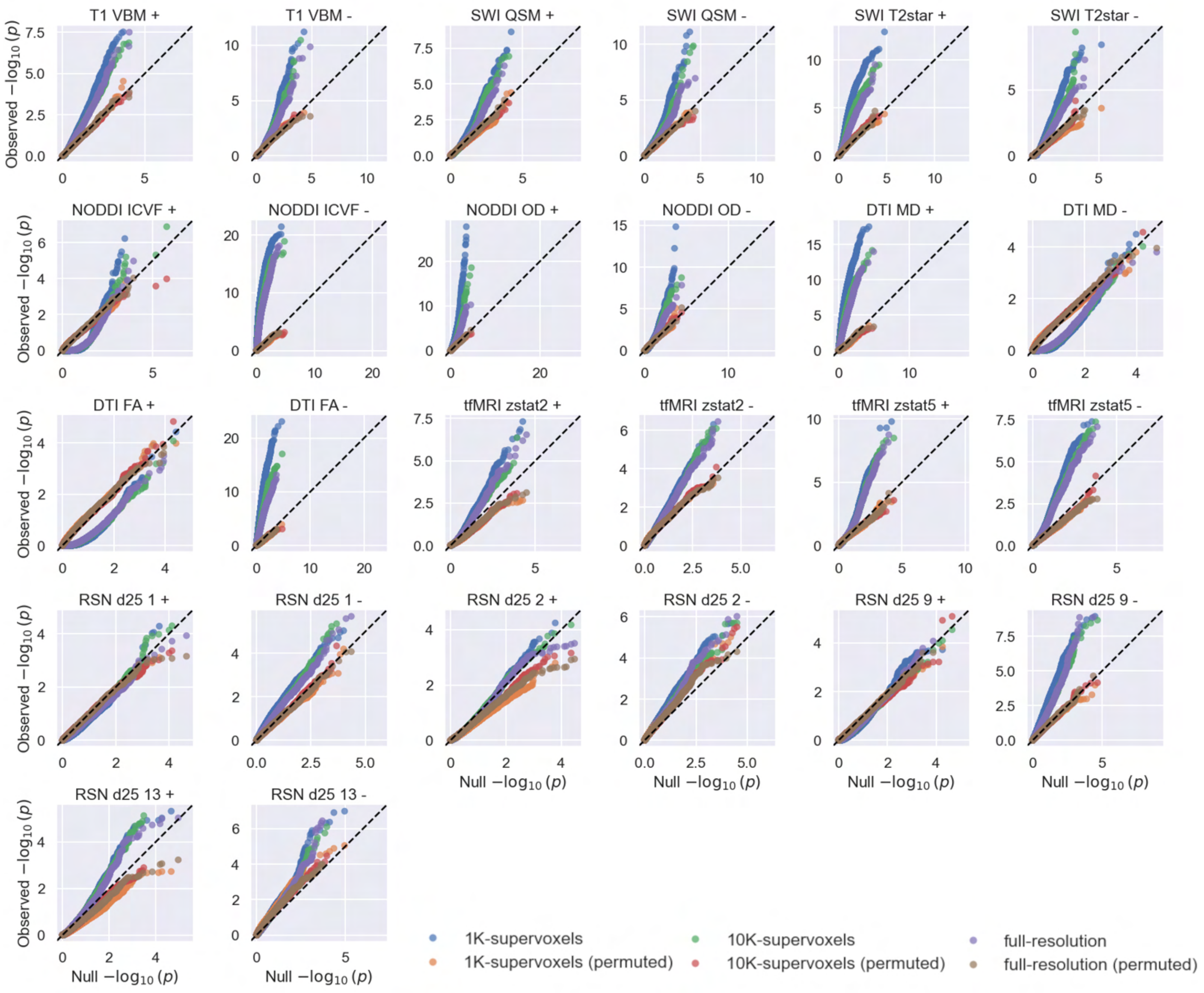


***Figure S.1.3. Observed vs. permuted vs. expected -log$_{10}$(p) distributions for the lifetime cumulative trauma multimodal analysis.*** *For each sub-modality, contrast and pipeline (1K-, 10K-supervoxels and full-resolution), we compare the empirical voxel/grayordinate-wise association p-values (observed) to a null distribution generated by permutation of the trauma regressor (permuted), and to the uniform-null expectation (expected; black dashed line). Upward deviation of the observed curves from expected indicates excess signal beyond chance, while agreement between permuted and expected indicates good null calibration. Significant effects at p_hFDR < 0.05 were detected in each sub-modality except for RSN d25-1 (visual network).*

### **Experiment 2:** Disentangling anxiety and depressive symptoms in the brain

Anxiety and depressive symptom principal components exhibited distinct effects across all 12 tested modalities (Figs. S2.1–2.6). In the following, we summarize the main associations and contrasts for these two symptom dimensions. We also revisited prior UK Biobank reports of absent Hariri-task amygdala associations (Tamm et al., 2022) by re-testing amygdala voxel-level reactivity using anxiety- and depression-specific symptom components, rather than using region-of-interest / IDP summaries (tfMRI zstat 1 – 5). Results are reported from the 10K-supervoxel analysis, except where otherwise noted.

**VBM:** Moderate evidence was found for a positive correlation of anxiety with the right cerebellum (VIIb), while moderate-to-strong evidence was found for positive changes associated with depression in bilateral putamen, left dlPFC, right frontal operculum and anterior insula, optic chiasm/hypothalamic region, posterior fornix/corpus callosum, left anterior intra-parietal sulcus, left visual cortex (V1), and bilateral cerebellum.

Moderate-to-strong evidence for negative associations of anxiety with grey matter volume were found along vm-/dmPFC (midline) and bilateral cingulate sulcus. Strong evidence for widespread grey matter reduction was found for depression. This included bilateral amygdalo-hippocampal complex, bilateral straight gyrus, subgenual anterior cingulate cortex (ACC), posterior insula/parietal operculum, and bilateral pulvinar.

**SWI QSM:** QSM was not significantly associated with anxiety symptoms. Moderate-to-strong evidence was, however, found for increase in the bilateral posterior insula, Heschl’s gyrus and cingulate sulcus for depression. QSM was negatively associated with the left bed nucleus of the stria terminalis (BNST)/anterior commissure.

**SWI $T2^*$:** Anxiety showed strong evidence for elevated $T2^*$ signal in bilateral dlPFC (peak in left hemisphere) extending into the grey matter/cerebrospinal fluid (CSF). No negative associations were found.

Depression associated with increased $T2^*$ in the subcallosal area, bilateral posterior inferior temporal gyrus and along right parieto-occipital white matter and cortex.

Depression scores were negatively associated with $T2^*$ across widespread cortical regions and along sulci (peak left dlPFC: $-\log(p)_{mapFDR} > 10$). Moreover, strong evidence for negative association with T2* was seen in anterior hippocampus.

We note that edge effects at brain/CSF boundaries should be interpreted with caution.

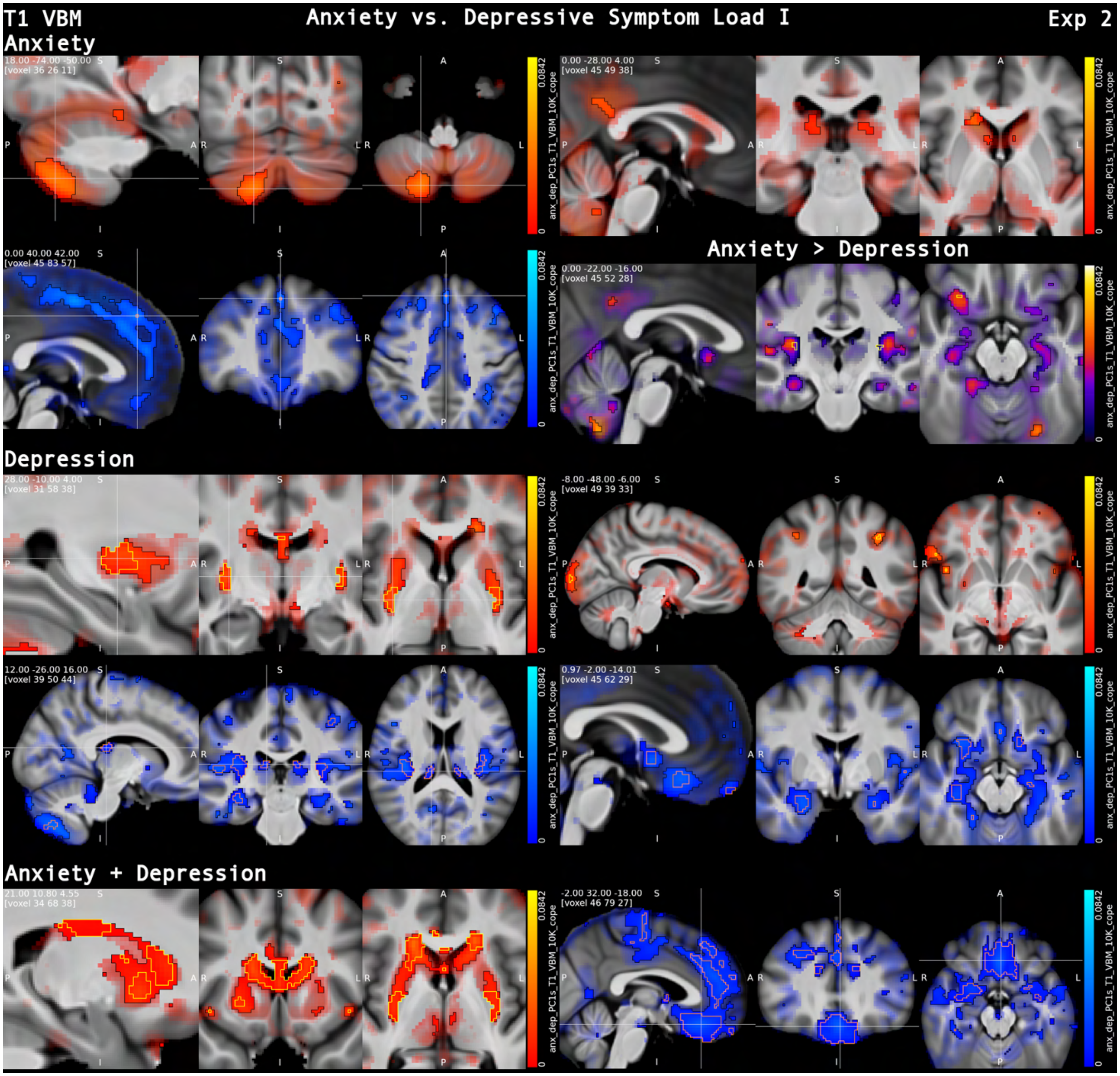


***Figure S2.1. Anxiety and depressive symptom load: voxel-based morphometry (VBM).*** *Anxiety and depression exhibited widespread volumetric brain changes such as volume loss across amygdala and hippocampus and increase in putamen for depression but volume increase in anterior caudate and posterior cingulate cortex (PCC) for anxiety. Volume loss for anxiety was focused on medial prefrontal cortex, while the increase in PCC was stronger for anxiety and decrease in subgenual anterior cingulate cortex (sgACC) and was stronger for depressive symptom load. Combined effects are displayed in the last row. <u>Common visualization conventions (Figs. S2.1–2.6)</u>: β-maps thresholded at p_hFDR < 0.05 (black outline) overlaid on non-significant effects modulated by Z-statistic (surface views in S2.4. and S.2.6.: non-significant effects shown by transparency, without Z-modulation), over the mean T1-weighted image. Positive associations are shown in red and negative associations in blue, contrast associations are displayed with a heatmap (NIH-fire); p_hFDR < 0.001 is outlined in yellow (positive and contrasts) or orange (negative), except on surface renderings, where p_hFDR < 0.001 is indicated in white. Group-mean resting-state network maps (Z-statistics) are thresholded at Z > 2.58 (approx. p < 0.01, two-tailed).*

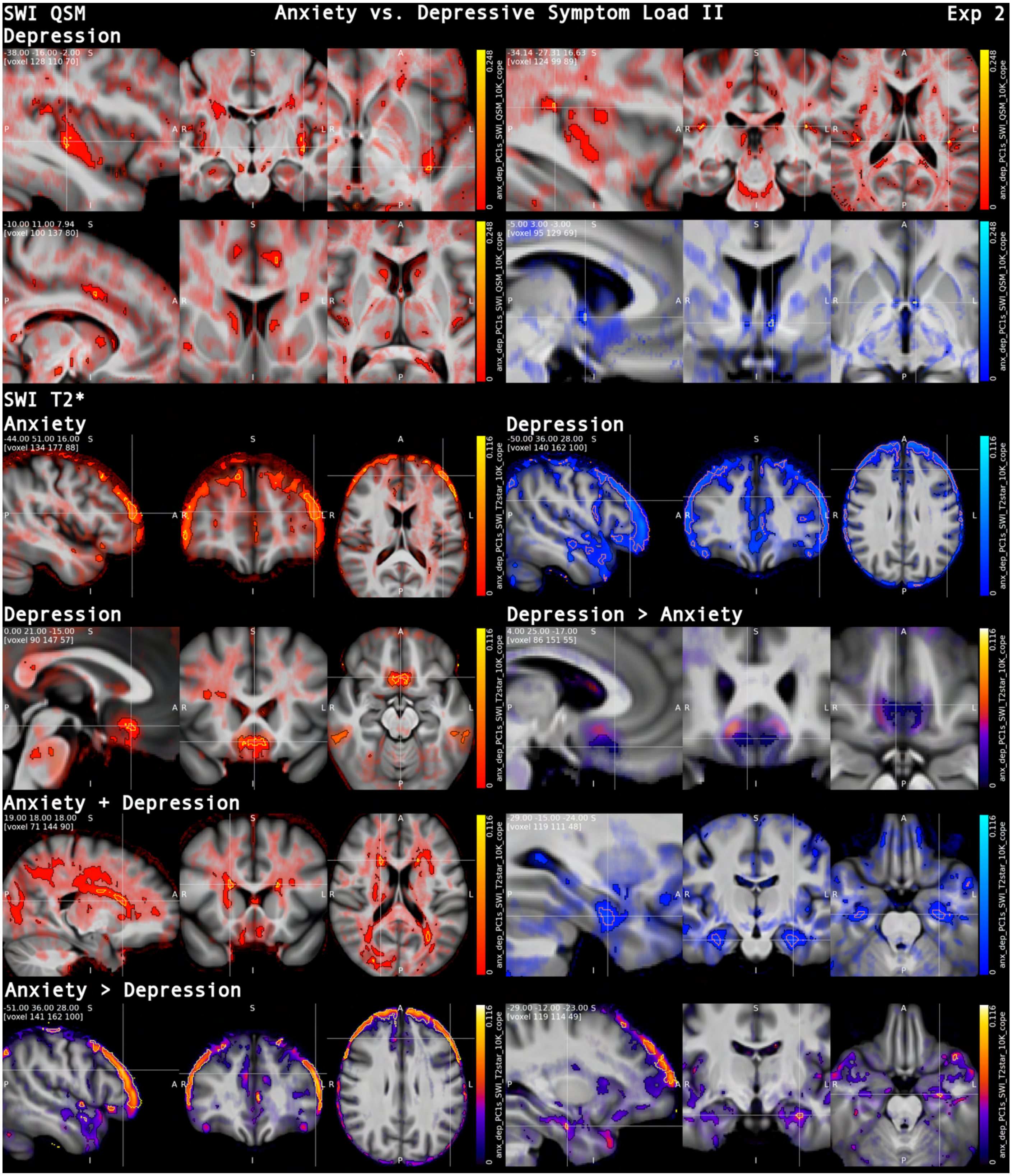


***Figure S2.2. Anxiety and depressive symptom load: SWI.*** *Depressive symptoms were associated (p_hFDR < 0.001) with higher QSM in posterior insula, left Heschl's gyrus and cingulate sulcus consistent with iron accumulation. T2* revealed a dissociated association with depression (↓) and anxiety (↑) within frontal lobe cortex (peak: dlPFC). The T2* decrease in depression, co-localized with QSM increase consistent with increased paramagnetic susceptibility (e.g., iron/venous effects). Surface-adjacent effects should, however, be interpreted with caution (e.g., partial volume, susceptibility artifacts or residual misalignment). (See figure S2.1. for visualization conventions.)*

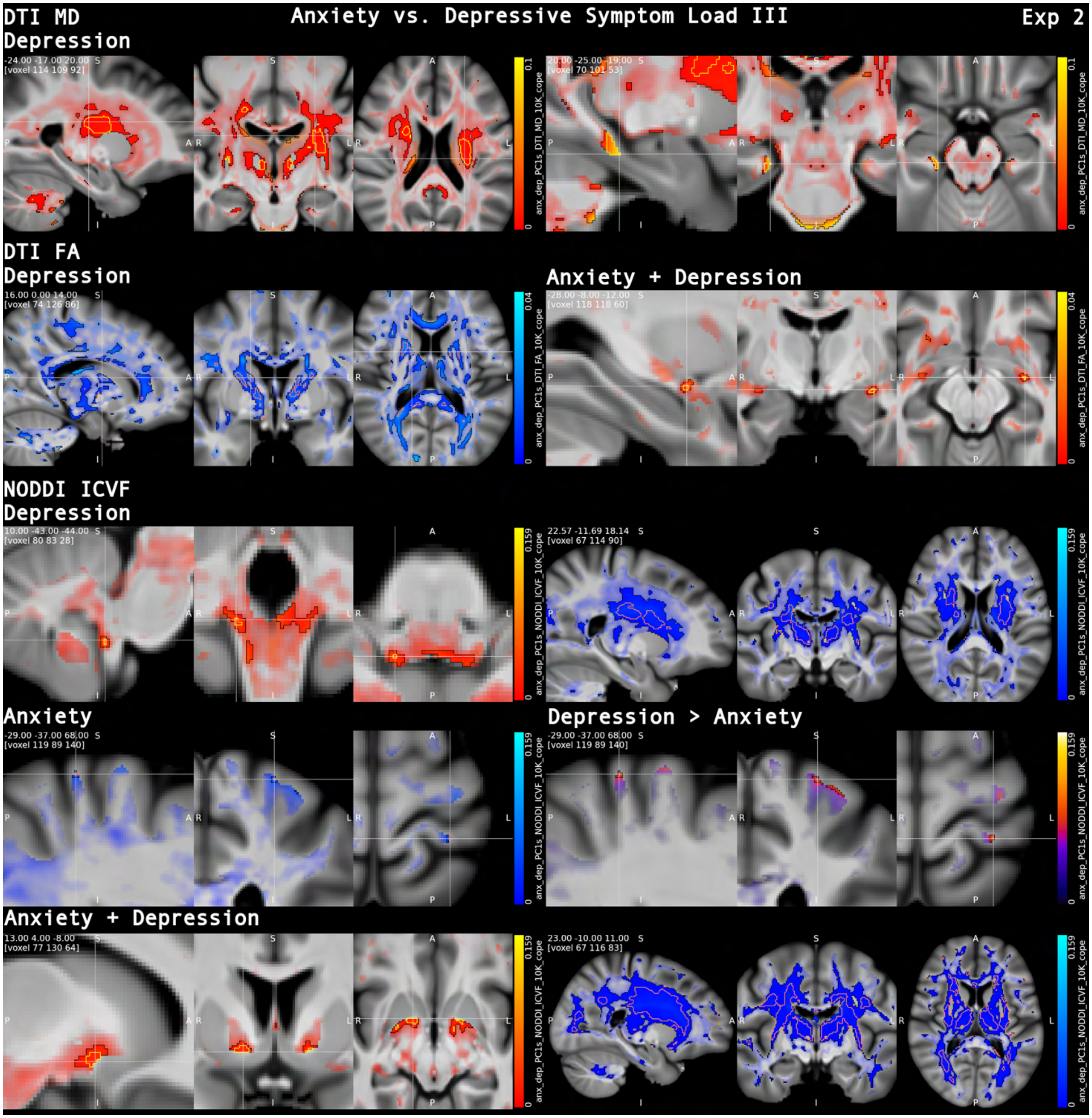


***Figure S2.3. Anxiety and depressive symptom load: MD, FA and ICVF.*** *Depressive symptoms were associated with a widespread pattern of increased mean diffusivity (MD) and decreased fractional anisotropy (FA) and neurite density (ICVF) across cortico-striatal, thalamic and fronto-parietal white matter, consistent with reduced microstructural integrity. In contrast, anxiety-related effects were more spatially circumscribed, including a focal decrease in ICVF at the WM/GM boundary of postcentral cortex, highlighting more localized microstructural alterations. Additive effects of anxiety and depressive symptoms were found in increased FA in anterior commissure adjacent to amygdala and increased ICVF in white matter superior to the substantia innominata. (See figure S2.1. for visualization conventions.)*

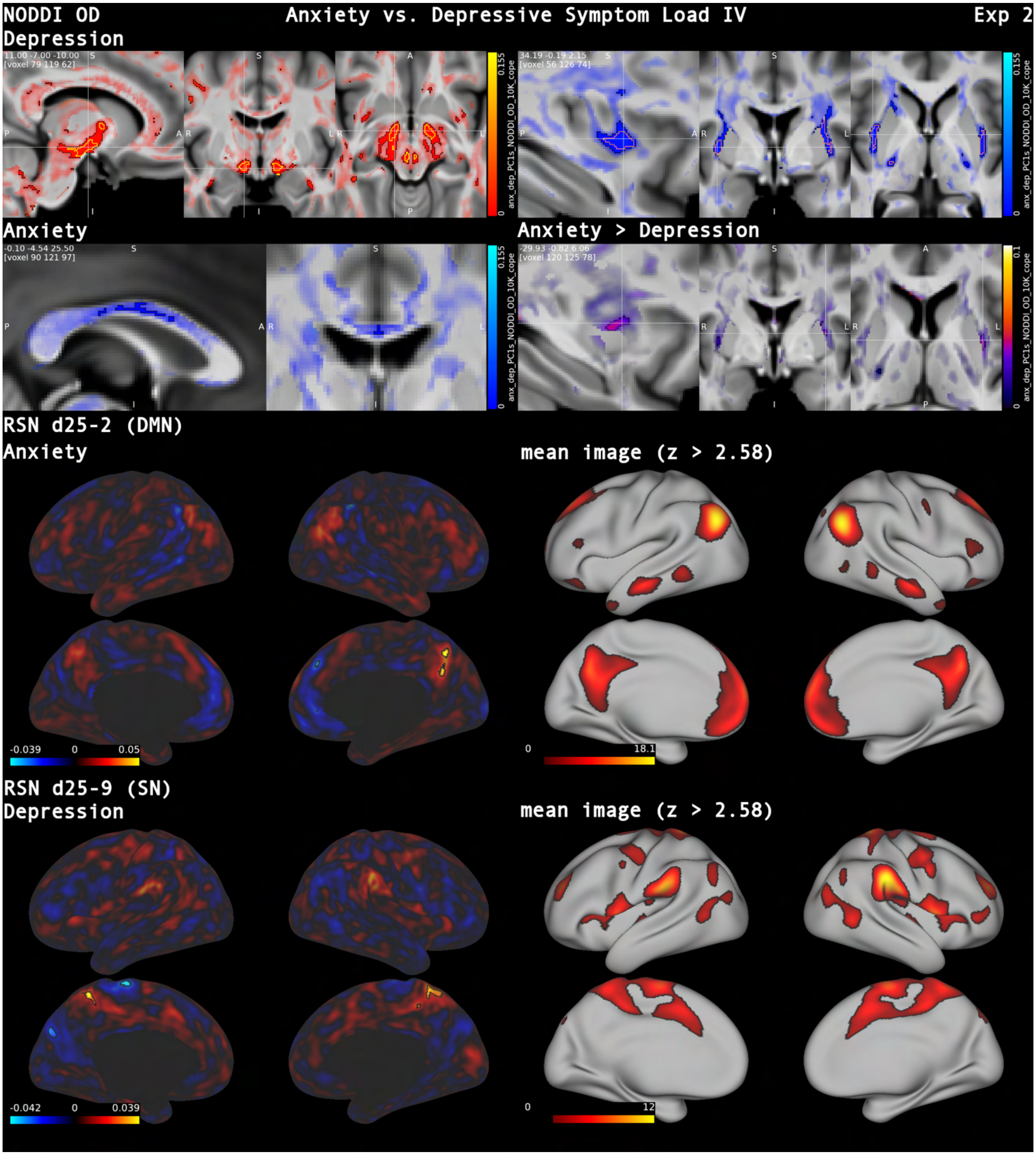


***Figure S2.4. Anxiety and depressive symptom load: OD and RSN.*** *Orientation dispersion (OD) was increased in bilateral hypothalamus for depressive symptoms, consistent with microstructural reorganization, and decreased in putaminal white matter, while anxiety showed additional OD reductions in callosal white matter. Resting-state analyses further revealed that anxiety symptom load was associated with increased coupling of posterior cingulate/precuneus and increased anticorrelation of dmPFC within the default mode network (RSN d25-2), whereas depressive symptoms were linked to increased precuneus coupling and reduced coupling of the left paracentral lobule within a salience-like network (RSN d25-9)**.** (See figure S2.1. for visualization conventions.)*

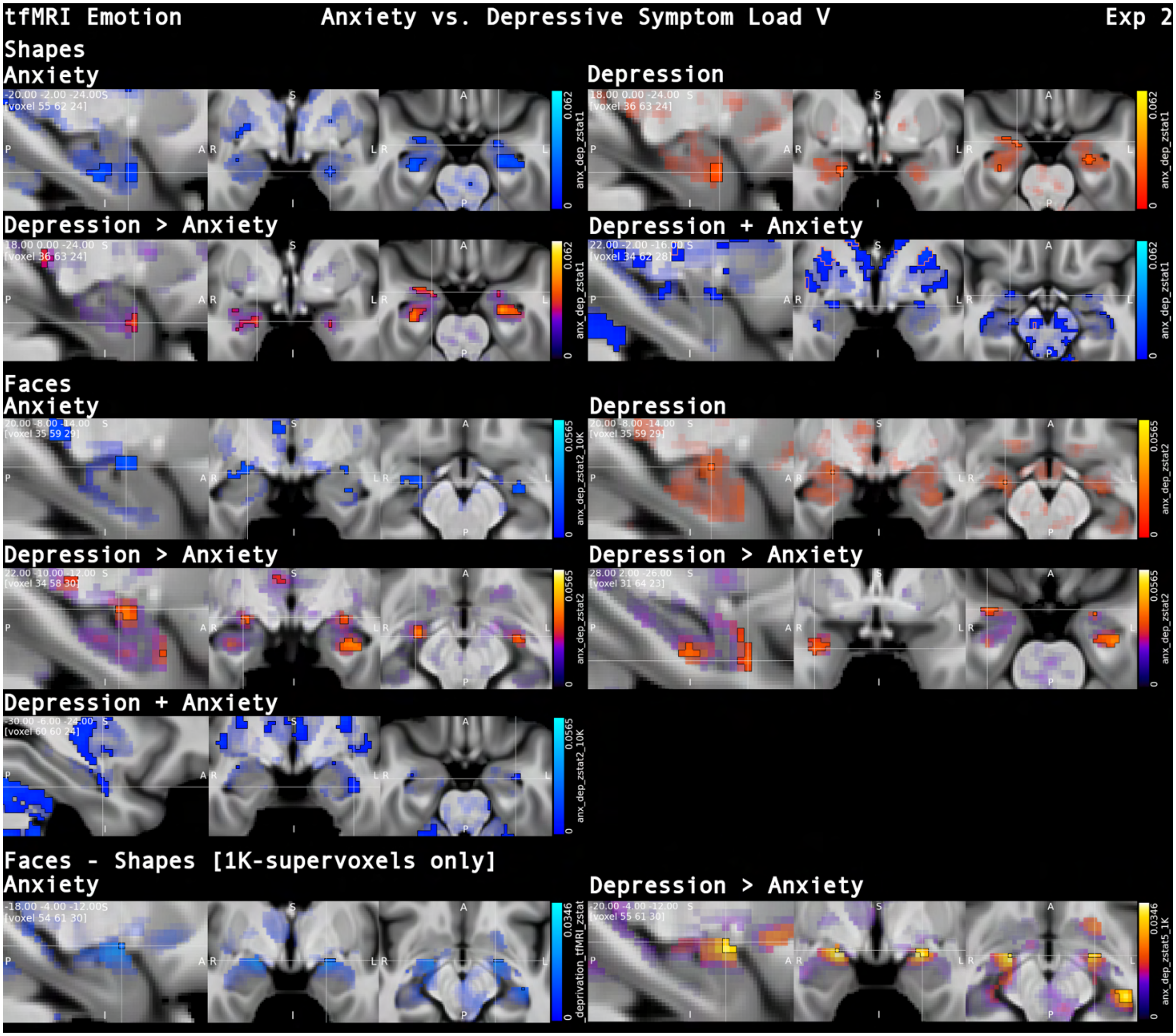


***Figure S2.5. Anxiety and depressive symptom load: Revisiting amygdala reactivity to negative faces.*** *Anxiety and depressive symptoms were associated with spatially and directionally distinct patterns of amygdala responses to neutral shapes and negative faces. While anxiety symptom load generally decreased amygdala reactivity, depressive symptoms exhibited an opposite relationship. A relation to faces>shapes responsiveness for depression over anxiety was detected only when using the more sensitive 1K-supervoxels. (See figure S2.1. for visualization conventions.)*

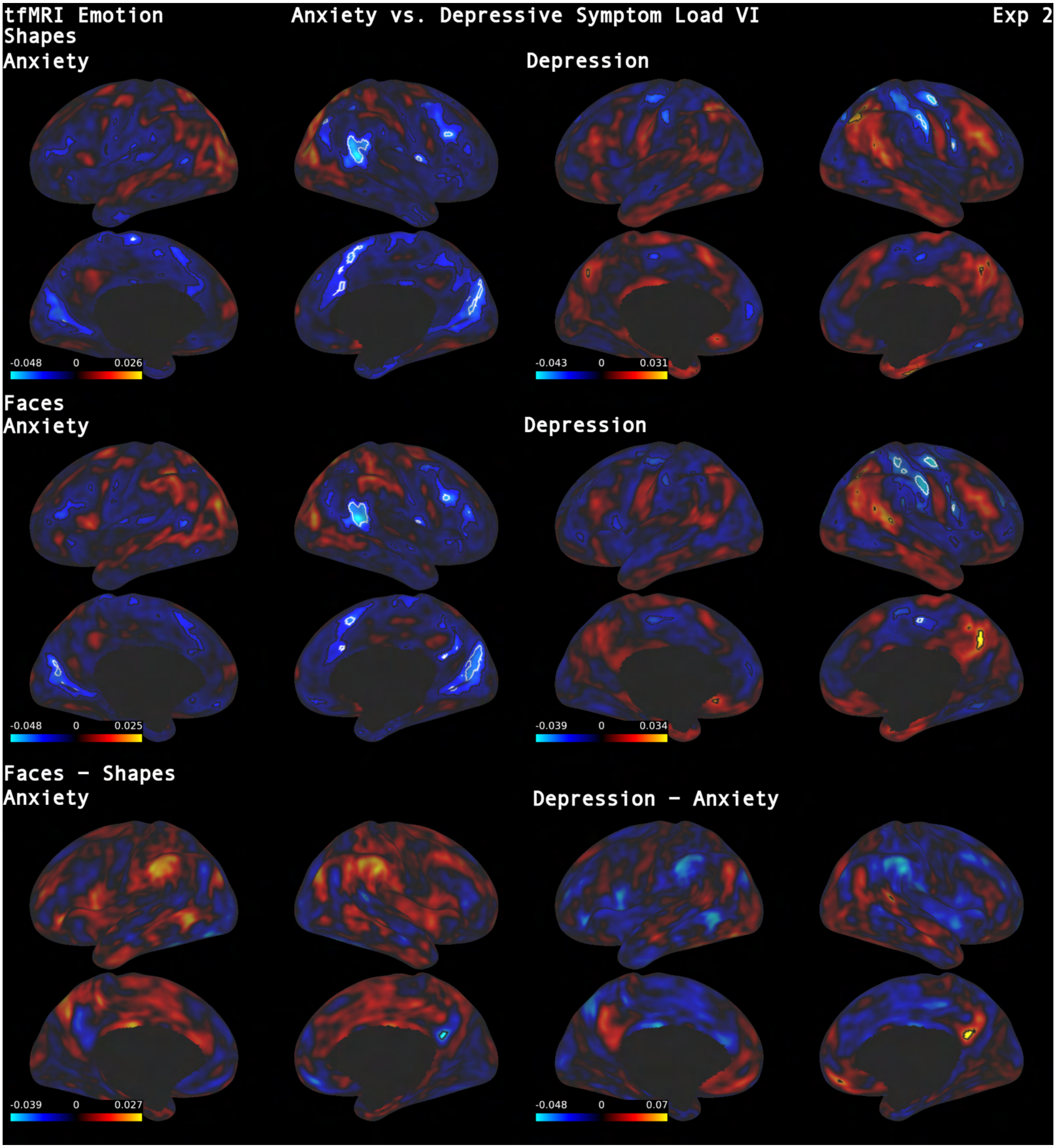


***Figure S2.6. Anxiety and depressive symptom load: Emotion task (surface).*** *Consistent with the decreased amygdala activation, anxiety symptoms related to decreased cortical activity in salience related regions as well as dlPFC. Depressive symptoms related to decreased somatomotor activation and increased activity in the precuneus. The faces–shapes contrast exhibited decreased preferential responsiveness to faces over shapes (relative to group-level image) of posterior ACC for anxiety symptoms only and significant when contrasting against depressive symptom load. Interestingly, posterior ACC has been recently shown to encode face familiarity (Afzalian & Rajimehr, 2021). (See supplementary figure S2.1. for visualization conventions.)*

**NODDI ICVF:** Strong evidence was found for reduced ICVF with anxiety in a small cluster at the white matter (WM)/ grey matter (GM) boundary of postcentral gyrus. No positive association was found. For depression evidence strongly suggested increased ICVF in the posterior medulla oblongata, and decreased signal in bilateral thalamus and extended regions of fronto-parietal white matter.

**NODDI OD:** Depressive symptoms exhibited strong evidence of positive association with bilateral hypothalamus and distributed patches along subcortical and cortical WM/GM boundaries, as well as negative association with bilateral external and internal capsule.

Anxiety showed significant decrease in orientation dispersion in the corpus callosum. There were no significant areas with positive association.

**DTI MD:** Depression score showed strong evidence for positive association with increased MD in cortico-striatal and parietal white matter, and bilateral thalamus. Anxiety score was positively associated with small patches of white matter throughout the brain

**DTI FA:** There was strong evidence for a negative association of depression with FA in bilateral internal capsule and disseminated supra- and infratentorial white matter tracts. FA positively associated with anxiety score at the cortex/white matter boundary next to the left callosal sulcus. Combined anxiety + depressive scores exhibited strong evidence of positive association with FA in temporal fibers of the anterior commissure (adjacent to amygdala).

**RSN d25-2 (DMN):** Anxiety symptom load exhibited moderate evidence of increased coupling of right precuneus (positive association) within the default mode network with significantly increased anticorrelation (negative association) of right dmPFC and supramarginal gyrus. We found no association for depressive symptom load.

**RSN d25-9 (SN):** Depressive symptom load exhibited strong evidence for increased coupling of left (and moderate evidence for contralateral) precuneus within the salience-like RSN d25-9 network (positive association). Moderate evidence was further found for reduced coupling with higher depression score in the border region of left premotor and motor cortex (M1), as well as left cuneus (negative association).

**tfMRI zstat (shapes):** Anxiety scores were broadly negatively associated with response to neural shapes (strong evidence) in right temporo-parietal junction, angular gyrus, dmPFC, dlPFC, right frontoparietal operculum, left anterior hippocampus, left fusiform cortex, left premotor/motor cortex, and bilateral cuneus directly posterior to parieto-occipital fissure. Depression was negatively associated (strong evidence) with fMRI response in right pre- and postcentral gyrus and superior parietal lobule; bilateral caudate nucleus, left putamen, and cerebellum with strong support for activation in left VIIIa being lower than for anxiety load.

Combined anxiety and depressive load exhibited additional strong evidence negative associations in bilateral red nuclei and right substantia nigra and paraventricular thalamic region, bilateral insula, as well as more extensive striatal, cortical (more clearly involving, e.g., supplementary motor area (SMA) and ACC) and cerebellar negative associations.

**tfMRI zstat (faces):** fMRI response to faces was similar to that for shapes. Anxiety again exhibited strong-evidence negative associations in right temporo-parietal junction, dm-/dlPFC, right frontoparietal operculum, and bilateral cuneus. Strong-evidence negative associations were, moreover, seen in right precuneus, bilateral lingual gyrus, right LGN and pulvinar. Depressive symptom load was negatively associated with strong evidence in right pre- and postcentral gyrus and right SMA; bilateral caudate nucleus and cerebellum. Strong support was seen for additional negative associations with combined depressive and anxiety load in left superior colliculus bordering on periaqueductal grey (PAG), paraventricular thalamic area, left red nucleus, bilateral insula, bilateral cingulate cortex and SMA, and again overall more negative signal across cortex, striatum and cerebellum.

**tfMRI zstat (faces − shapes):** Anxiety was negatively associated with higher responsiveness to faces over shapes in right precuneus, i.e., reduced preference relative to group level activation. This finding extended more strongly to the anxiety−depression score difference which was negatively associated in precuneus with moderate evidence, together with small significant regions in right mPFC and supramarginal gyrus.

### Experiment 3: *EPHA3* (rs987748)

Robust evidence was found for rs987748-associated effects across all tested sub-modalities (Figs. S3.1 – 3.5), showing distributed patterns across major white-matter pathways, alongside highly localized signals in the anterior commissure and further cortical/subcortical associations.

**VBM:**

The minor variant of rs987748 was most strongly associated with increased grey matter signal in midline anterior commissure and temporal anterior commissure / uncinate fascicle region. Volume increase was, moreover, seen with strong evidence across frontal, and most prominently parieto-occipital cortices. Areas with highest evidence were parietal operculum, left Heschl’s gyrus (inferior limiting sulcus of insula), bilateral pulvinar, LGN, left cingulate sulcus, bilateral calcarine sulcus (V1), left angular gyrus, and occipital gyri. Other strong evidence areas included orbitofrontal cortices, anterior and posterior cingulate cortices, superior temporal cortices, bilateral LGN, supramarginal gyri, and cerebellum.

Volume decrease was seen most robustly for temporal lobes, specifically, in fusiform cortices and middle temporal gyri. Other strong evidence areas included bilateral anterior insula, amygdalo-striatal transition area, dorsal thalamus, right anterior hippocampus, PAG, bihemispheric lateral occipital cortex, and cerebellum (crus I/II, I-IV, VI).

**SWI QSM:**

QSM for the minor variant was most robustly increased in anterior commissure. Further areas with strong evidence for increase were found in left putaminal surface, bilateral superior temporal and fusiform cortices, right anterior hippocampus, right Heschl’s gyrus, middle longitudinal fascicles, and occipital lobe white matter.

QSM decrease was seen with highest evidence in capsula interna (genu) and periventricular white matter, between cingulum bundle and corticospinal tracts. Further areas of strong evidence included bilateral temporal pole, substantia innominata, centromedial amygdalae, bilateral putamen, dorsolateral thalamus, posterior corpus callosum, parieto-occipital and cerebellar white matter.

**SWI T2*:**

T2* exhibited strong evidence for positive relation with the minor variant in right pulvinar, bilateral middle longitudinal fascicle and straight sinus.

Strong evidence for T2* decrease was seen in right basal operculum, bilateral orbitofrontal cortices (medio-posterior), inferior temporal gyrus, fusiform gyrus with adjacent white matter, right middle temporal gyrus, amygdalo-striatal transition area, right posterior insula and Heschl’s gyrus, bilateral optic radiation, lateral occipital grey and white matter.

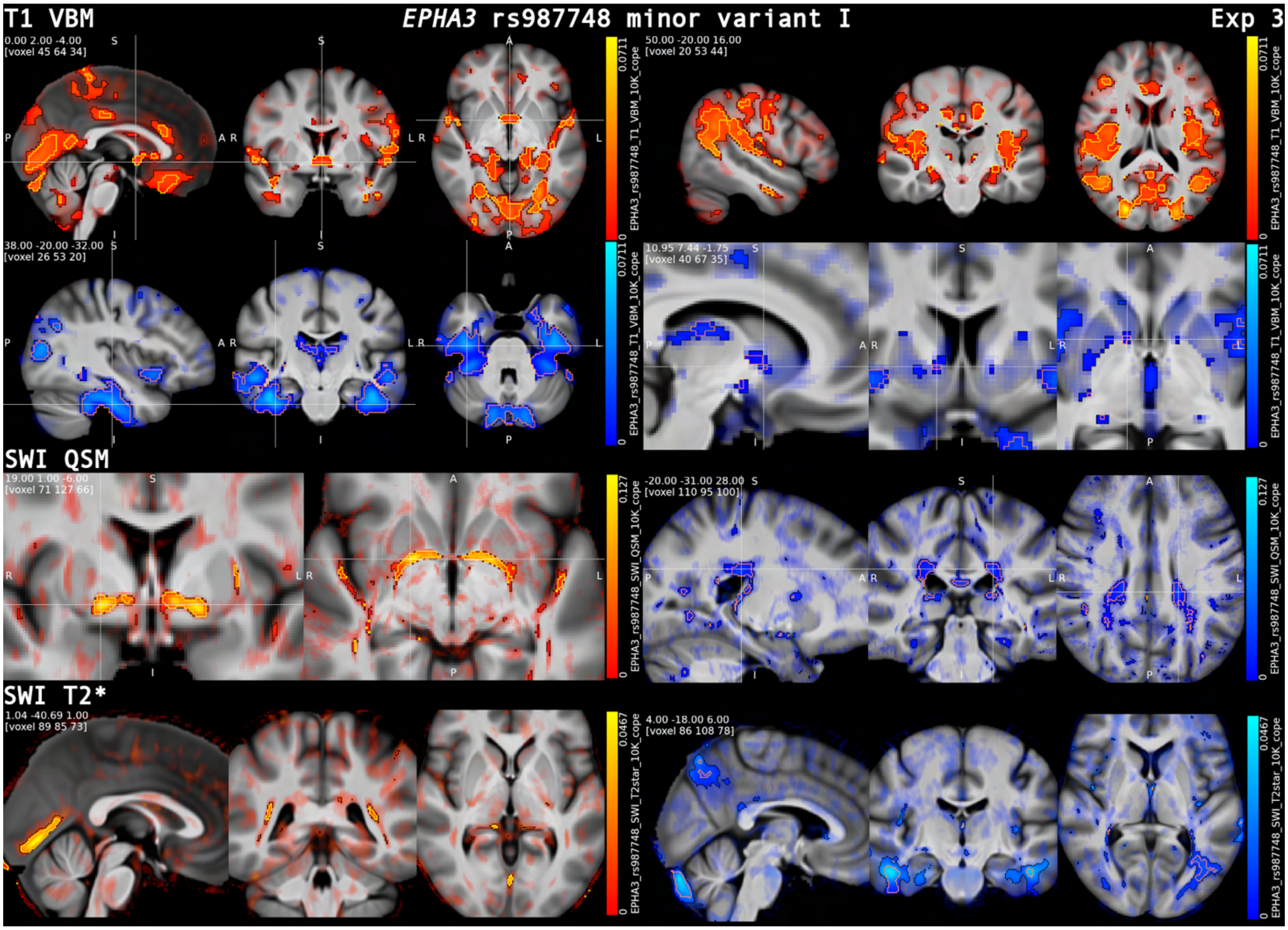


***Figure S3.1. EPHA3 rs987748: VBM and SWI.*** *The rs987748 minor allele showed a convergent signature across modalities centered on interhemispheric and temporal connectivity pathways. VBM revealed robust anterior commissure involvement alongside distributed fronto-parieto-occipital volume increases and temporally weighted volume decreases. SWI measures highlighted a particularly strong positive association with anterior commissure QSM (compatible with decreased myelin signal) , and additional cortical/subcortical susceptibility and T2* alterations extending into temporal and occipital cortices, consistent with widespread, pathway-anchored structural differences associated with this axon-guidance-related variant. Common visualization conventions (Figs. S4.1–4.5): β-maps thresholded at p_hFDR < 0.05 (black outline) overlaid on non-significant effects modulated by Z-statistic (surface views: non-significant effects shown by transparency, without Z-modulation), over the mean T1-weighted image. Positive associations are shown in red and negative associations in blue, contrast associations are displayed with a heatmap (NIH-fire); p_hFDR < 0.001 is outlined in yellow (positive and contrasts) or orange (negative), except on surface renderings, where p_hFDR < 0.001 is indicated in white. Group-mean resting-state network maps (Z-statistics) are thresholded at Z > 2.58 (approx. p < 0.01, two-tailed).*

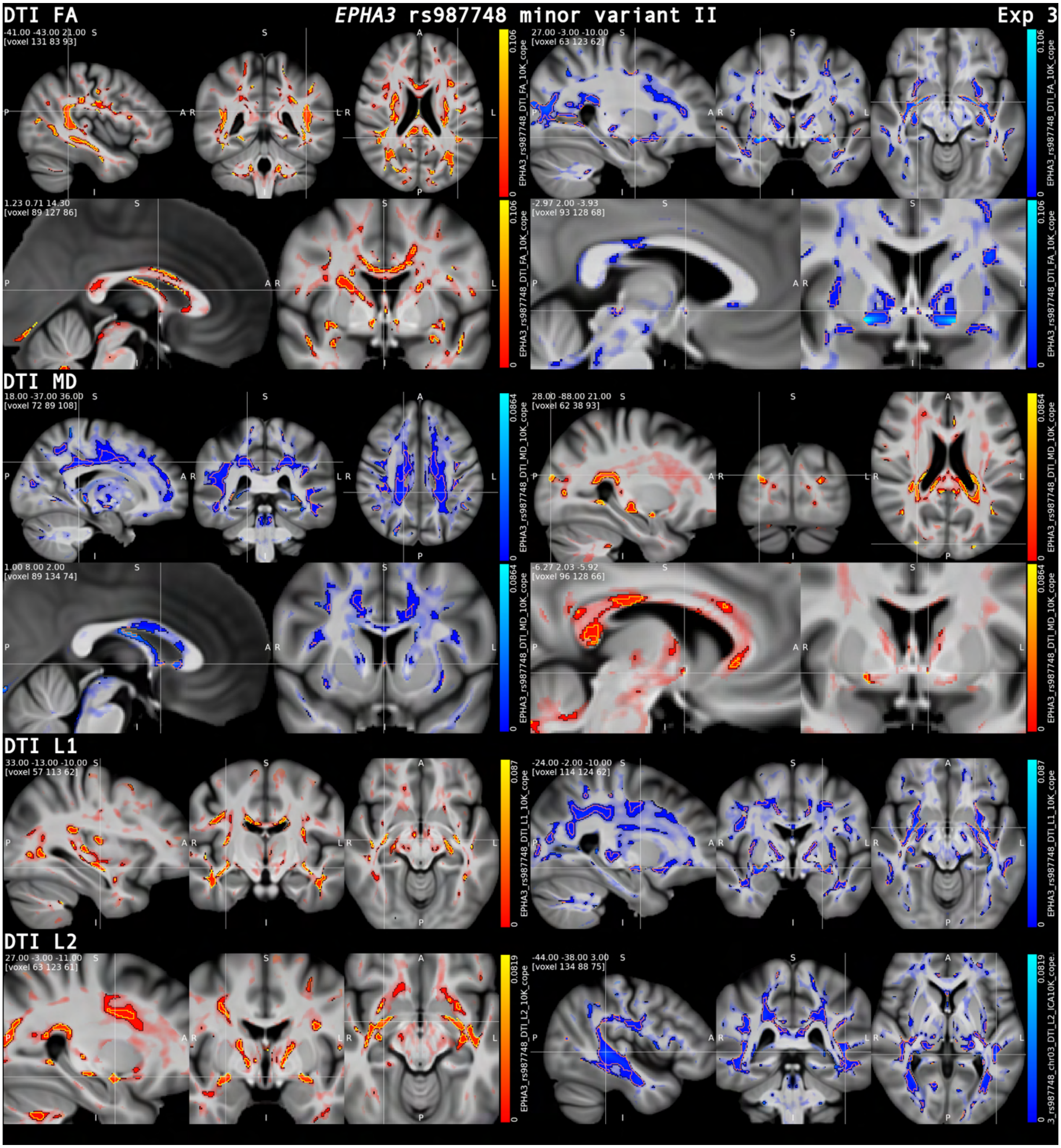


***Figure S3.2. EPHA3 rs987748: DTI.*** *Diffusion metrics showed anatomically structured rs987748 associations with prominent effects in the anterior commissure and distributed long-range pathways. Across DTI indices, significant clusters were reported in the temporal anterior commissure/uncinate region, posterior corpus callosum, arcuate and superior longitudinal fascicles, optic radiations, and occipital white matter. Notably, positive and negative associations frequently occurred in close spatial proximity (often adjacent along the same tract), consistent with fine-grained, boundary- and geometry-sensitive microstructural differences. (Visualization convention: Fig. S4.1.)*

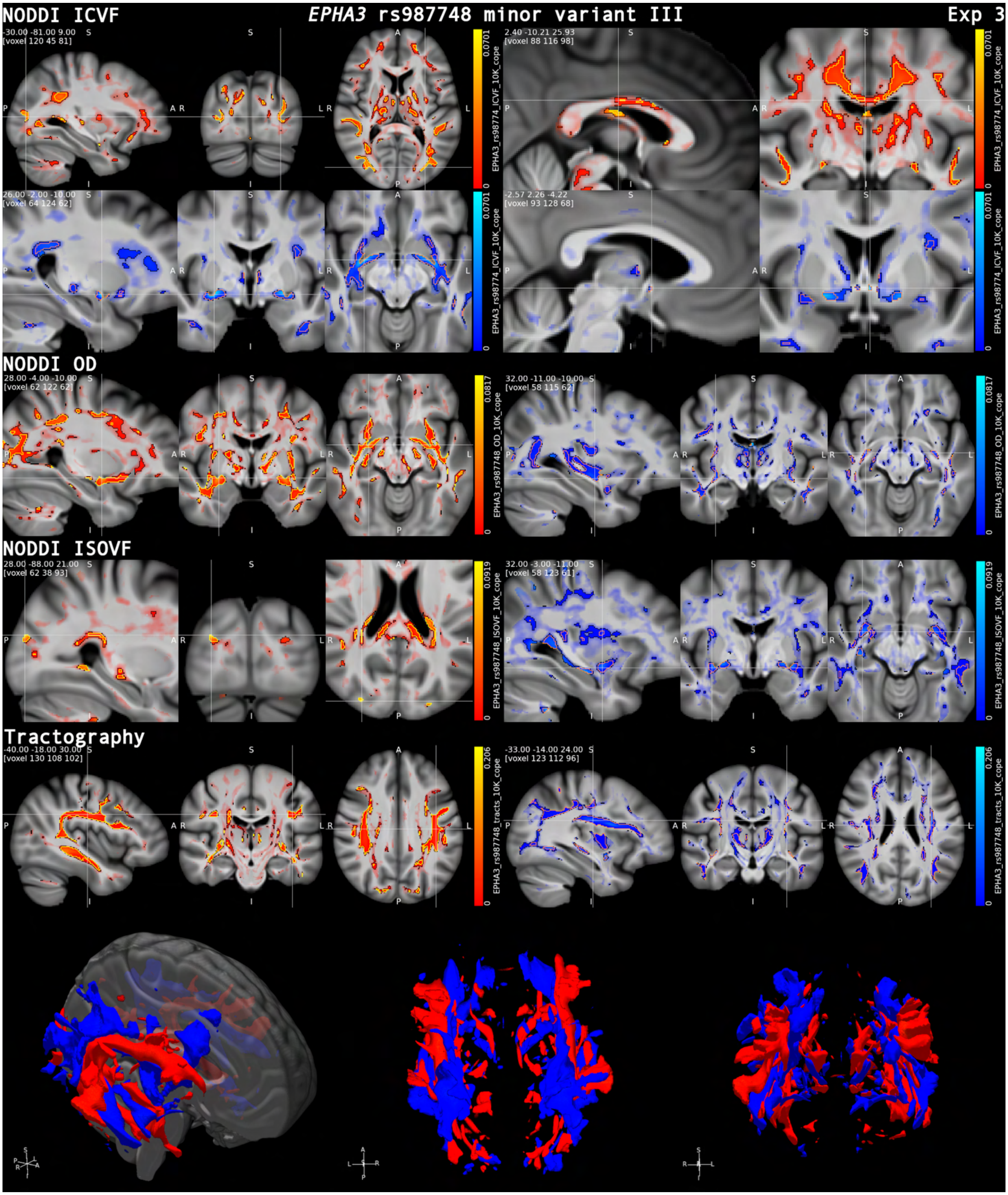


***Figure S3.3. EPHA3 rs987748: NODDI and tractography.*** *Robust rs987748 associations across commissural and distributed long-range pathways, with additional prominent effects in U-shaped fibers in temporal and occipital lobe. As for DTI, positive and negative associations were often directly adjacent along the same tract, suggesting fine-grained phenotypic variation (Visualization convention: Fig. S4.1.)*

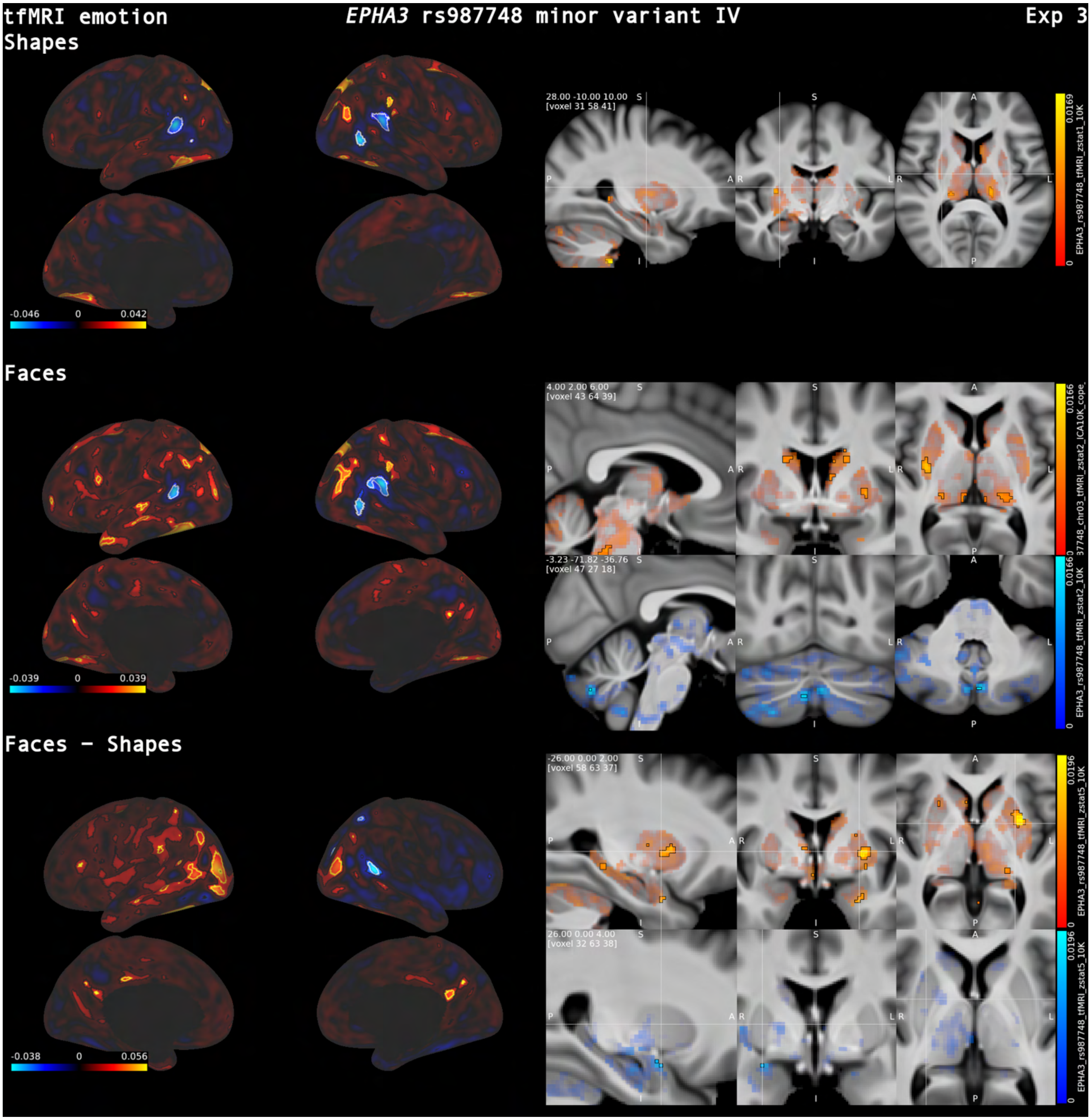


***Figure S3.4. EPHA3 rs987748: tfMRI emotion****. In the emotion task, rs987748 showed robust activation differences for shapes and faces, with consistent involvement of fusiform and inferior temporal cortex, supramarginal/angular gyri, and parieto-occipital transition areas, alongside frontal (dmPFC/vlPFC) and cingulate components. The faces−shapes contrast additionally highlighted occipito-temporal and posterior cingulate effects with subcortical involvement (left putamen). (Visualization convention: Fig. S4.1.)*

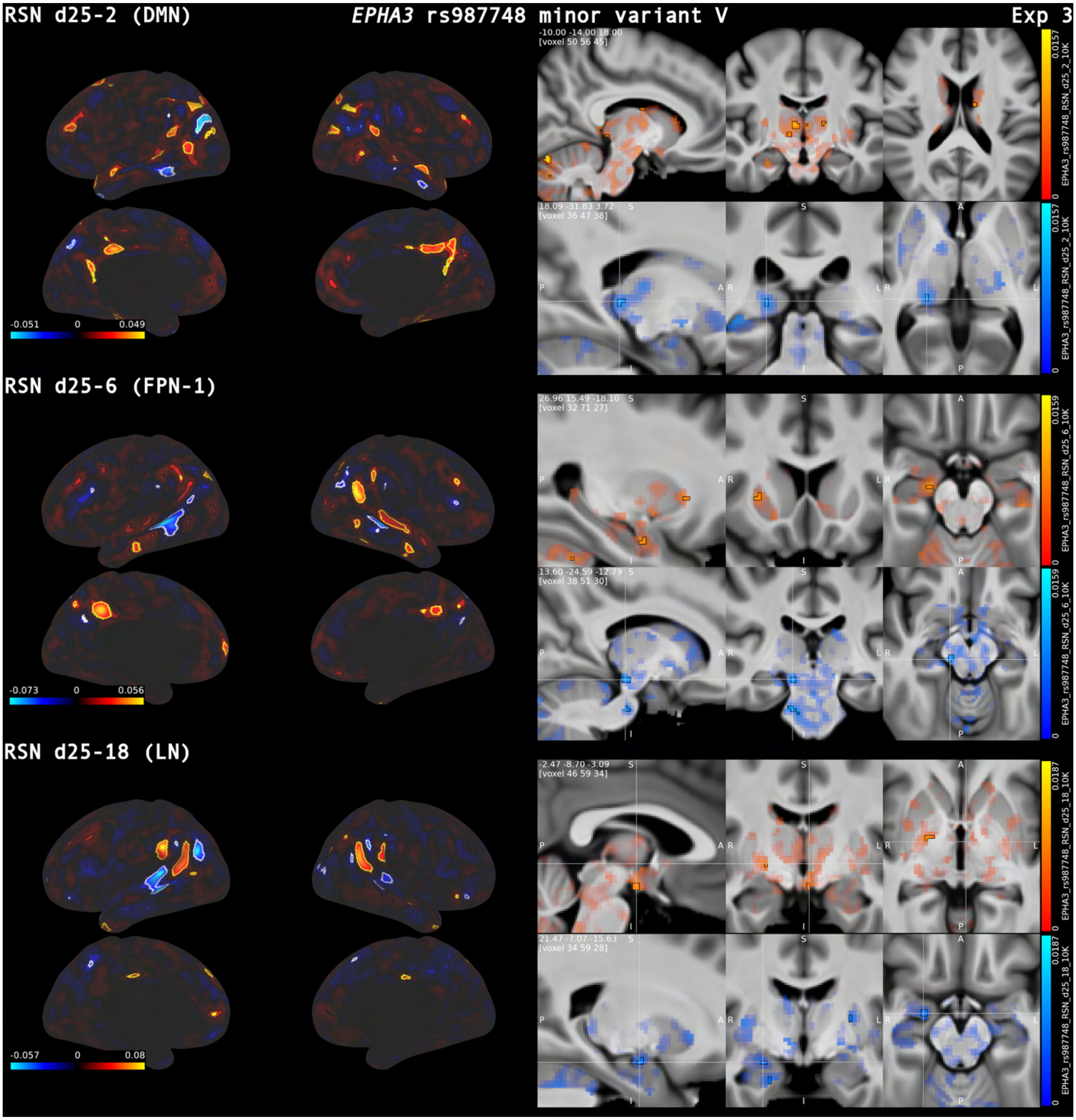


***Figure S3.5. EPHA3 rs987748: RSN.*** *Resting-state network analyses revealed rs987748-associated shifts in coupling across the DMN (RSN d25-2), a fronto-parietal network (RSN d25-6) and a language-like network (RSN d25-18). Effects were strongest in posterior midline and temporo-parietal hubs (e.g., posterior cingulate/precuneus, temporo-parietal junction, supramarginal/angular gyri) with additional involvement of prefrontal cortex and subcortical regions. Analogous to structural findings in diffusion imaging, coupling increases and decreases were often spatially adjacent within the same network, producing paired "up/down" patterns rather than uniformly signed shifts. (Visualization convention: Fig. S4.1.)*

**DTI MD:**

Diffusivity was increased for the minor variant with most robust evidence in temporal part of anterior commissure, bilateral LGN, and posterior callosum. Further increases with strong evidence included left ACC (grey matter/white matter boundary), bilateral acoustic radiation, arcuate fascicle, and lateral occipital cortex / white matter.

MD decrease was related most robustly seen in bilateral amygdalo-striatal transition area posterior to anterior commissure, uncinate fascicles, superior temporal gyrus, pulvinar, between posterior cingulum bundle and superior longitudinal fascicle, in arcuate fascicles, and occipital white matter. Further areas included radiation of corpus callosum, and small patches of frontal white matter.

**DTI FA:** Strongest evidence for increased FA was found in the paraventricular hypothalamic region, directly posterior to anterior commissure, adjacent to lateral amygdalae, in U-fibers connecting middle and superior temporal gyri, left uncinate fascicle, bilateral posterior putamen, mediodorsal thalamus, right Heschl's gyrus, right arcuate fascicle, bilateral optic radiation, and occipital short association fibers. Further strong evidence was found for FA increase in middle and posterior corpus callosum, bilateral FAT, and cerebellar white matter.

The minor variant exhibited most robust evidence for FA decrease in bilateral anterior commissure, inferior fronto-occipital fascicles (temporal part), bilateral LGN, arcuate fascicles, superior longitudinal fascicles, bilateral tapetum and stratum, and cerebellar white matter.

**NODDI ICVF:** Strongest evidence ($-\log_{10}(p)$ mapFDR > 10) for positive association of minor variant with neurite density was found for U-shaped fibers between middle and superior temporal gyri, along right middle longitudinal fasciculus, bilateral arcuate fasciculi, bilateral amygdalo-striatal transition area, in callosal fibers lateral of bilateral cingulum bundles, between superior longitudinal fasciculi 1 and 2, in posterior callosum, and in U-shaped fibers along occipital gyri. Further areas of strong evidence included thalamic radiations, right fornix, and left capsula externa.

Robust evidence for negative association with minor variant ($-\log_{10}(p)$ mapFDR > 60) was found for temporal parts of bilateral anterior commissure with further very strong evidence regions in bilateral LGN and tapetum (>10). Other strong evidence regions included fibers adjacent to sulcal fundi in perirhinal cortices, parahippocampal, fusiform and inferior frontal gyri as well as white matter deep in superior temporal gyri; bilateral arcuate fascicles, U-shaped and planar fibers along occipital gyri, and cerebellar white matter.

**NODDI isotropic volume fraction (ISOVF):** ISOVF increase was most evident ($-\log_{10}(p)$ mapFDR > 10) in posterior callosal fibers, with further strongly supported increases seen co-localized with U-shaped fibers seen for ICVF between middle and superior temporal gyri, in right middle longitudinal fasciculus, adjacent to bilateral LGN, and in vertical occipital fasciculi.

ISOVF decrease was supported most strongly ($-\log_{10}(p)$ mapFDR > 15) in bilateral anterior commissure (lateral parts); further strong evidence regions included bilateral pulvinar, arcuate fascicles, inferior and middle longitudinal fascicles, bilateral optic radiations, planar white-matter patches along parietal and occipital cortices.

**NODDI OD:** Increase of orientation dispersion with the minor variant in lateral anterior commissure was one of the most statistically significant findings of the study with $-\log_{10}(p)$ mapFDR above 100. Further strong evidence was found in bilateral capsula interna and externa, bilateral cingulum bundle, fronto-basal white matter, inferior and middle longitudinal fasciculi, arcuate fasciculi, superior longitudinal fasciculi, vertical occipital fasciculi, and in mesencephalic and cerebellar white matter.

Evidence was strongest for decrease (>10) directly posterior to anterior commissure close to the midline, left FAT, in posterior callosum, bilateral optic radiations, arcuate fasciculi, U-shaped white matter between middle and superior temporal gyri, and amygdalo-striatal transition area. More strong evidence alterations were found in anterior callosum, thalamic radiations, hypothalamic region, superior longitudinal fasciculi, parieto-occipital white matter, between substantia nigra and red nuclei, and in cerebellar white matter.

**Tractography:** Robust evidence ($-\log_{10}(p)$ mapFDR > 60) was found for positive association of minor variant in the region of bilateral arcuate fascicle and superior longitudinal fascicle 3**.** Further regions of very strong evidence ($-\log_{10}(p)$ mapFDR >10) included bilateral uncinate fascicles, anterior commissure, FAT, inferior fronto-occipital fasciculi, U-shaped fibers between middle and superior temporal gyrus, bilateral optic radiations. Further regions of strong evidence included occipital and cerebellar white matter.

Decrease was most robustly supported with adjacent regions of left arcuate fascicle (>60) followed by right fascicle (>30), and U-shaped fibers between postcentral gyrus and supramarginal gyrus. Further regions of very strong evidence (>10) included anterior commissure and external capsule. Other regions of strong evidence included middle longitudinal fascicles, and right FAT.

**RSN d25-2 (DMN):** Evidence for increased coupling with minor variant was most robust in left posterior cingulate cortex ($-\log_{10}(p)$ mapFDR > 15). Further strong evidence increases included bilateral dlPFC, left dmPFC, temporal cortices, supramarginal and angular gyri, right precuneus, occipital gyri and left cerebellum. Coupling was decreased with strong evidence in adjacent regions in temporal cortex, left angular gyrus, and left precuneus.

**RSN d25-6 (FPN-1):** Coupling was increased with most robust evidence in left precuneus followed by right temporo-parietal junction ($-\log_{10}(p)$ mapFDR >15; >10). Further strong evidence regions were found in right precuneus, across temporal cortices, right dlPFC, left mPFC, bilateral supramarginal and angular gyri.

Evidence for decreased coupling was most robust in bilateral middle temporal gyrus ($-\log_{10}(p)$ mapFDR > 20). Other regions with strong evidence for decrease included bilateral vlPFC, bilateral precuneus, supramarginal gyri and right angular gyrus.

**RSN d25-18 (LN):** Coupling in the language-like network RSN_d25-18 was most robustly increased in left supramarginal gyrus and right temporo-parietal junction ($-\log_{10}(p)$ mapFDR > 20). Other strong evidence areas included bilateral temporal pole, right anterior insula, bilateral mid/posterior cingulate cortex.

Coupling was most robustly decreased in adjacent areas in left superior temporal cortex and angular gyrus ($-\log_{10}(p)$ mapFDR > 20; >15). Other strong evidence areas with decrease included bilateral middle temporal cortex, bilateral vlPFC, right dmPFC, left precuneus, and right occipital gyri.

**tfMRI zstat (shapes):** Increased activation was strongly supported for the minor variant in right dmPFC, bilateral fusiform cortices, inferior temporal gyri, right supramarginal and posterior angular gyrus and bilateral parieto-occipital transition area. Strongest decrease ($-\log_{10}(p)$ mapFDR >10) was seen in bilateral supramarginal gyrus/superior temporal gyrus and posterior part of right middle temporal gyrus.

**tfMRI zstat (faces):** Minor variant exhibited strongly supported increase in a more scattered pattern across frontal, temporal and parietal regions, again including fusiform cortices, parieto-occipital transition and supramarginal regions. Decrease was strongly supported in posterior parts of superior temporal gyri and right middle temporal gyrus.

**tfMRI zstat (faces − shapes):** Strongest support ($-\log_{10}(p)$ mapFDR >10) was found for increased fMRI response in left occipital cortex posterior to middle temporal gyrus. Further strong evidence regions were found in left vlPFC and dmPFC, bilateral fusiform cortices, left middle temporal gyrus, inferior temporal gyri, left mid- and posterior cingulate cortex, right posterior cingulate cortex, left supramarginal gyrus, bilateral angular gyrus, temporo-occipital area and left putamen. Strong evidence for decreased activation was found in right middle temporal gyrus, right superior parietal lobule, and bilateral occipital lobes.

### **Experiment 4:** Multimodal effects of separate polygenic scores derived from early- and late-diagnosis autism latent factor GWAS

Multimodal associations of the early- and late-diagnosis autism polygenic scores were found in 6 / 10 tested sub-modalities (Figs. S4.1–4.3), revealing dissociable white-matter signatures as well as convergent effects in amygdala.

**T1 VBM:**

Moderate evidence was found for decreased grey matter left cerebellum (VI / crus I) for early-ASD polygenic score (PS), while evidence supported decrease in contralateral side. The additive combination of early- + late-ASD PS was further associated with increased volume in left amygdala and bilateral cerebellum (crus I). Left dlPFC and right putamen exhibited greater volume for late-ASD than for early-ASD PS.

**SWI T2*:** T2* was not significantly associated with either early- or late-ASD PS using the 10K-supervoxels. Using the more sensitive 1K-supervoxels, left mid/anterior hippocampus was associated with early-ASD PS (significant after hierarchical FDR correction within 1K analysis using same sub-modalities as for primary analysis) with moderate evidence, and to a lesser extent to the additive combination of both scores.

**SWI QSM:** Same as for T2*, no associations were found using 10K-supervoxel analysis. Using 1K-supervoxels, early-ASD PS was associated with moderate evidence with decrease in a small cluster in right posterior insula and supported increase in left posterior putamen (surface region). Late-ASD PS was associated with decreased QSM in anterior commissure, bilateral pallidum and left substantia nigra (bordering on white matter).

**NODDI ICVF:** Neurite density was positively associated with late-ASD PS in frontal and fronto-basal white matter, as well as between bilateral forceps major and cingulum bundle, superior temporal white matter, and fusiform pathways. Negative association with ICVF was found in bilateral pallidum. Early-ASD PS was associated with decreased ICVF (moderate evidence) in a longitudinal segment of white matter between right corticospinal tract and cingulum bundle, and with decrease in anterior callosum. Moderate evidence was further found in direct contrast with early-ASD PS for higher density in frontal lobe white matter, right inferior claustrum, white matter between corticospinal tract and cingulum bundles, optic radiations, and occipital white matter.

**NODDI OD:** Moderate evidence was found for increased orientation dispersion for late-ASD PS in right uncinate fascicle and more clearly in direct contrast with early-ASD PS. Evidence further supported decreased OD for right cerebellar white matter for early-ASD PS and combined early- and late-ASD PS. Right pallidal OD was higher for early-ASD PS than for late-ASD PS in a small significant cluster.

**DTI FA:** Moderate evidence supported decreased FA adjacent to right claustrum, anterior callosum, left posterior cingulate bundle, and across bilateral corticospinal tract for early-ASD score. Evidence supported higher FA for late-ASD score than early-ASD along bilateral amygdala, posterior capsula externa, callosum and in longitudinally oriented patches across frontal, parietal and occipital white matter. FA was not significantly increased for either early- or late-ASD PS.

**DTI MD:** Evidence supported higher MD for early-ASD PS localized in left FAT. Late-ASD PS was not significantly associated with MD. Direct contrasting between the two scores, however, revealed widespread higher MD signal for early-ASD PS centered on frontal lobe and extending into parieto-occipital white matter.

**tfMRI zstat (shapes):** No significant associations were found for shapes using 10K analysis.

**tfMRI zstat (faces):** Moderate evidence was found for negative association in right precuneus in response to negative faces for the combination of high early-ASD plus late-ASD PS, signifying increased deactivation with respect to the group-level mean, while evidence supported increased deactivation in left precuneus also.

**tfMRI zstat (faces − shapes):** No significant associations were found for faces over shapes using 10K analysis. An exploratory analysis with the more sensitive 1K-supervoxels (across same sub-modalities) supported widespread cortical and subcortical reductions in fMRI response for early-ASD and combined early-ASD + late-ASD scores.

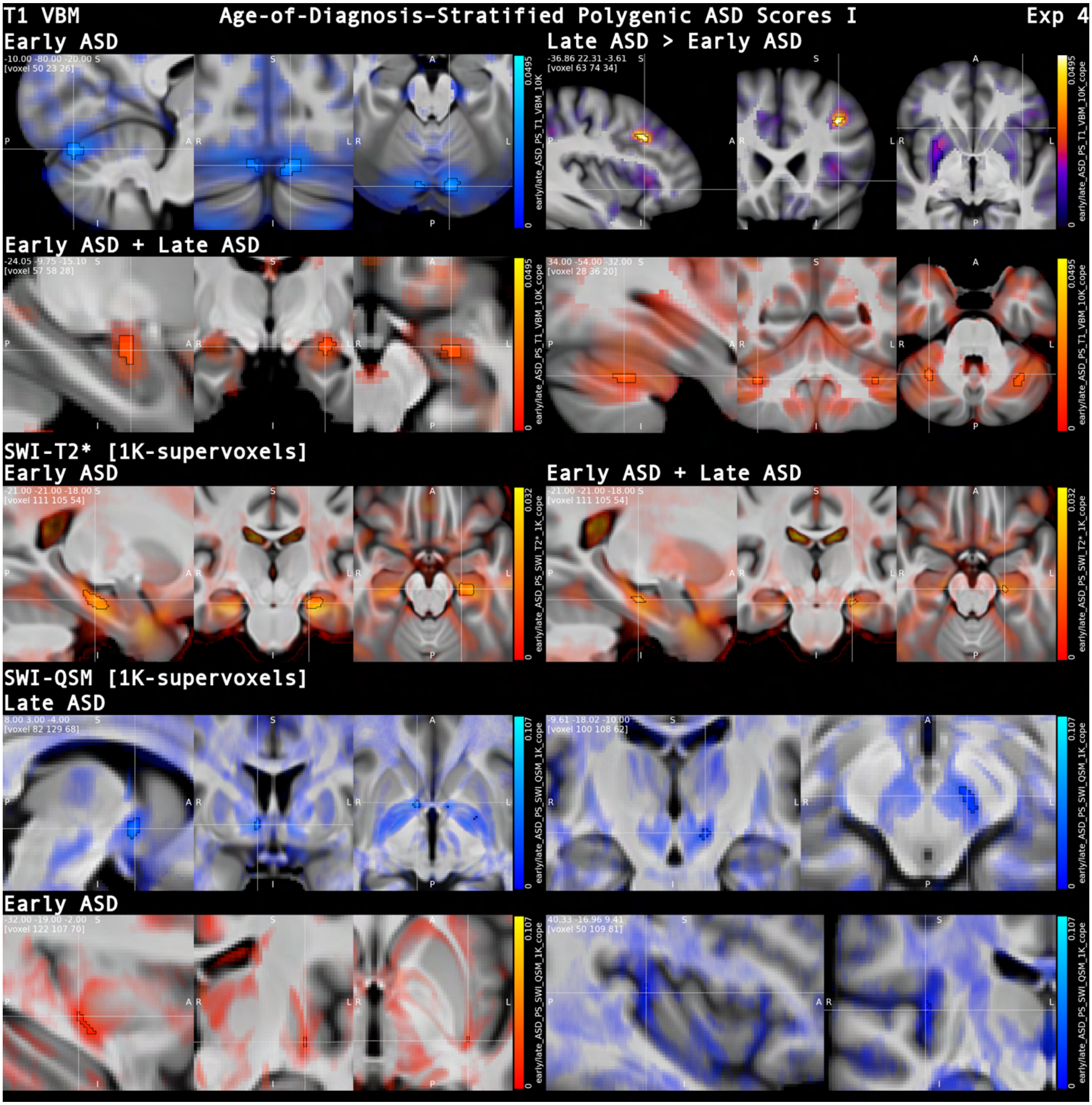


***Figure S4.1. Early/late-diagnosis ASD polygenic score: VBM and SWI.*** *VBM showed distinct and convergent effects of early- vs. late-ASD polygenic scores, including cerebellar reductions for early-ASD PS and combined early- plus late-ASD score increases in left amygdala; contrasts highlighted greater left dlPFC volume for late-ASD PS. No significant SWI (T2*/QSM) effects were detected with 10K-supervoxels; in another illustration of its higher sensitivity, exploratory 1K-supervoxel analysis revealed focal SWI associations such as increased T2* in hippocampus for early- and combined ASD score and reduced QSM in anterior commissure and substantia nigra for late-ASD score. <u>Common visualization conventions (Figs. S5.1–5.3)</u>: β-maps thresholded at p_hFDR < 0.05 (black outline) overlaid on non-significant effects modulated by Z-statistic (surface views: non-significant effects shown by transparency, without Z-modulation), over the mean T1-weighted image. Positive associations are shown in red and negative associations in blue, contrast associations are displayed with a heatmap (NIH-fire).*

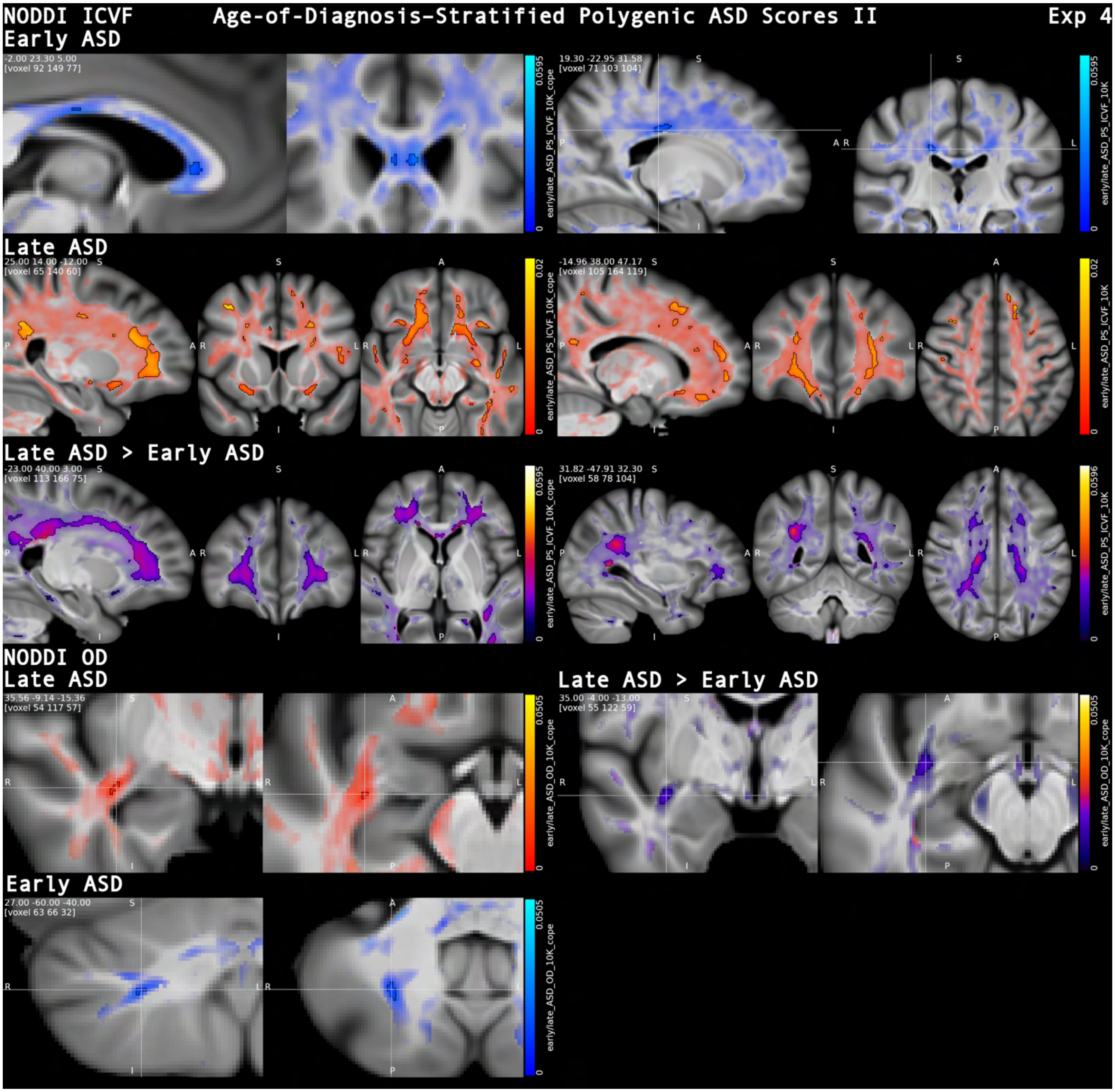


***Figure S4.2 Early/late-diagnosis ASD polygenic score: NODDI.*** *NODDI highlighted dissociable white-matter signatures, with late-ASD PS showing increases in neurite density (ICVF) concentrated in frontal white matter, whereas early-ASD PS showed ICVF reductions in anterior callosum and a longitudinal tract segment between corticospinal and cingulum pathways. Contrast maps further highlighted higher ICVF for late- than early-ASD PS across frontal, temporal and occipital pathways. Late-ASD PS further associated with increased OD in right uncinate fascicle. (Visualization convention: Fig. S5.1.)*

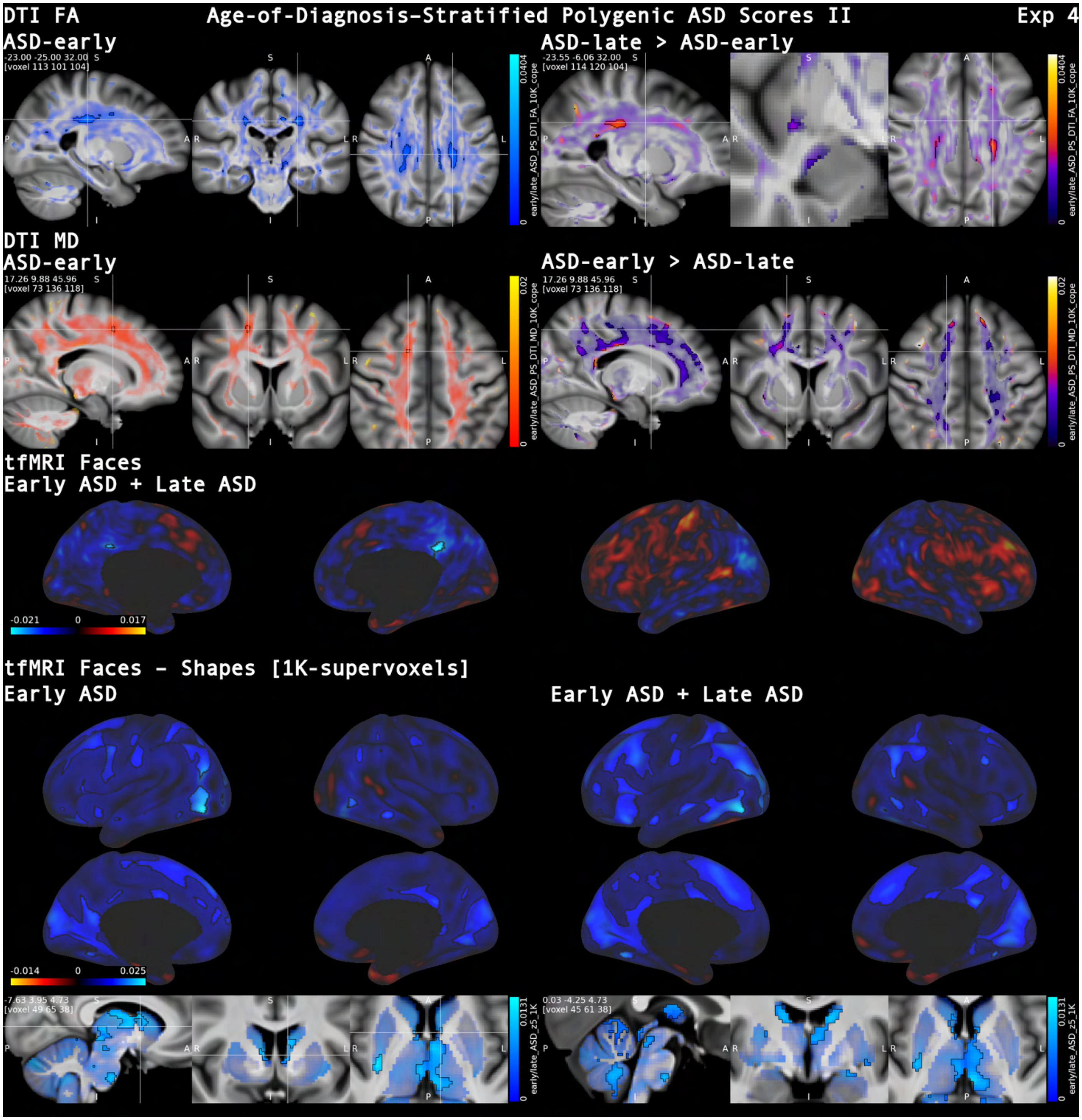


***Figure S4.3 Early/late-diagnosis ASD polygenic score: DTI and tfMRI emotion.*** *DTI showed dissociable microstructural associations for early- vs. late-ASD polygenic scores: early-ASD PS was linked to FA reductions (e.g., anterior callosum, posterior cingulum and corticospinal tracts), whereas late-ASD PS showed higher FA than early-ASD along amygdala-adjacent white matter. Frontal MD was further higher for early-ASD than for late-ASD scores. In tfMRI, the combined scores showed moderate-evidence reduced posterior cingulate cortex/precuneus responses to negative faces; exploratory 1K-supervoxel analysis suggested widespread cortical and subcortical reductions in fMRI responsiveness to faces over shapes. (Visualization convention: Fig. S5.1)*

### **Experiment S1:** broad associations of nicotine, alcohol and coffee with brain structure and function

Substance-related brain differences were found with strong evidence across all 13 tested sub-modalities (Figs. S5.1–S5.12), with both main effects and direct contrasts highlighting distinct spatial signatures of past tobacco use, alcohol intake frequency, and coffee consumption.

**VBM:**

**Smoking:** Strong evidence was found for positive association in bilateral dorsolateral pulvinar (peak left hemisphere; $-\log_{10}(p)$_mapFDR > 10). Bordering voxels in ventricles were also found to be significant, which may fit with a shift in grey matter boundaries. Further areas of strong evidence were found in left ventromedial and dorsomedial PFC, right dlPFC, corpus callosum (presumably related to white matter decrease), right hippocampus (mid-anterior), bilateral parahippocampal gyrus, left paracentral lobule, visual cortex (V1/V2) and cerebellum (I-IV). In contrasting against coffee, vm-/dmPFC volume stood out as being higher in smokers with strong evidence. There was further strong evidence for volume increase along calcarine sulcus (V1) for smoking in contrast to alcohol and coffee consumption.

Volume loss associated with smoking was extensive covering infra-/ and supratentorial grey matter and subcortex with robust evidence ($-\log_{10}(p)$_mapFDR > 15) found across bilateral cingulate cortex, striatum and basal forebrain.

**Coffee:** Strongest evidence ($-\log_{10}(p)$_mapFDR > 15) was found for increased volume in bilateral nucleus accumbens region, substantia innominata, BNST, hypothalamus, ventral tegmental area (VTA) and encompassing the complete PAG. We found further strong evidence for increased grey matter volume in centromedial amygdala and amygdalo-striatal transition area, corpus callosum, right cerebral peduncle bordering on lateral substantia nigra. Strong evidence was found for extensive cerebellar volume increase (I-VI, VIII, IX, X; vermis). Moderate evidence was found for greater bilateral anterior hippocampus volume.

Strong evidence for larger subcortical volumes (striatum, amygdala, thalamus, VTA, PAG) as well as cerebellum was found for coffee consumption contrasting both smoking and alcohol consumption. Higher volume was further found in ACC.

While far less extensive than for smoking, coffee consumption was associated with decreased volume across frontal and parieto-occipital lobe. Some areas of strongest evidence ($-\log_{10}(p)$_mapFDR > 5) were bilateral straight gyrus, right ventrolateral prefrontal cortex (vlPFC), left dlPFC, bilateral precentral gyrus, and lateral occipital gyrus.

**Alcohol:** Strong evidence positive associations were found for right dmPFC, right anterior insula and frontal operculum, corpus callosum and fornix (to be interpreted as white matter loss), right basolateral amygdala extending into uncus and entorhinal cortex, right anterior hippocampus, bilateral parahippocampal cortex, dorsal pulvinar, right parietal operculum, paracentral lobule and precuneus, and temporo-occipital fusiform cortices extending into neighboring cerebellum. Contrasting coffee consumption, strong evidence for higher volume was solely found in bilateral mPFC, bilateral dorsal thalamus / ventricle region and a small region in lingual gyrus. Contrasting against smoking highlighted higher volume in bilateral substantia innominata, and subcallosal area.

Similar to smoking, alcohol consumption revealed extensive strong evidence associations with volume loss across the brain. The pattern differed with most robust evidence ($-\log_{10}(p)$_mapFDR > 20) not only for striatum but also in bilateral precentral gyrus, lateral amygdala, hippocampus, hypothalamus and thalamus.

**SWI QSM:**

**Smoking:** Evidence was most definitive in bilateral putamen ($-\log_{10}(p)$_mapFDR > 50), followed by caudate nuclei and the area between substantia nigra, red nucleus and VTA (> 20). Further strong evidence associations were found in anterior hippocampi, entorhinal cortices, thalamus, habenulae, and supra- and infratentorial sulci, in particular, along cingulate sulcus.

In direct comparisons, QSM was higher for smoking than for coffee consumption in dorsal striatum and for coffee and alcohol consumption medial substantia nigra.

Strong evidence was found for decreases in QSM in white matter associated with smoking, in particular, in fronto-temporal lobes but with highest evidence in capsula interna and externa. A further area of very strong evidence for QSM increase was the bilateral centromedian nucleus (CN) of the thalamus.

Contrast analysis revealed the white matter QSM to be lower in smoking than coffee but not alcohol consumption.

**Coffee:** Similar to smoking but to a lesser extent coffee consumption showed strong evidence for increased QSM in the striatum with most robust evidence, however, for the caudate nucleus rather than the putamen ($-\log_{10}(p)$_mapFDR > 20), and for the genu of capsula interna (consistent with myelin decrease), while peak evidence for QSM increase was found in the right habenula (> 35; > 20 for left habenula). Further areas of strong evidence included bilateral cingulate sulci, medial substantia nigra, habenulae, superior temporal sulcus, left Heschl's gyrus, and scattered parieto-occipital areas. Marked associations were further observed in bilateral cerebellum.

Negative associations were limited to internal capsule white matter and observed to a smaller degree than for smoking, which was also reflected in the direct contrast.

**Alcohol:** Alcohol exhibited a similar pattern of QSM increase across subcortex and sulci. Here most robust evidence was found for bilateral putamen ($-\log_{10}(p)$_mapFDR > 50), followed by caudate nucleus, and cerebellum, with further particularly significant areas (> 10) encompassing bilateral anterior hippocampi, superior temporal sulci, and lateral pulvinar nuclei. Similar to smoking, the area between substantia nigra, VTA and red nucleus exhibited high QSM, consistent with increased myelinization.

Negative associations with QSM were similar to smoking and affected global white matter, in particular, internal and external capsulae and cerebellar as well as bilateral thalamic nuclei with highest evidence.

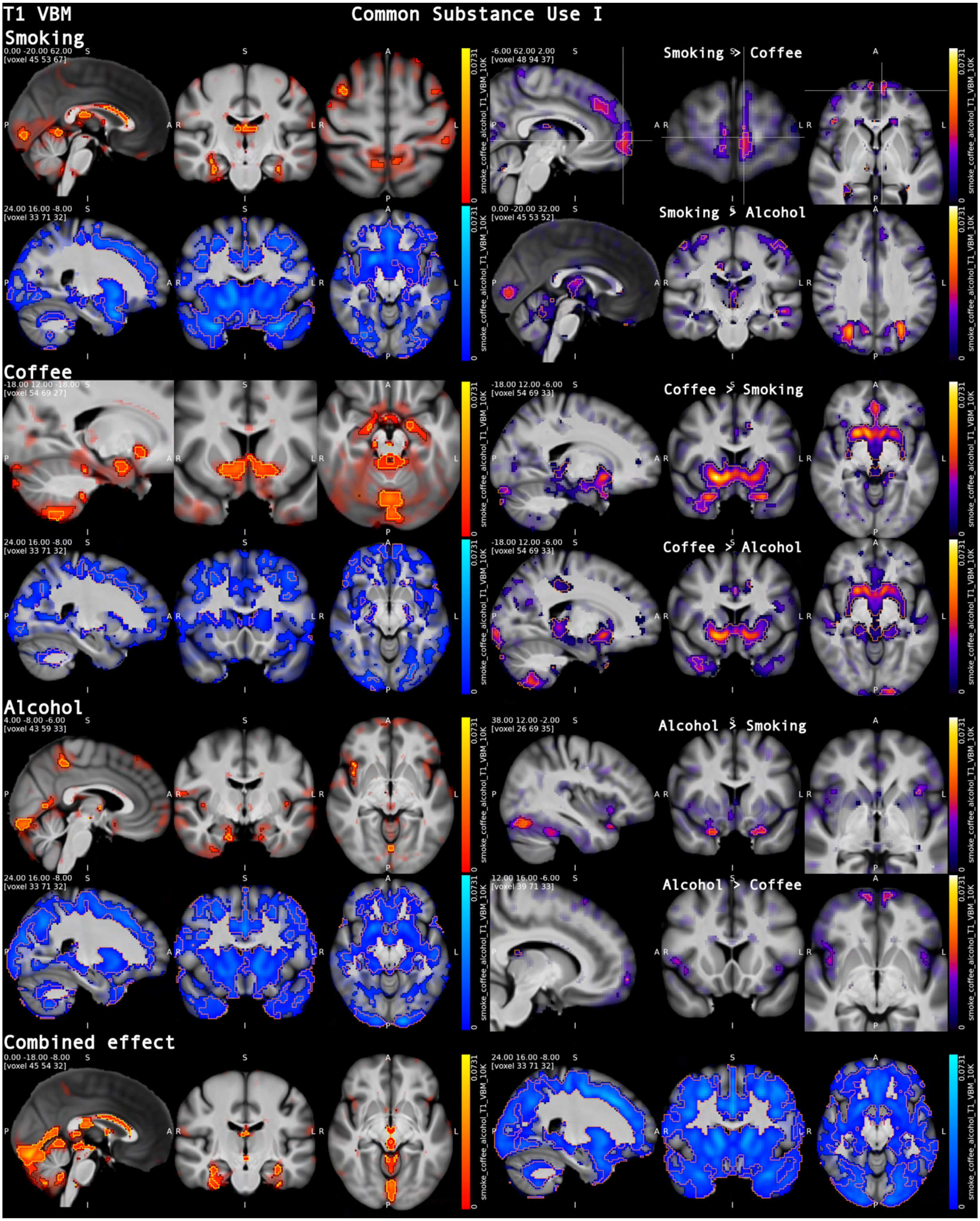


***Figure S5.1. Common substance use:*** *VBM. Extensive volume reductions were seen in association with alcohol, followed by smoking and to a lesser extent for coffee. Strong evidence was also found for localized and substance-dependent effects such as higher nucleus accumbens volume for coffee or anterior insula for alcohol.* <u>*Common*</u>

*visualization conventions (Figs. S3.1–3.12): β-maps thresholded at p_hFDR < 0.05 (black outline) overlaid on non-significant effects modulated by Z-statistic (surface views: non-significant effects shown by transparency, without Z-modulation), over the mean T1-weighted image. Positive associations are shown in red and negative associations in blue, contrast associations are displayed with a heatmap (NIH-fire); p_hFDR < 0.001 is outlined in yellow (positive and contrasts) or orange (negative), except on surface renderings, where p_hFDR < 0.001 is indicated in white. Group-mean resting-state network maps (Z-statistics) are thresholded at Z > 2.58 (approx. p < 0.01, two-tailed).*

**SWI T2*:**

**Smoking:** T2* was positively associated with strong evidence in fronto-parietal white matter, aligning with dmPFC and pre- and postcentral gyri, occipital lobe white matter, as well as within anterior entorhinal cortices and white matter aligning with more posterior parts of entorhinal cortex.

Strongest evidence for T2* increase was, however, seen outside the brain, surrounding the frontal lobe ($-\log_{10}(p)$_mapFDR > 10). While this may be interpreted as a sign of frontal lobe atrophy, caution is warranted when interpreting extracranial signal changes.

In direct contrasts, T2* was higher for smoking with strong evidence in small clusters in pre- and postcentral cortex and left hippocampus (mid-anterior) than for coffee consumption. Contrasting against alcohol revealed higher signal in bilateral LGN (presumably related to negative association of alcohol below).

Congruent with evidence from QSM, strongest evidence for T2* decrease was seen in bilateral putamen. Other negatively associated areas with particularly high significance were seen in bilateral anterior commissure, caudate, along bilateral central sulcus of the insula, in cerebrospinal fluid (CSF) cranial to superior colliculi. We, moreover, found strong evidence for cortical T2* decrease across frontal operculum and cerebellum, as well as white matter associated decrease in mesencephalon.

**Coffee:** There were no voxels with strong evidence for T2* increase with coffee. Strongest evidence for T2* ($-\log_{10}(p)$_mapFDR > 10) decrease was seen in left anterior commissure, bilateral habenula and hypothalamus. Other strong evidence areas included bilateral striatum, pallidum, medial substantia nigra, superior colliculi and posterior horns of lateral ventricles.

**Alcohol:** Alcohol revealed the strongest evidence for associated T2* signal increase across the brain with most significant areas located in bilateral mPFC, middle temporal gyrus, right forceps major, as well as surrounding infra- and supratentorial brain, possibly atrophy-related. Further areas of strong evidence were found across frontal grey and white matter, temporal cortices, parieto-occipital grey and white matter, amygdalae, anterior hippocampi, anterior pons and medulla oblongata. Direct contrasting revealed the signal alterations to be stronger than for smoking and coffee in most regions.

Like for smoking, alcohol was most strongly associated with T2* decrease in striatum, with other very significant areas ($-\log_{10}(p)$_mapFDR > 10) found in bilateral LGN, bilateral fasciculus retroflexus, right habenula, bilateral pulvinar, white matter next to VTA, pons, and cerebellar white matter. Other areas of strong evidence included subthalamic nuclei, inferior temporal gyri, and cerebellar cortices.

**DTI MD:**

**Smoking:** Smoking exhibited strong evidence for globally increased MD across supra- and infratentorial white matter. Regions of strongest evidence ($-\log_{10}(p)$_mapFDR > 10) were seen in corpus callosum, bilateral corticospinal tract and superior thalamic radiations, aligned with right precentral gyrus, forceps major, and bilateral thalamus.

Direct contrasting showed MD to be higher than for coffee in most of the brain, while changes in corticospinal and thalamic tracts were higher than in alcohol.

**Coffee:** While lobar white matter appeared mostly unaffected, coffee consumption showed strong evidence for MD increase in the corpus callosum, aligning with cortical patches in dlPFC, pre- and postcentral cortices, in paraventricular thalamus, and in cerebellar white matter. There was no strong evidence of reductions in MD.

**Alcohol:** Similar to smoking, alcohol consumption was related to extensive increase of MD across the brain. The increase was, however, primarily located in frontal and occipital lobes and less spread out than for smoking. Particularly high evidence for MD increase was found in the corpus callosum and fornix ($-\log_{10}(p)$_mapFDR > 15), as well as along lateral ventricles (including temporal horns), third and fourth ventricles.

We further found strong evidence for decreased MD in bilateral cingulum, putamen, hypothalamus, right lateral amygdala, bilateral acoustic radiations and right Heschl's gyrus, anterior pulvinar, mesencephalon, pons, and cerebellar white matter.

**DTI FA:**

**Smoking:** Bilateral uncinate fascicle and putamen exhibited strongest evidence of increased FA for smoking ($-\log_{10}(p)$_mapFDR > 10). Other strong evidence areas included bilateral hypothalamus, red nucleus, left substantia nigra, bilateral precentral gyrus and postcentral gyrus, thalamus, and white matter aligned with left entorhinal cortex. Uncinate fascicle FA was higher for smoking than coffee or alcohol with strong evidence. Reductions in FA were seen with strong evidence across all major white matter tracts, with particularly robust evidence ($-\log_{10}(p)$_mapFDR > 15) in bilateral optic radiations and posterior putaminal surface white matter.

**Coffee:** Moderate evidence was seen for FA increase in mediodorsal thalamus (parvocellular part). While coffee consumption exhibited the least extensive reductions in FA, particularly strong evidence was found for reduction corpus callosum, bilateral fornix, and between superior longitudinal fasciculi and cerebrospinal tracts.

**Alcohol:** Evidence was strongest ($-\log_{10}(p)$_mapFDR > 6) for alcohol-related FA increase along bilateral striatum, paraventricular thalamus, red nuclei, along cranial parts of PAG, left middle longitudinal fasciculus, parabrachial pons, and cerebellar white matter.

Complementary to MD findings, FA reduction exhibited strongest evidence for corpus callosum. Interestingly, this was also the case for the cingulum bundle, which was associated with decreased MD. Corpus callosum and cingulum exhibited strong evidence for FA reduction in direct comparison with coffee and smoking.

Further reductions were seen across major white matter tracts, albeit to a lesser extent than for smoking.

**DTI MO**

**Smoking:** Mode of anisotropy was positively related with smoking in internal capsule, bilateral uncinate fascicle, corticospinal tracts, superior longitudinal fasciculi, along pre- and postcentral cortices as well as bilateral dlPFC, and ventral posteromedial thalamus (VPM). Strongest evidence was found for genu of internal capsule and left uncinate fascicle ($-\log_{10}(p)$_mapFDR > 10).

Signal was decreased with strongest evidence (> 10) along dorsal superficial thalamic nucleus, bilateral putamen, cerebral pedunculus and right anterior commissure (temporal portion), and right claustrum. Further areas of strong evidence were found in corpus callosum, bilateral substantia innominata, right cingulum bundle and along internal surface of caudate.

**Coffee:** Strong evidence for positive association with DTI MO was found in anterior and posterior segments of corpus callosum, forceps major, and bilateral mediodorsal thalamus. Strong negative evidence was also found in the corpus callosum as well as crus cerebri and posterior pons.

**Alcohol:** Areas with highest evidence for positive shift of anisotropy mode ($-\log_{10}(p)$_mapFDR > 10) were found in corpus callosum including forceps minor and major, internal capsule, frontal aslant tract, mediodorsal thalamus, lateral pulvinar, and cerebellar white matter. More regions with strong evidence were found in fiber tracts across the brain.

As for coffee, negative alteration in diffusion mode was also found for corpus callosum, here extending into cingulum bundles. Further areas with particularly strong evidence (> 10) were found in temporal portions of bilateral anterior commissure, right uncinate fascicle, and aligned with dorsal putamen and PAG. The negative association with the cingulum bundle was again shown to be specific to alcohol consumption in direct contrasts.

**NODDI ICVF:**

**Smoking:** Particularly strong evidence ($-\log_{10}(p)$_mapFDR > 10) was found for increased neurite density along putaminal surfaces, ventral BNST/substantia innominata area, bilateral subthalamic region and medial substantia nigra.

Negative association with ICVF was complementary to findings in MD and FA, with strong evidence for extensive reductions across supra- and tentorial white matter, exhibiting particularly robust evidence (> 10) in pre-, para- and postcentral white matter, cortico-spinal tract and internal capsule as well as bilateral thalamus. Most of these associations were stronger than for alcohol and coffee in direct comparison.

**Coffee:** Strongest evidence of neurite density increase ($-\log_{10}(p)$_mapFDR > 10) for coffee was found along left caudate, medial forebrain bundle, and in bilateral hypothalamus, followed by further areas of strong evidence in bilateral globus pallidus, along lateral amygdalae and amygdalo-striatal transition area, and in bilateral crus cerebri as well as cerebellar vermis. Strong evidence associations for decreased density were found in left precentral cortex.

**Alcohol:** ICVF along putamen was increased with substantially strong evidence (-$\log_{10}(p)$_mapFDR > 10) across putamen, mid- and posterior corpus callosum, red nuclei, white matter adjacent to VTA, dorsal to locus coeruleus and in cerebellar white matter. Additional areas of strong evidence were found in bilateral pulvinar and anterior mediodorsal nuclei of thalamus.

Negative associations with ICVF mirrored findings in FA and MD, with extensive regions of reduction in frontal and occipital lobes and particularly strong evidence for reductions in anterior corpus callosum and forceps minor but also along straight gyri and dmPFC.

**NODDI OD:**

**Smoking:** Evidence for increased orientation dispersion was highest (-$\log_{10}(p)$_mapFDR > 10) in bihemispheric substantia nigra, around bilateral striatum and aligned with substantia innominata. Other areas of strong evidence included orbitofrontal fibers, the crossing of bilateral frontal aslant tract with superior longitudinal fasciculus, left cingulum bundle, white matter lateral to bihemispheric LGN (possibly stria terminalis), and fibers along left dlPFC, and bilateral pre- and postcentral cortices. Evidence was, moreover, strong for higher signal than alcohol in bilateral substantia nigra, and higher than coffee in the left substantia nigra.

Consistent with findings from other diffusion metrics, robust evidence was found for decrease in orientation dispersion of bilateral uncinate fascicle (-$\log_{10}(p)$_mapFDR > 20), also in direct comparison with coffee and alcohol consumption. Further areas of strong evidence were temporal portion of anterior commissure, fibers along entorhinal cortices, corpus callosum, anterior and superior thalamic radiations, corticospinal tracts, internal capsules, bilateral thalamus, superior longitudinal fascicles, and in proximity of bilateral locus coeruleus.

**Coffee:** Coffee consumption was also associated most robustly with OD in a slightly more medial region of substantia nigra (-$\log_{10}(p)$_mapFDR > 8). Further regions of strong evidence included callosal fibers, fibers lateral to cingulum bundle, internal capsule bordering on nucleus accumbens, anterior commissure, bilateral fornix, fibers along pre- and postcentral cortices, and along bilateral longitudinal white matter tracts.

Strong evidence for OD decrease was found in bilateral thalamus (centromedian / mediodorsal).

**Alcohol:** OD increase with alcohol was most strongly supported (-$\log_{10}(p)$_mapFDR > 10) for bilateral internal capsule along caudate nucleus, corpus callosum, cingulum bundle, along lateral putaminal surfaces, fornix and anterior commissure, left pre- and postcentral cortex, in a longitudinal bundle crossing right corticospinal tract, right and mesencephalic white matter between bilateral red nuclei and PAG. Further mostly longitudinal cerebral tracts as well as cerebellar white matter tracts were found to associate with alcohol with strong evidence.

Most robust evidence for OD decrease (> 10) was found in forceps minor and apical of forceps major, frontal aslant tract (FAT), paraventricular hypothalamus, amygdalo-striatal transition area, right superior PAG, posterior pons along locus coeruleus region, and cerebellar white matter. Further areas of strong evidence were mostly found along longitudinal association tracts but also in bilateral thalamus, left pallidum, and along entorhinal cortices.

**Warpfield Jacobian**:

**Smoking:** Strong evidence was found for local volume expansion in bilateral internal capsule surrounding and including lateral parts of caudate nucleus for smoking (note: in warpfield Jacobian maps, higher values indicate that a region had to be compressed more strongly to match the MNI group template, and therefore reflect larger native volume before normalization), as well as between callosal sulci, in bilateral dmPFC, parahippocampal gyrus, anterior insular cortex (bordering on central sulcus), left postcentral cortex, bilateral postcentral sulcus, right paracentral lobule, posterior horns of lateral ventricles, and cerebellum (VIII/IX and left I-IV). Insular expansion was also higher in direct comparison with alcohol and coffee.

Negative association analysis exhibited edge effects surrounding frontal and occipital lobes, which are probably atrophy related but should be interpreted with caution. However, clearly intracortical areas with volume contraction included bilateral vm-, dm-, and dlPFC  lateral orbitofrontal cortices, and parieto-occipital transition area. Further areas with strong evidence for contraction included corpus callosum (genu), basal forebrain area extending into anterior striatal regions and amygdalae, bilateral thalamus, mesencephalon, cerebellar peduncles and white matter, and medulla oblongata.

**Coffee:** Coffee consumption was associated with local expansion in fornix and anterior lateral ventricle, right supplementary motor area (lateral part), transition from fornix to corpora mammillaria, left postcentral cortex and superior parietal lobule, bilateral region including posterior caudate and lateral ventricles, posterior horns of lateral ventricle, cerebellar peduncles and cerebellum (vermis, bilateral VIIIa, VIIb, X).

Coffee exhibited more restricted areas of contraction. These were limited to bilateral frontal and right fronto-parietal white matter, corpus callosum (genu) including ACC, bilateral pallidum extending into thalamus and posterior internal capsule, and right occipital white matter.

**Alcohol:** Alcohol exhibited strongest evidence ($-\log_{10}(p)$_mapFDR > 10) for expansion of anterior and posterior horns of lateral ventricles. The expanded region also included large parts of caudate nucleus. Further areas roughly corresponded to bilateral cingulate cortex and callosal sulcus, anterior hippocampus, entorhinal cortices, postcentral cortex, sulcus and superior parietal lobules, third ventricles, subarachnoid cisterns, and venous sinuses.

As for smoking, strongest evidence was found for edge effects, this time surrounding most of the brain, and likely atrophy-related. Cortical contraction was seen across frontal, parietal and occipital lobes, with temporal lobes being less affected, and in cerebellum. Furthermore, strong evidence was seen for contraction across subcortical areas, global white matter including corpus callosum, in mesencephalon and medulla oblongata.

Direct comparison revealed these global effects to be stronger in alcohol than in smoking and coffee.

**RSN d25-2 (DMN):**

**Smoking / Coffee:** There were no strong evidence effects for smoking and coffee.

**Alcohol:** Strong-evidence positive associations, comprising increased coupling and decreased anticorrelation changes with respect to the group-level network, were found in bilateral m-/ and dlPFC, orbitofrontal cortices, temporal poles, middle temporal gyri, left postcentral gyrus, right supramarginal gyrus, bilateral precuneus and superior parietal lobule; right mid-hippocampus, thalamus and cerebellum.

**RSN d25-6 (FPN-1):**

**Smoking:** No strong evidence alterations were found within the fronto-parietal component RSN d25-6.

**Coffee:** Strong evidence was found for reduced coupling (negative association) in left temporo-parietal junction with RSN d25-6.

**Alcohol:** Strong evidence was found for positive associations, comprising primarily increased coupling changes, with alcohol in bilateral vlPFC, supramarginal gyri (and bordering on angular gyri), left middle temporal gyrus, bilateral precuneus and occipital gyri. Increased anticorrelation (negative associations) was strongly supported in left ACC, left mPFC, right vlPFC and frontal operculum, bilateral precuneus, and cerebellum.

**RSN d25-9 (SN):**

**Smoking:** Reduced anticorrelation of left precentral cortex within the salience-like RSN_d25_9 network was supported strongly for smoking (positive association).

**Coffee:** No strong evidence alterations were found.

**Alcohol:** Positive associations (strong evidence), signifying increased coupling with respect to the mean image, were seen in right precentral gyrus, SMA, parieto-frontal operculum, posterior section of bilateral middle temporal gyri, and right superior parietal lobule. Decreased anticorrelation with respect to the mean, was seen in precuneus along the parieto-occipital sulcus (strong-evidence positive association).

**RSN d25-11 (FPN-2):**

**Smoking:** Increased anticorrelation (negative association) of left middle temporal gyrus was strongly supported in a second fronto-parietal network.

**Coffee:** No strong evidence alterations were found.

**Alcohol:** Coupling within the network was increased (positive associations) most robustly in left dlPFC bordering on vlPFC ($-\log_{10}(p)$_mapFDR > 10). Other strong evidence areas, all signifying coupling increase relative to the mean, included bilateral frontal pole, vl-/dlPFC, left vmPFC, bilateral anterior insula, right SMA, bilateral precuneus, left temporal cortices, bilateral angular gyrus, and occipital gyri; left putamen and bilateral cerebellum.

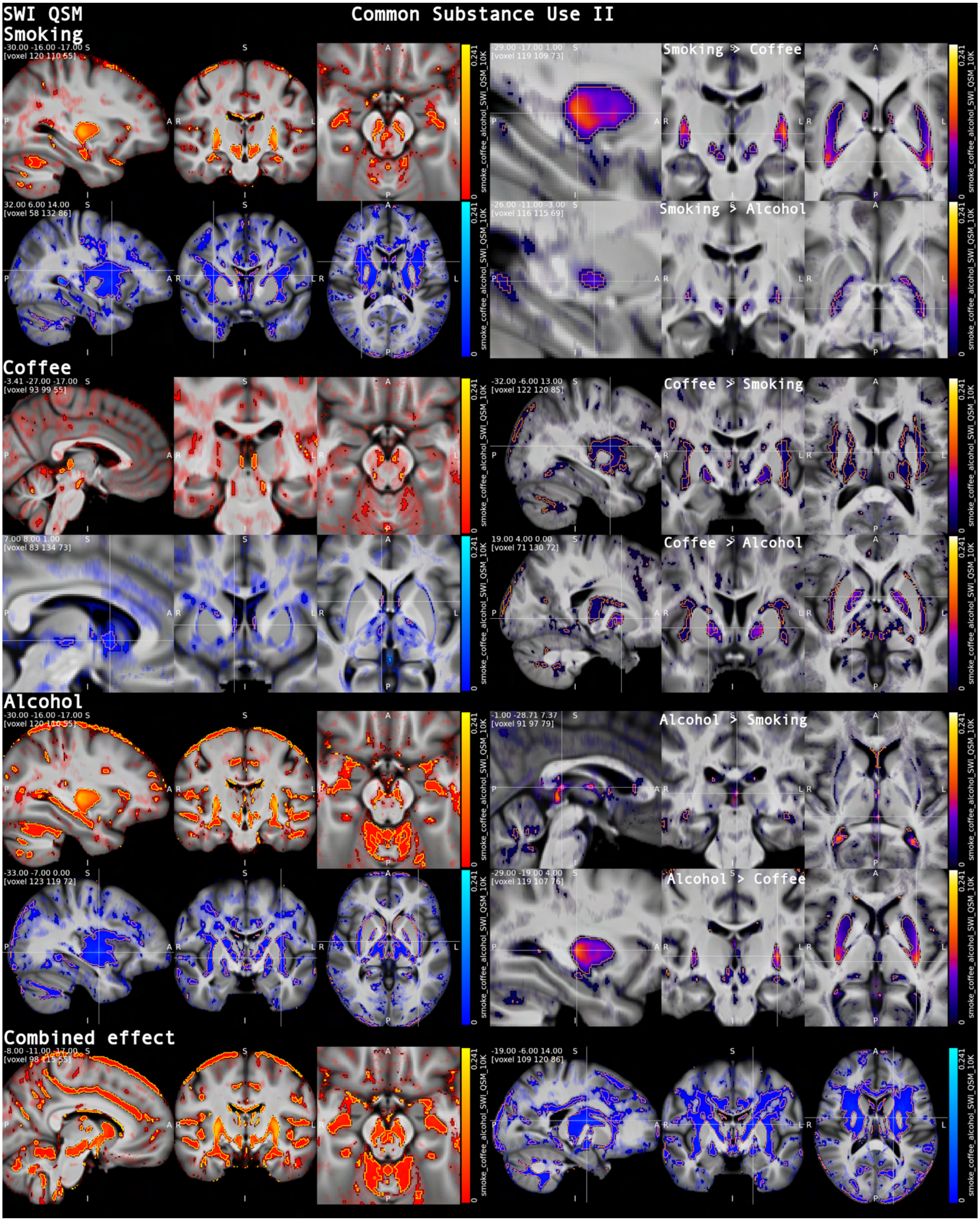


***Figure S5.2. Common substance use: QSM***. *Smoking, alcohol and to a lesser extent coffee intake exhibited robust evidence for QSM signal increase in striatum and nigral pathways consistent with iron accumulation (peak for coffee in habenula). WM QSM, particularly, in internal and external capsule exhibited decrease. (Visualization see Fig. S3.1.)*

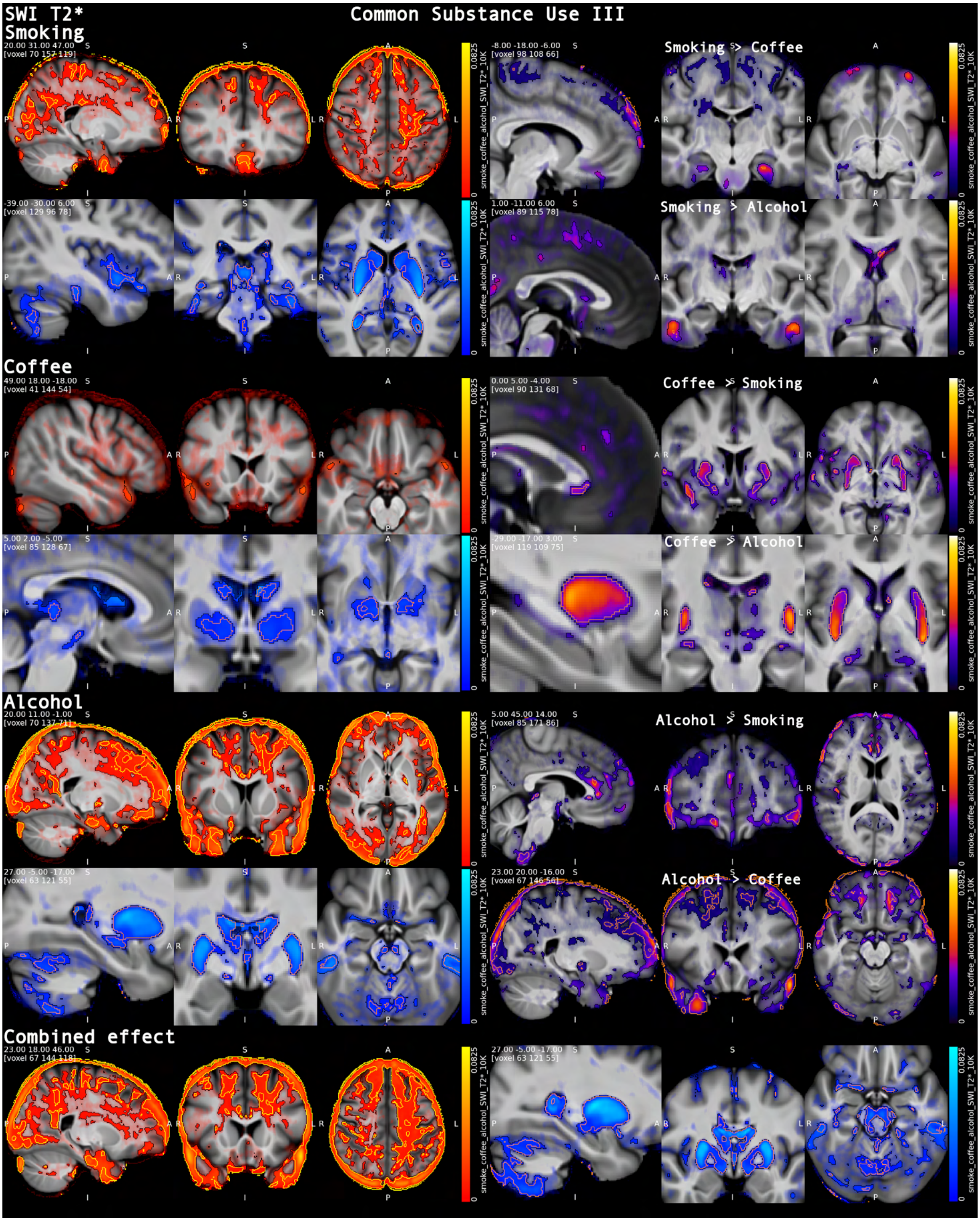


***Figure S5.3. Common substance use: T2****. *Consistent with QSM and iron accumulation, T2* decreased with substance use in striatum. Alcohol and smoking showed global WM/GM T2* elevations with distinct local propensities. Possibly atrophy-related edge effects should be interpreted with caution. (Vis. conv.: Fig. S3.1.)*

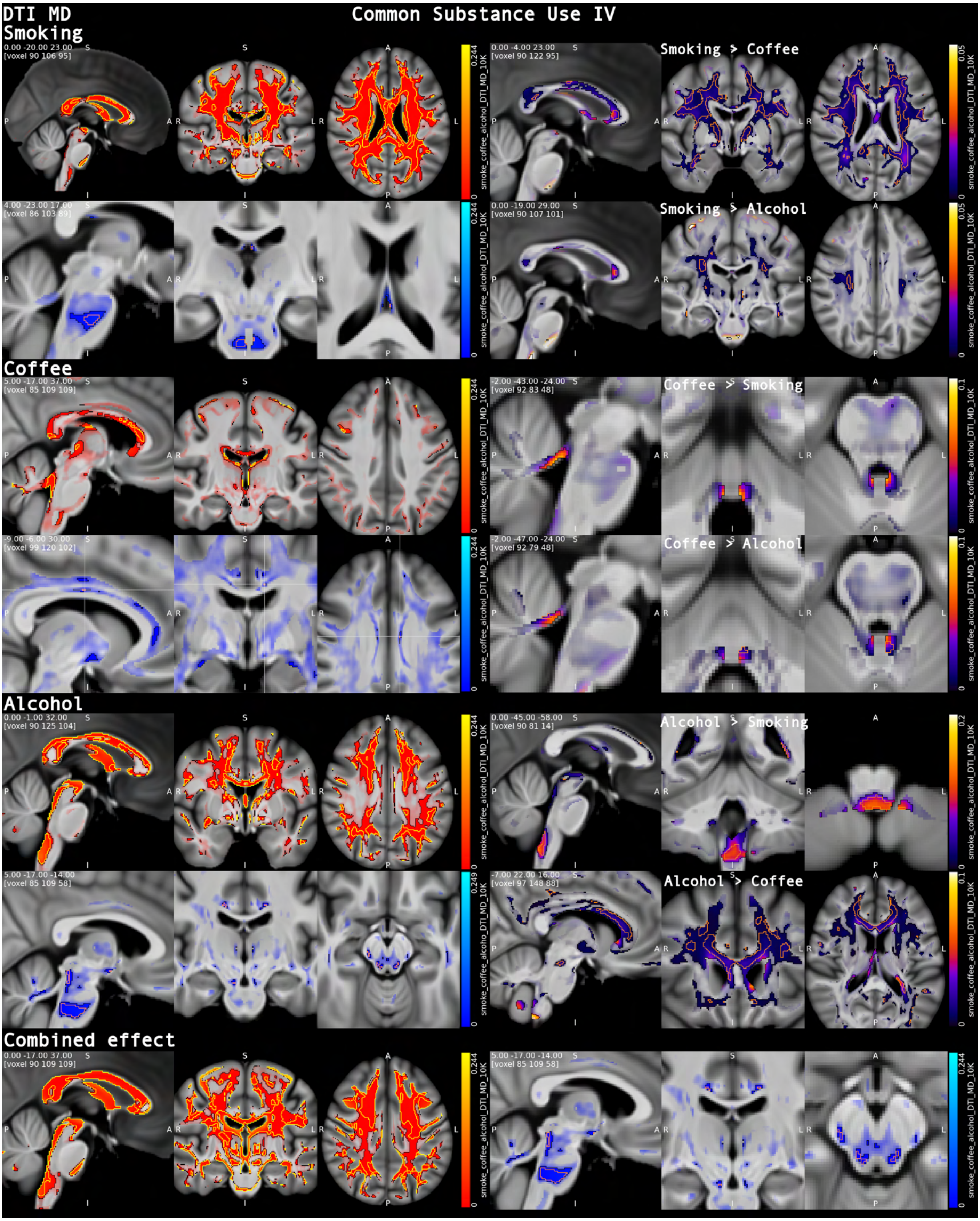


***Figure S5.4. Common substance use: DTI MD**. Global increase of MD was seen for smoking, followed by alcohol. Effects for coffee were localized, e.g., in corpus callosum. Combined effect analysis highlighted decrease of MD in uncinate fascicles, adjacent to amygdalae, and periaqueductal grey (PAG). (Visualization convention: Fig. S3.1.)*

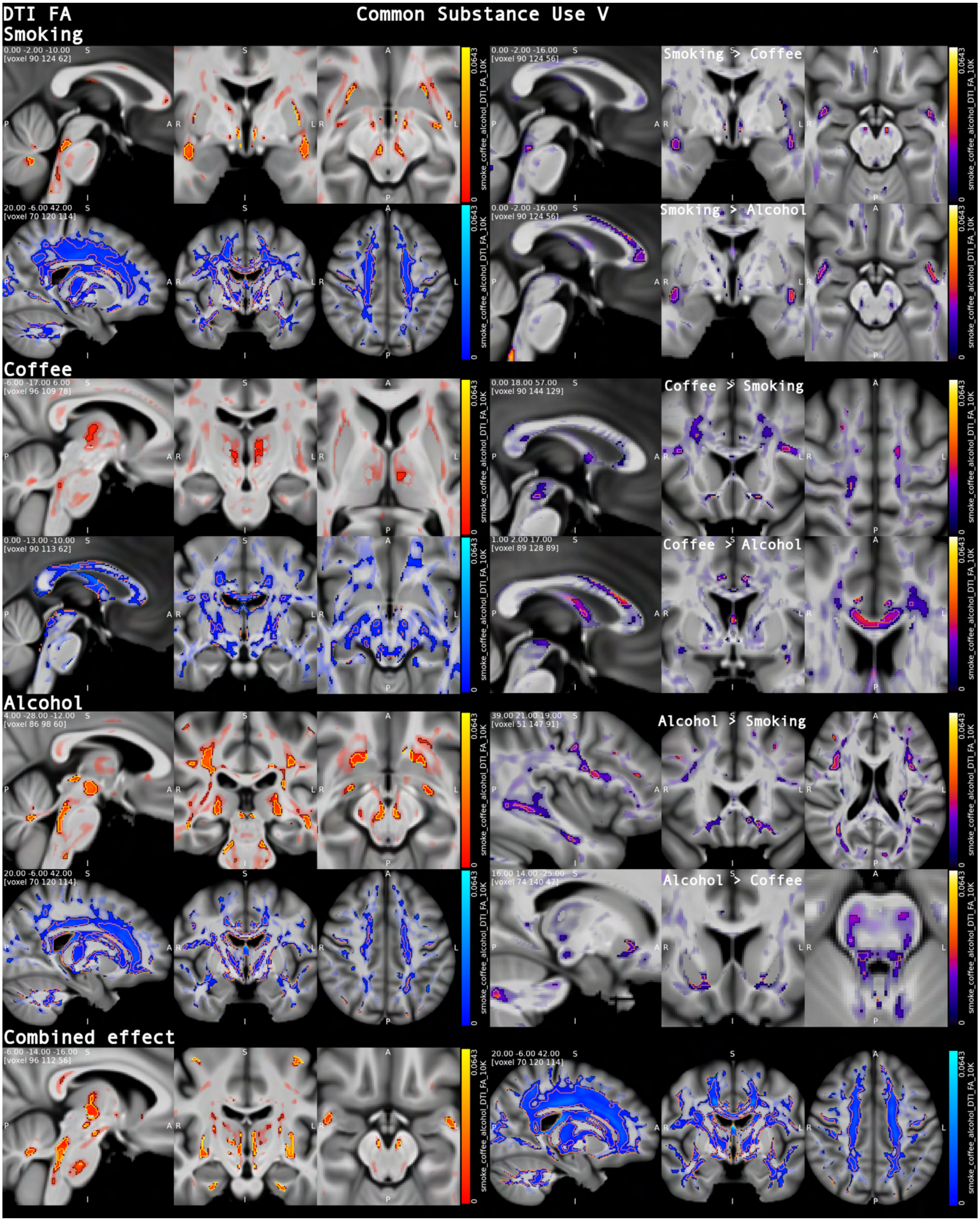


***Figure S5.5. Common substance use: DTI FA***. *FA analysis confirmed widespread microstructural deterioration with nicotine and alcohol consumption, with again more limited effects for coffee. Uncinate fascicle FA was particularly increased for smoking as evidenced by direct contrast analyses. (Visualization convention: Fig. S3.1.)*

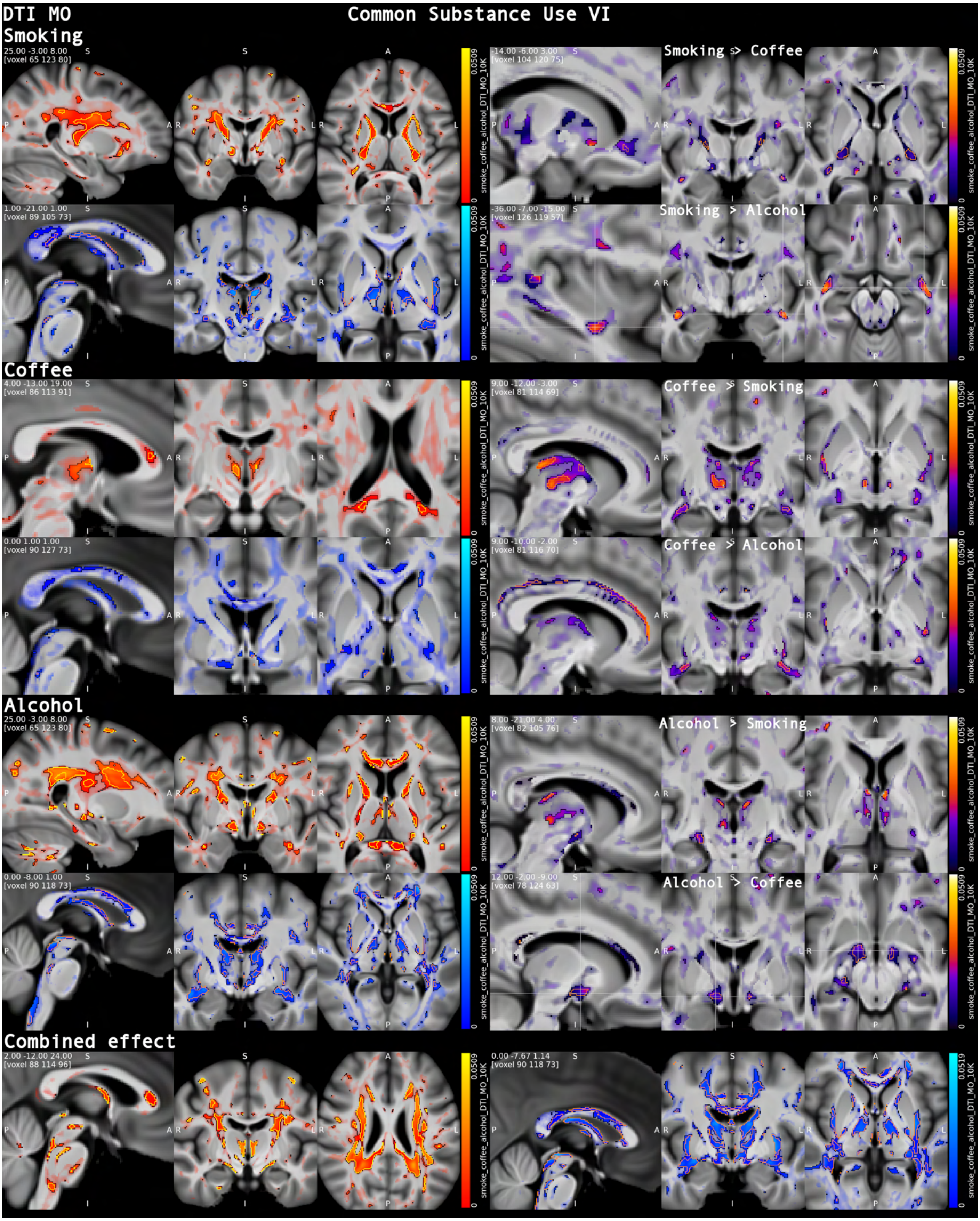


***Figure S5.6. Common substance use: DTI MO****. Changes in anisotropy mode (↑: prolate-shaped tensor, i.e., principal eigenvalue λ1 dominates over λ2 and λ3, ↓: oblate-shaped, λ1 ≈ λ2 ≫ λ3) were widespread for smoking and alcohol. Coffee was primarily related to changes in thalamus, callosum and anterior commissure. (Vis. conv.: Fig. S3.1.)*

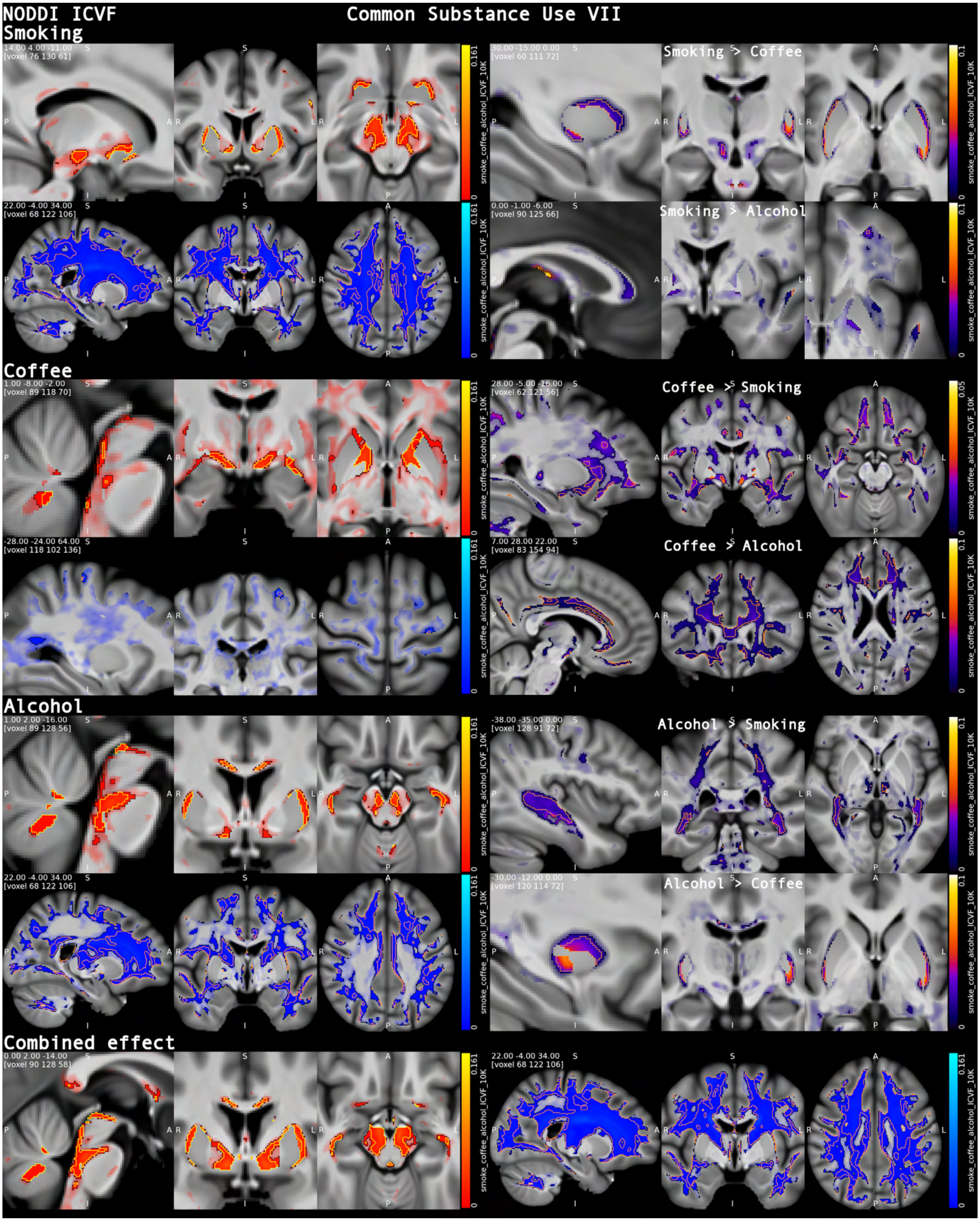


***Figure S5.7. Common substance use: NODDI ICVF****.* *Consistent with DTI findings, alcohol and smoking exhibited general patterns of reduced WM neurite density with associations with smoking being more extensive. Increases were primarily localized around putamen, in basal forebrain area and mesencephalon. (Visualization convention: Fig. S3.1.)*

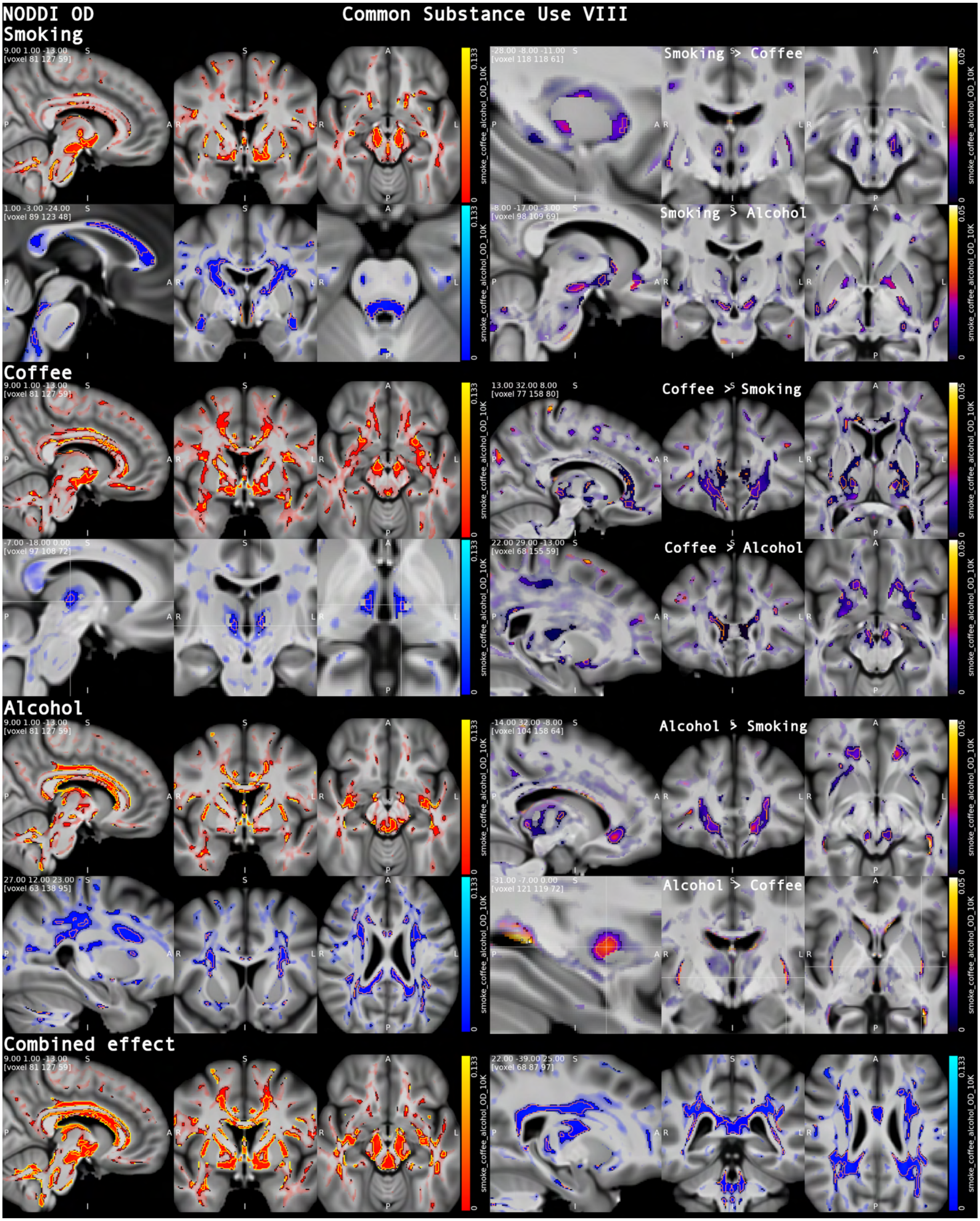


*Figure S5.8. **Common substance use: NODDI OD**. OD increases with were concentrated in nigrostriatal/basal-forebrain and major association pathways, whereas OD decreases were most consistent in uncinate fascicles (smoking) and in callosal/frontal and thalamic/brainstem regions (alcohol; thalamus for coffee). (Vis. conv.: Fig. S3.1.)*

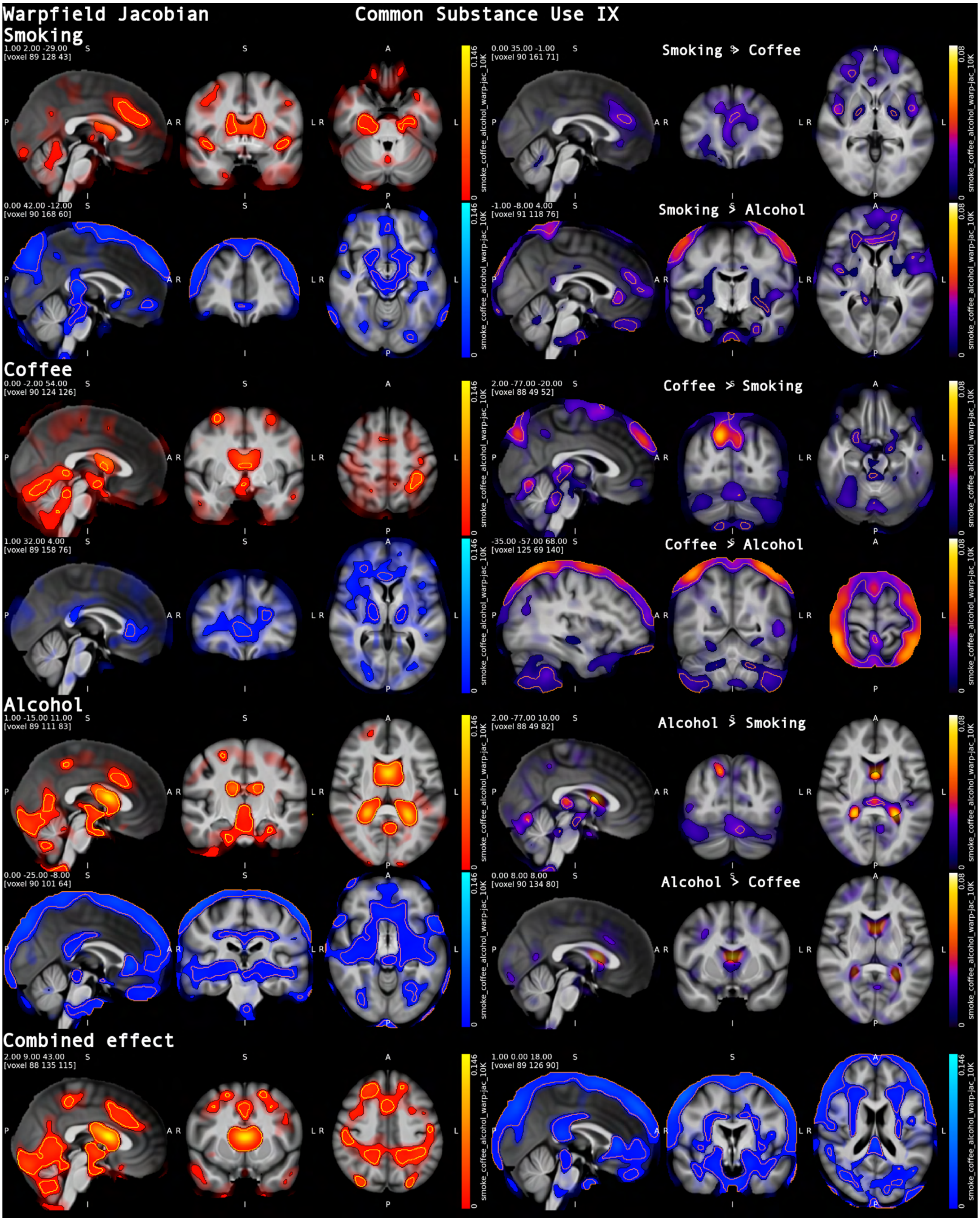


***Figure S5.9. Common substance use: warpfield Jacobian***. *Smoking, alcohol and coffee were associated with distinct patterns of local volume expansion (+) and contraction (-). Alcohol, followed by smoking, related to extensive expansion of CSF space and contraction in WM and GM. Interpret edge effects with caution. (Vis. conv.: Fig. S3.1.)*

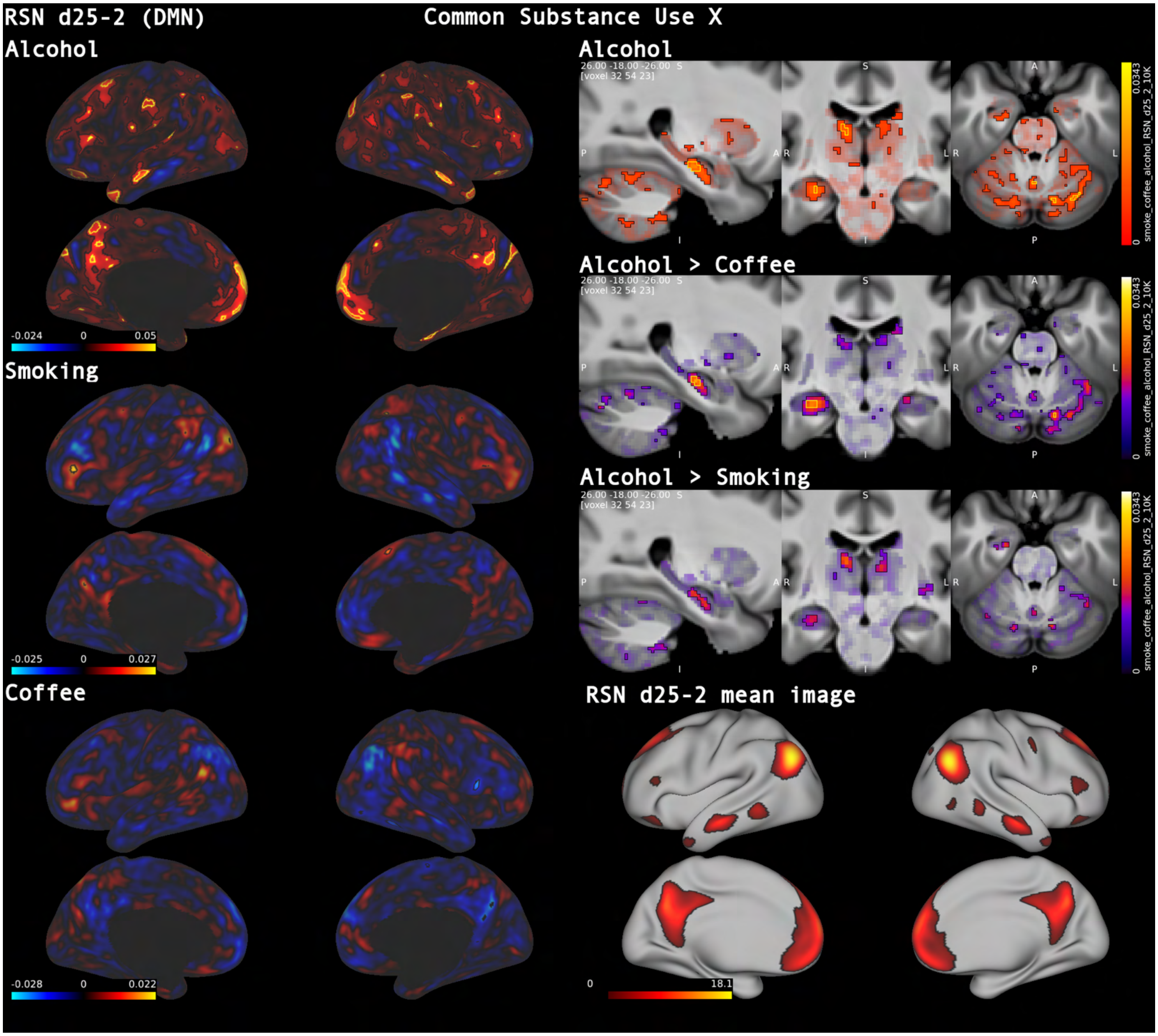


***Figure S5.10. Common substance use: RSN d25-2 (DMN).*** *Alcohol consumption was related to increased coupling in core regions of the default mode network but also in many distributed patches across cortex and subcortex, consistent with reduced network specificity (dedifferentiation). Small regions in left dlPFC and right dmPFC showed increased coupling in smoking with further small effects in right precuneus and frontal operculum for coffee (Visualization convention: Fig. S3.1.).*

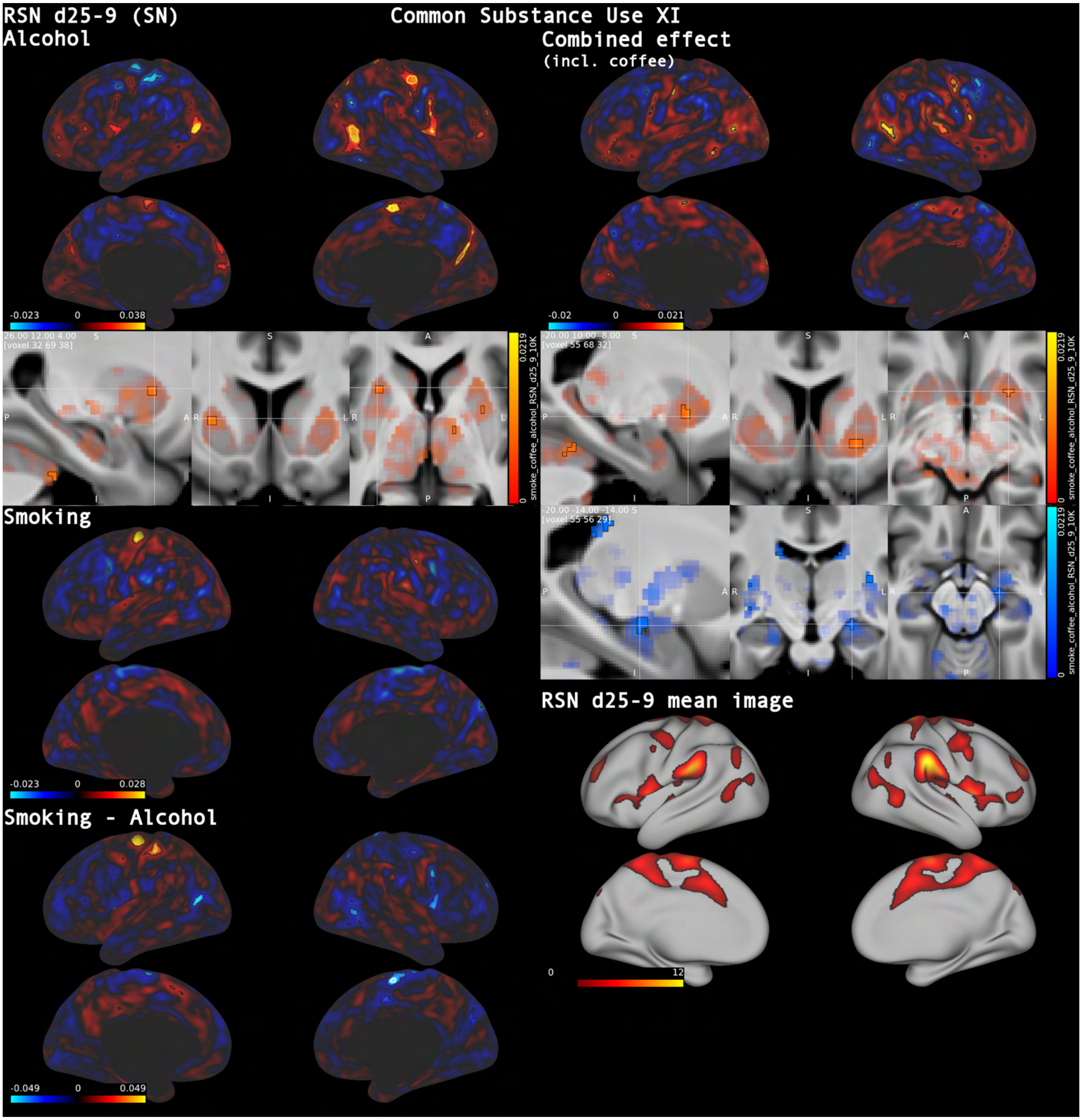


***Figure S5.11. Common substance use: RSN d25-9 (SN).*** *A salience-like network again exhibited more extensive alterations with alcohol including striatal involvement (also seen as combined effect), while smoking was associated with increased coupling of left motor cortex. (Visualization convention: Fig. S3.1.)*

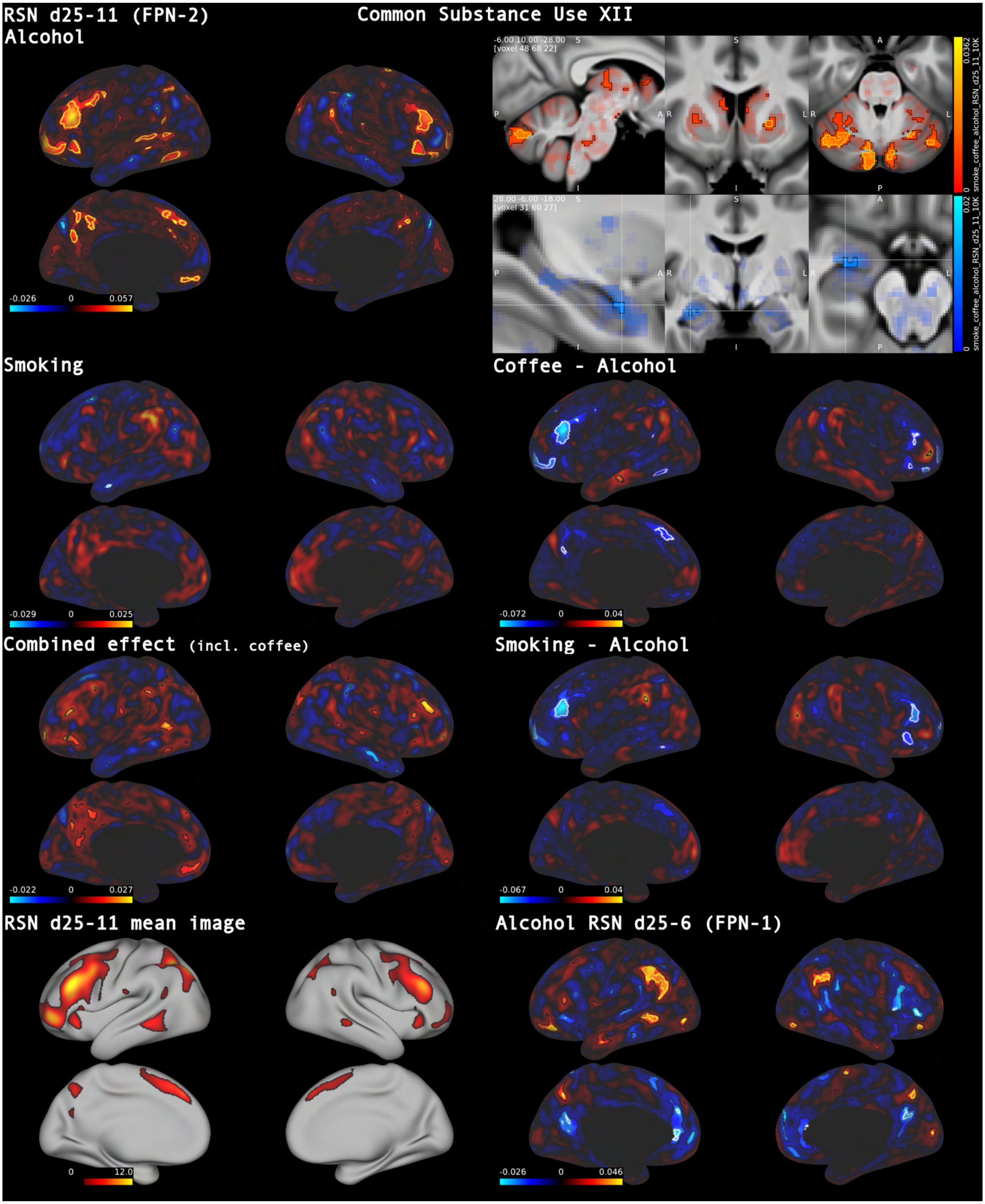


***Figure S5.12. Common substance use: RSN d25-11 (FPN-2).*** *Alcohol related with increased network coupling and added recruitment of cortical areas (e.g., vmPFC and superior temporal gyrus) and subcortex. A second fronto-parietal network (FPN-1) exhibited increased gain of positive and anti-correlated parts. (Vis. conv.: Fig. S3.1.)*

**Discussion of Supplementary Experimental Results**

**Experiment 1:** Lifetime trauma load, nigrostriatal iron and multimodal structural-functional alterations in the general population

In a first multimodal single-regressor experiment, we investigated associations between cumulative traumatic experiences and the brain (Figs. 4 and S1.1 – S1.2).

Lifetime trauma was associated with increased QSM signal in the substantia nigra with strong evidence, compatible with higher iron content. Strikingly, posttraumatic stress disorder (PTSD) has been linked to an increased risk of Parkinson's disease (PD) with reported hazard ratios ranging from 1.48 to 3.46 (Barer et al., 2022; Chan et al., 2017). While chronic restraint stress has been shown to trigger dopaminergic loss in substantia nigra in rats (Sugama et al., 2016), to the best of our knowledge, a direct anatomical link in humans has not previously been reported. Bilateral putamen, a downstream region of pathology for PD, was another area with strong evidence for increased QSM. A rat model of PTSD exhibited neuronal injury in the striatum caused by iron deposition (Zhao et al., 2016). Intriguingly, trauma was also associated with decreased QSM signal (e.g., compatible with more myelin or less iron) in a cerebellar thalamic region (VLpv), which has been related to tremor in PD due to lack of inhibition via dopamine (Dirkx et al., 2017).

Strong evidence indicated global white matter signal shifts across diffusion sub-modalities and T2* with increasing trauma load, compatible, for example, with a degenerative process. Specifically, trauma load was associated with infra- and supratentorial increases in MD and decreases in FA and neurite density (ICVF).

White matter changes have been reported in relation to PTSD and stress across many studies (Dennis et al., 2021; Li et al., 2016). A meta-analysis in adults and children with the disorder reported FA reductions in the corpus callosum, cingulate and frontal regions but could not pinpoint specific areas in a coordinate-based analysis (Siehl et al., 2018). In a multi-cohort study of 1,426 individuals with PTSD and controls Dennis et al. (2021) reported lower FA in the tapetum region of the corpus callosum. An early structural study in a small sample of PTSD patients and controls also reported generalized white matter atrophy (Villarreal et al., 2002), which would be consistent with our findings here.

Dysregulation of the hypothalamic-pituitary-adrenal (HPA) axis has been further implicated in PTSD (Raise-Abdullahi et al., 2023). We found strong evidence for microstructural alterations in hypothalamus with increased fiber orientation dispersion (OD) co-localized with decreased QSM signal, which can be a sign of increased myelination. Together these changes are consistent with remodeling of hypothalamic connectivity, which has been shown to be predictive of prolonged stress (Jensen et al., 2024).

In our analysis of voxel-based morphometry (VBM) maps, we found strong evidence for volume reduction in bilateral amygdalo-hippocampal complex with peak evidence for the right posterior amygdala.

While hippocampal volume loss has been reported for PTSD across many studies (Logue et al., 2018), results regarding amygdala have been mixed (review: Ben-Zion et al., 2024). Amygdala volumes have, for instance, been reported to be enlarged in war-veterans with PTSD relative to veterans without PTSD (Kuo et al., 2012). However, greater combat exposure was associated with smaller amygdala volume in the same study. Another study from the same year, also conducted in veterans with and without PTSD, found smaller amygdala volumes in PTSD and no association with trauma load (Morey et al., 2012). Notably, smaller amygdala volume has been associated with PTSD severity in veterans and traumatized youths (Homan et al., 2019; Ousdal et al., 2020).

A variety of methods have been used to measure amygdala volume, which, together with the fact that amygdala consists of many subnuclei, may well contribute to these inconsistencies. Here, we observed peak evidence localized in the in right posterior parts of amygdala (Z = -7.1). For comparison, right amygdala IDP (UKB ID: 25889) showed much lower evidence (Z = -4.2).

Retinotopic visual cortex VBM associations with lifetime trauma are discussed in the main text.

In an emotion task, we found increased activation in anterior hippocampus and the default mode network (DMN) in response to fearful and angry faces over neutral shapes (faces–shapes contrast) with higher traumatic experiences load. In a previous experimental study, anterior hippocampal fMRI response was related to level of threat in an approach-avoidance task but better explained by avoidance behavior (Abivardi et al., 2020), a core symptom of PTSD (American Psychiatric Association, 2013; World Health Organization, 2022). Remarkably, fearful facial expression have been shown to provoke approach behavior in healthy subjects (Hammer & Marsh, 2015), while the default mode network has been found to deactivate in response to negative faces (Sreenivas et al., 2012).

We further found strong evidence of altered resting-state networks including reduced coupling of precuneus within DMN, which is consistent with findings in women with early-life trauma-related PTSD and veterans (Bluhm et al., 2009; Miller et al., 2017).

It should be noted that findings from clinical PTSD samples may not fully generalize to trauma-exposed individuals with subclinical symptoms or to the broader general population. Because our analyses focus on exposure rather than clinical symptomatology, the observed brain effects are also likely to capture mechanisms of resilience. In fact, both peri- and posttraumatic ventromedial prefrontal cortex (vmPFC) volume, which has been reported to relate to resilience across several studies (Roeckner et al., 2021), and an anterior region in dorsolateral prefrontal cortex (dlPFC) identified to be expanded in trauma-exposed healthy controls vs. PTSD patients (Zilcha-Mano et al., 2022), exhibited greater grey matter volume here.

We further note that our cumulative trauma score does not differentiate between childhood and adult traumatic experiences.

**Experiment 2:** Dissociable brain signatures of anxiety and depressive symptom dimensions

In a second experiment, we investigated differential effects of depressive and anxiety symptoms in the general population (Figs. 4 and S2.1 – S2.6). Specifically, we compared the first principal component of recent symptom load from depression- and anxiety-specific online follow-up questionnaires. Of note, we did not include sleep disturbances in the analysis, which are bidirectionally related to both anxiety and depression.

Strong evidence linked depressive symptoms to volume loss in bilateral amygdalo-hippocampal complex with peak loss in right centromedial amygdala, whereas anxiety was associated with increased volume in anterior hippocampus. Directly contrasting anxiety–depressive symptom load further strongly supported larger left anterior hippocampus volume for anxiety.

Hippocampal volume loss has been consistently shown for major depressive disorder (Roddy et al., 2019; Wise et al., 2017), while results for amygdala involvement, similar to PTSD, have been mixed (Roddy et al., 2021). Our findings with respect to anxiety symptom load and hippocampus resonate with Rusch et al.

(2001), which showed that hippocampal volumes were positively associated with trait anxiety in both healthy and depressed subjects.

Strong evidence was found for an association of depressive symptom load with increased volume in posterior putamen, while anxiety symptom load supported increased volume in anterior caudate. Striatal VBM results are discussed in the main text.

A recent study examining amygdala responses to negative faces in depression in UKB found no association (Tamm et al., 2022). Specifically, the study investigated the relationship between current depressive symptoms and median fMRI effect for faces–shapes contrast in a group-defined amygdala-activation mask (UKB ID: 25052) for 28,638 individuals. The summed depressive index (from the imaging date) included three questions related to core depressive symptoms, and one anxiety-related item.

We revisited the question of missing amygdala activation in the Hariri emotion task (Hariri et al., 2002), albeit with the anxiety and depressive symptom principal components, and across all three contrast z-statistic tfMRI sub-modalities.

We found increased amygdala reactivity for the first depressive symptom principal component in right amygdala to both shapes and faces, while the principal component for anxiety related negatively to task amygdala activation for both. Correspondingly, the contrast of depressive–anxiety symptoms was positively associated with bilateral basolateral amygdala activation to shapes, and bilateral centromedial amygdala activation to faces. The combined effect of anxiety and depressive components on amygdala fMRI response was negative in response in both subtasks.

Not unexpectedly, as we found increased reactivity for both shapes and emotional faces in higher depressive symptom load, we did not find an association with the faces–shapes contrast. However, using the higher sensitivity 1K-supervoxel analysis, we found a positive association between greater depression–anxiety symptom difference and higher left centromedial amygdala reactivity to faces over shapes.

Diffusion-weighted analyses exhibited stronger signs of white matter deterioration associated with depressive symptoms than with anxiety score, with supra- and infratentorial decreases in ICVF and FA as well as accompanying increase in MD. Similar to the effect of cumulative trauma, depressive symptoms were also associated with increased orientation dispersion in hypothalamus. Peak evidence for the combined positive effect of anxiety and depressive symptom load on orientation dispersion was seen in the right medial forebrain bundle, a known target of deep brain stimulation in treatment-resistant depression (Fenoy et al., 2022).

**Experiment 3:** An *EPHA3* variant shaping interhemispheric and temporal connectivity

In a third main experiment, and first of two genetic studies, we analyzed the effect of *EPHA3* SNP rs987748 (A:C) on the brain (Figs. 5 and S3.1 – 3.5). Ephrin signaling plays core roles in axon guidance, cell migration (Kania & Klein, 2016), and tissue patterning (Janis et al., 1999); *EPHA3* gene mutations have been implicated in glioblastoma (Day et al., 2013) and autism (Casey et al., 2012).

In a previous study, we found that the minor allele of rs987748 was associated with occipital lobe resting-state functional connectivity with the left hemisphere (ICA100 edge 383), as well as with rfMRI connectivity

ICA-feature 3, a derived summary measure of whole-brain functional connectivity (Smith et al., 2021; see Supplementary Figure S18.3 for the spatial pattern of this component). A weaker association of the major allele with baseline fluid intelligence is reported in Schoeler et al. (2025; Supplementary Data 3).

Across diffusion modalities and QSM a common picture emerged with some of the most robust evidence. The minor allele was associated with decreased microstructural integrity of anterior commissure (e.g., peak evidence for increased fiber dispersion of $-\log_{10}(p)$_mapFDR > 100) and temporal lobe connectivity via anterior commissure/uncinate fascicle. By contrast, we found increased integrity of the mid corpus callosum and of long-range temporo-parietal and parieto-occipital fiber tracts. We commonly saw positive and negative associations in the same tracts but with slightly shifted location, compatible with fine-grained control of tract geometry/positioning and local fiber architecture. These microstructural effects were combined with strong evidence for volumetric differences associated with the minor variant, specifically, reduced temporal lobe and dorsal thalamus volumes, and increased volumes in fronto-parieto-occipital lobes. Functional analyses exhibited striking left-lateralization in face perception tasks across cortical and subcortical regions, while tests in resting state networks revealed distinct anatomical shifts in activity. Left-lateralization of a face-perception network has previously been described in left-handed men (Thome et al., 2022). A different SNP near the *EPHA3* gene has recently been related to hemispheric differences in language network connectivity in UKB (Amelink et al., 2024). Interestingly, high *EphA3* expression has been found in the developing dorsal thalamus of rats (Mackarehtschian et al., 1999).

*EphA3* is expressed in anterior commissure and corpus callosum of mice (Kudo et al., 2005). While EphA3 has been implicated in callosal axon segregation (Nishikimi et al., 2011) and *EphA4* is required for anterior commissure formation (Dottori et al., 1998), evidence for a link between disrupted anterior commissure found in *Olig2* knockout mice and *EphA3* has previously remained inconclusive (Gotoh et al., 2023).

**Experiment 4:** Divergent frontal–callosal architecture and a converging amygdala signature of early and late autism risk

In a final main experiment, we analyzed the associations of early- and late-diagnosis autism polygenic scores with the brain (Figs. 5 and S4.1 – S4.3).

Zhang et al. (2025) reported that genetic profiles in autism differ by age at diagnosis, with partially distinct polygenic architectures and only modest genetic correlation between early- and late-diagnosed groups. An early-diagnosis factor was associated with lower early social and communication abilities, whereas a late-diagnosis factor related to higher socioemotional problems and showed positive genetic correlations with ADHD and other psychiatric disorders. Using the age-of-diagnosis–stratified GWAS summary statistics, we constructed separate scores in UKB and entered them jointly into a single GLM. In our sample, the scores were effectively uncorrelated and derived from non-overlapping SNP sets, allowing straightforward estimation of dissociable effects as well as their combined associations.

A key finding from this analysis was increased neurite density (ICVF) in frontal white matter specific to the late-autism polygenic score (PS) and divergent from early-autism PS effect. Early-autism PS, in contrast, revealed reductions of neurite density in corpus callosum. Alterations in local frontal lobe connectivity have been implicated in autism across numerous studies (Courchesne & Pierce, 2005; Kana et al., 2014). A mixed age (range 3-36 y) study of corpus callosum found increased variability in midsagittal callosal area in autism vs. neurotypical controls, while higher callosal area was correlated with higher intelligence and faster processing speed within autism (Prigge et al., 2013). Interestingly, complete agenesis of corpus callosum has also been associated with increased autistic symptoms and communication problems (Paul et al., 2014).

Early-autism PS was further associated with decreased FA in anterior callosum and cingulum, while late autism PS was not. Late-autism PS was associated with higher FA values than early-PS in amygdalar anterior commissure, adjacent to uncinate fasciculus. A longitudinal study of white matter development in 125 autistic children (age range 2.5 – 7.0) revealed slower development of fractional anisotropy in cingulum and corpus callosum compared with controls (Andrews et al., 2021). Interestingly, greater increases in uncinate fasciculus FA were associated with decreased symptom severity over time.

Orientation dispersion of right uncinate fasciculus was further increased with higher late-autism PS against baseline and contrasting against early-autism PS. Right uncinate fasciculus microstructure has been related to reduced facial emotion discrimination (Coad et al., 2020).

Prior work on amygdala volume in ASD has generally reported overgrowth in early childhood (Mosconi et al., 2009; Sparks et al., 2002), and normal or reduced volumes in adolescent and adult individuals with ASD (Nacewicz et al., 2006; Schumann et al., 2004). Surprisingly, only the additive effect of early- plus late-autism scores was related to increased left amygdala volume. A recent study comparing 72 autistic participants and controls using the Hariri paradigm (Hariri et al., 2002) was not able to detect any difference in mean amygdala activity during emotional face processing, specifically the faces–shapes contrast (Langenbach et al., 2024). We likewise found no significant effects in the primary 10K-supervoxel analysis. In contrast, the 1K-supervoxel approach identified a distributed negative cortico-subcortical pattern associated with higher early-ASD polygenic load, consistent with an overall reduction in face-selective responses. Effects for the combined early + late PS were even more widespread and included the amygdala, whereas late PS alone showed no significant association.

**Experiment S1:** Brain-wide structural, iron and connectivity effects associated with smoking, coffee and alcohol

In a supplementary experiment, we investigated the most commonly used neuromodulators in the population, i.e., smoking, coffee and alcohol (Figs. S5.1 – S5.12). We found widespread and varied results across sub-modalities. Broadly speaking alcohol and nicotine were associated with more general, degenerative changes, while effects of coffee were less pronounced. Here, we will discuss a few findings of interest.

VBM analysis revealed widespread volume reductions across all substances, with direct comparisons in general suggesting that the associations with alcohol were the strongest overall, closely followed by nicotine, which specifically seemed to have the strongest association with the striatum.

Smoking showed strong evidence for greater volume in bilateral parahippocampal cortices and moderate to strong evidence for increased volume in dlPFC. Interestingly, reduction of dlPFC coupling with parahippocampal gyrus using electrical stimulation reduces craving in smokers (Yang et al., 2017).

Relation of coffee intake with volume increase in bilateral nucleus accumbens, bed nucleus of stria terminalis (BNST), hypothalamus and (centromedial) amygdala was strongly supported. Coffee consumption was further associated with volume reduction in vmPFC, which was also apparent when contrasting the effect of smoking.

Strong evidence was found for volume increase in bilateral precuneus with higher alcohol consumption.

We found robust evidence for iron accumulation (QSM) in striatum, substantia nigra and to a lesser extent hippocampus for both smoking and alcohol consumption, while alcohol was further associated with substantially increased QSM in cortex, cerebellum and amygdala. While to lesser extent, coffee consumption also showed strong evidence for accumulation in the striatum. Peak evidence for QSM increase and T2* decrease with coffee consumption was interestingly found in the sleep-regulating habenula (Hikosaka, 2010).

Diffusion sub-modalities further reinforced evidence for degenerative changes with respect to smoking and alcohol consumption but not coffee, showing global MD increases, and FA/ICVF decreases for the former two. Coffee consumption was, however, associated with loss of integrity in corpus callosum, albeit to a lesser extent than alcohol. Moreover, analysis of the warpfield Jacobian exhibited signs of smaller callosal volume in its anterior and posterior aspects. Remarkably, random allocation to caffeine or placebo for apnea in premature infants resulted in smaller corpus callosum at age 11 (Kelly et al., 2018).

Neurite density increase was found for coffee consumption in medial forebrain bundle, hypothalamus and white matter bordering on lateral amygdala surfaces, while both smoking and alcohol exhibited strong evidence for higher density surrounding the putamen.

Coffee use has been reported to reduce PD risk linearly and to be protective against Alzheimer's disease (Kolahdouzan & Hamadeh, 2017; Zhou & Zhang, 2021). In our analysis, coffee intake was, nevertheless, linked to increased QSM signal in striatum and medial substantia nigra. In PD, substantia nigra degeneration starts in its lateral compartment (Duke et al., 2007), which has been related to motor function, while medial substantia nigra co-activates with limbic regions (Zhang et al., 2017). For tobacco, amount smoked has been inversely related to PD risk (Mappin-Kasirer et al., 2020). Here, we observed stronger evidence for QSM increase in white matter between substantia nigra and red nucleus rather than substantia nigra.

Resting-state networks exhibited strong evidence for associations between alcohol and default-mode, a salience-like, and two frontoparietal networks. In the default mode network (RSN d25-2), alcohol intake was associated not only with increased coupling across core regions but also across numerous distributed cortical patches, consistent with reduced network specificity. This is consistent with reports of functional dedifferentiation after just one month of chronic alcohol intake (Pérez-Ramírez et al., 2022), commonly seen with aging (Malagurski et al., 2020). Alcohol similarly showed extensive coupling alterations including broader cortical recruitment in salience-like RSN d25-9 and two tested frontoparietal networks (RSN d25-6/11). Smoking and coffee, in contrast, related to focal and limited effects.

### Analysis of shared variance

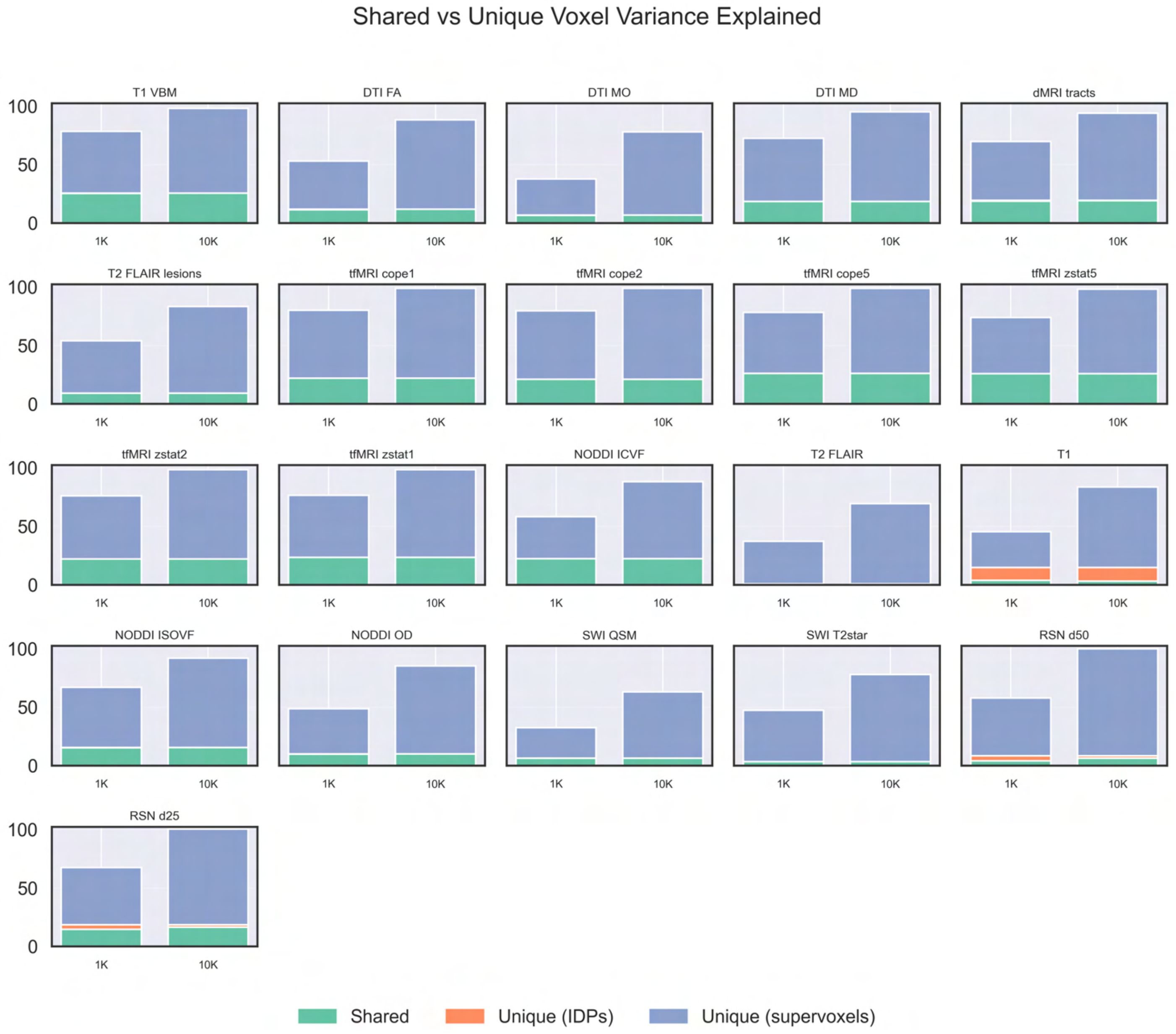


***Figure S6.1. Shared vs. unique voxel-space variance explained by IDPs and supervoxels (1K, 10K).*** *For each sub-modality, stacked bars (1K and 10K) show total explained voxel-space variance, split into shared variance and variance uniquely captured by IDPs or supervoxels. Across sub-modalities, IDPs contribute minimal variance beyond what is already captured by supervoxels, whereas supervoxels, especially at 10K, explain substantial additional unique voxel-space variance.*

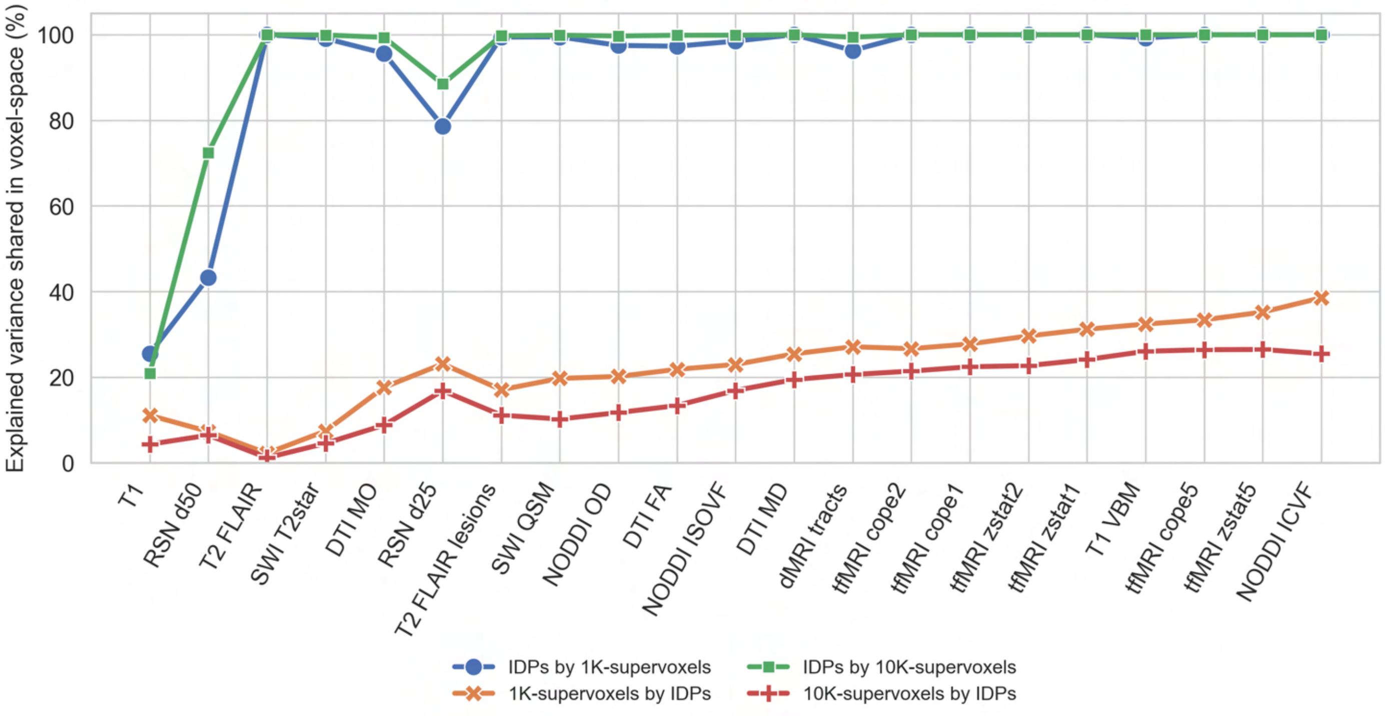


***Figure S6.2. Shared voxel-space variance between IDPs and supervoxels across sub-modalities.*** *Percent of each representation's explained voxel-space variance that is captured by the other.*

### Supervoxel visualization

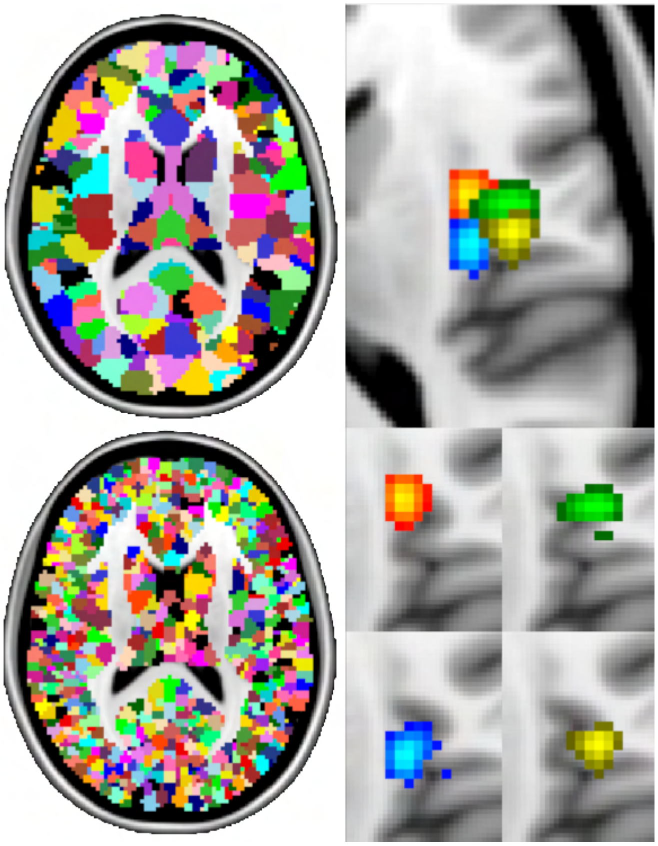

***Figure S7. Supervoxel parcellations and example units****. Left: binarized 1K- and more fine-grained 10K-supervoxels (T1 VBM). Right: four example 10K-supervoxels (non-binarized, thresholded at 25% of the peak value) in anterior insula/opercular region (for clarity, shown both separately from each other and combined into a single view).*
