## Supplementary material for "PANDORA: Population Archive of Neuroimaging Data Organized for Rapid Analysis": Peak Statistics

**Supplementary Peak Statistics Tables for Experiments 1-4 and S1**

**Peak statistics for experiment 1**: Lifetime Trauma

| *Sub-modality* | +/- | Area | Peak (MNI) | *Z* | $-\log_{10}(p, q)$ | *β* | p_hFDR |
|---|---|---|---|---|---|---|---|
| *VBM* | + | Visual cortex lh (V1) | 8, -102, 8 | 5.623 | 8.102, 3.337 | 0.015 | <0.001 |
| | − | Central amygdala rh | 18, -10, -12 | -7.110 | 12.237, 6.903 | 0.014 | <0.001 |
| *SWI QSM* | + | Substantia nigra rh | 10, -24, -15 | 7.502 | 13.504, 7.230 | 0.055 | <0.001 |
| | − | Lateral geniculate nucleus rh | 25, -22, -5 | -7.343 | 12.981, 6.697 | 0.027 | <0.001 |
| *SWI T2** | + | Fusiform gyrus lh (WM/GM boundary) | -48, -26, -22 | 6.939 | 11.704, 5.496 | 0.012 | <0.001 |
| | − | Anterior insula rh | 36, 7, -13 | -6.544 | 10.523, 4.649 | 0.029 | <0.001 |
| *NODDI ICVF* | + | Putamen lh | -23, 13, -6 | 5.609 | 7.991, 2.261 | 0.011 | <0.01 |
| | − | Centrum semiovale rh | 20, -19, 39 | -9.390 | 20.525, 15.383 | 0.024 | <0.001 |
| *NODDI OD* | + | Hypothalamus rh | 7, -7, -7 | 9.200 | 19.748, 13.969 | 0.010 | <0.001 |
| | − | Internal capsule lh (posterior limb) | -21, -16, 1 | -7.011 | 11.927, 6.453 | 0.004 | <0.001 |
| *DTI MD* | + | Anterior corona radiata rh | 25, 33, -1 | 7.983 | 15.147, 9.872 | 0.008 | <0.001 |
| | − | Medulla oblongata / spinal cord | 2, -45, -63 | 4.671 | 5.824, 0.500 | 0.061 | n.s. |
| *DTI FA* | + | Anterior putamen rh | 23, 18, 0 | 4.189 | 4.854, -0.342 | 0.006 | n.s. |
| | − | Cerebral peduncle lh | -15, -15, -15 | -10.054 | 23.357, 17.584 | 0.016 | <0.001 |
| *tfMRI zstat (faces)* | + | Precuneus rh | 8, -66, 35 | 5.263 | 7.150, 2.591 | 0.026 | <0.01 |
| | − | Supramarginal gyrus rh | 64, -32, 43 | -4.981 | 6.501, 2.054 | 0.024 | <0.05 |
| *tfMRI zstat (faces − shapes)* | + | Middle temporal gyrus lh | -63, -17, -19 | 6.040 | 9.112, 4.438 | 0.031 | <0.001 |
| | − | Angular gyrus rh | 40, 28, -6 | -5.475 | 7.660, 3.498 | 0.026 | <0.001 |
| *RSN d25-2 (DMN)* | + | Cerebellum lh (VIIb) | -38, -60, -54 | 3.892 | 4.303, 0.303 | 0.012 | n.s. |
| | − | Precuneus lh | -6, -59, 37 | -5.010 | 6.565, 2.074 | 0.033 | <0.05 |
| *RSN d25-9 (SN)* | + | Precuneus rh | 12, -66, 61 | 4.278 | 5.026, 0.514 | 0.021 | n.s. |
| | − | Middle temporal gyrus lh | -57, 2, -22 | -6.106 | 9.292, 4.787 | 0.022 | <0.001 |
| *RSN d25-13 (SMN)* | + | Postcentral gyrus lh | -30, -35, 72 | 4.408 | 5.283, 1.500 | 0.029 | n.s. |
| | − | Angular gyrus lh | -48, -51, 50 | - 5.017 | 6.580, 2.351 | 0.023 | <0.05 |

**Peak statistics for experiment 2:** Anxiety vs. Depressive Symptom Load

**VBM**

| *Contrast* | Area | Peak (MNI) | $Z$ | $-\log_{10}(p, q)$ | $\beta$ | p_hFDR |
|---|---|---|---|---|---|---|
| *Anxiety +* | Cerebellum VIIb rh | 18, -74, -50 | 5.175 | 6.943, 2.668 | 0.041 | <0.01 |
| *Depression +* | Putamen rh | 28, -10, 4 | 7.376 | 13.088, 8.156 | 0.021 | <0.001 |
| *Anxiety −* | dmPFC midline | 0, 40, 42 | -5.793 | 8.460, 3.427 | 0.038 | <0.001 |
| *Depression −* | Dorsal thalamus rh / lateral ventricle (posterior horn) | 12, -26, 18 | -7.181 | 12.460, 7.126 | 0.01 | <0.001 |
| *Anxiety > Depression* | lateral ventricle (posterior horn) rh / dorsal thalamus | 12, -26, 16 | 5.903 | 8.748, 4.217 | 0.024 | <0.001 |
| *Depression > Anxiety* | Cerebellar white matter rh | 30, -58, -40 | 5.294 | 7.223, 2.027 | 0.016 | <0.05 |
| *Combined +* | Putamen rh | -26, -8, 10 | 8.057 | 15.408, 10.477 | 0.011 | <0.001 |
| *Combined −* | Straight gyrus lh / inferior rostral gyrus | -2, 32, -18 | -9.107 | 19.374, 14.102 | 0.029 | <0.001 |

**SWI QSM**

| *Contrast* | Area | Peak (MNI) | $Z$ | $-\log_{10}(p, q)$ | $\beta$ | p_hFDR |
|---|---|---|---|---|---|---|
| *Anxiety +* | SMA lh | -27, -1, 71 | 4.815 | 6.133, 0.532 | 0.005 | n.s. |
| *Depression +* | Posterior insula rh | -38, -16, -2 | 6.611 | 10.720, 4.540 | 0.027 | <0.001 |
| *Anxiety −* | dmPFC lh | -16, 53, 39 | -5.254 | 7.128, 0.951 | 0.012 | n.s. |
| *Depression −* | BNST / anterior commissure lh | -5, 3, -3 | -6.537 | 10.505, 4.221 | 0.052 | <0.001 |
| *Anxiety > Depression* | Outside brain (temporal pole rh) | 20, 17, -31 | 4.996 | 6.534, 0.451 | 0.002 | n.s. |
| *Depression > Anxiety* | Posterior insula lh | -36, -16, 3 | 5.531 | 7.797, 2.078 | 0.036 | <0.05 |
| *Combined +* | Inferior fronto-occipital fasciculus rh | 36, -57, 1 | 6.877 | 11.516, 5.635 | 0.013 | <0.001 |
| *Combined −* | BNST lh | -5, 2, -3 | -7.325 | 12.923, 6.639 | 0.035 | <0.001 |

**Peak statistics for experiment 2:** Anxiety vs. Depressive Symptom Load

**SWI T2***

| *Contrast* | Area | Peak (MNI) | *Z* | $-\log_{10}(p, q)$ | *β* | p_hFDR |
|---|---|---|---|---|---|---|
| *Anxiety +* | dlPFC lh | -44, 51, 16 | 6.704 | 10.994, 5.505 | 0.055 | <0.001 |
| *Depression +* | Subcallosal area lh | -4, 21, -15 | 6.864 | 11.476, 5.118 | 0.011 | <0.001 |
| *Anxiety −* | Superior parietal lobule rh | 27, -76, 41 | -4.705 | 5.897, 0.352 | 0.014 | n.s. |
| *Depression −* | dlPFC lh | -50, 36, 29 | -8.423 | 16.738, 10.646 | 0.054 | <0.001 |
| *Anxiety > Depression* | dlPFC lh | -51, 36, 28 | 8.05 | 15.384, 9.411 | 0.08 | <0.001 |
| *Depression > Anxiety* | Subcallosal area rh (inferior rostral gyrus) | 4, 25, -17 | 5.828 | 8.552, 2.656 | 0.013 | <0.01 |
| *Combined +* | Periventricular white matter adjacent to right caudate nucleus | 19, 18, 18 | 6.615 | 10.730, 4.875 | 0.022 | <0.001 |
| *Combined −* | Anterior hippocampus lh | -29, -15, -24 | -6.711 | 11.013, 4.915 | 0.028 | <0.001 |

**NODDI ICVF**

| *Contrast* | Area | Peak (MNI) | *Z* | $-\log_{10}(p, q)$ | *β* | p_hFDR |
|---|---|---|---|---|---|---|
| *Anxiety +* | Postcentral gyrus | -60, -9, 32 | 4.263 | 4.996, -0.013 | 0.029 | n.s. |
| *Depression +* | Posterior medulla oblongata rh | 10, -43, -44 | 6.021 | 9.062, 3.569 | 0.029 | <0.001 |
| *Anxiety −* | Postcentral gyrus lh (WM/GM boundary) | -29, -37, 68 | -6.11 | 9.304, 3.525 | 0.052 | <0.001 |
| *Depression −* | Thalamus rh | 11, -13, 10 | -7.655 | 14.015, 8.660 | 0.016 | <0.001 |
| *Anxiety > Depression* | Thalamus rh | 12, -11, 8 | 5.23 | 7.072, 2.392 | 0.019 | <0.01 |
| *Depression > Anxiety* | Postcentral gyrus lh | -29, -37, 68 | 6.053 | 9.147, 3.368 | 0.095 | <0.01 |
| *Combined +* | Substantia innominata rh adjacent to anterior commissure | 13, 4, -8 | 6.567 | 10.589, 5.061 | 0.028 | <0.001 |
| *Combined −* | Thalamus lh | -9, -17, 7 | -8.757 | 17.999, 12.417 | 0.011 | <0.001 |

**Peak statistics for experiment 2:** Anxiety vs. Depressive Symptom Load

**NODDI OD**

| *Contrast* | Area | Peak (MNI) | *Z* | $-\log_{10}(p, q)$ | *β* | p_hFDR |
|---|---|---|---|---|---|---|
| *Anxiety* + | dmPFC lh | -11, 60, 21 | 4.188 | 4.851, 0.268 | 0.028 | n.s. |
| *Depression* + | Hypothalamus rh | 11, -7, -10 | 6.962 | 11.775, 6.393 | 0.008 | <0.001 |
| *Anxiety* − | Posterior corpus callosum | 3, -17, 24 | -5.587 | 7.938, 2.159 | 0.005 | <0.05 |
| *Depression* − | External capsule rh | 34, 0, 2 | -7.438 | 13.291, 7.552 | 0.016 | <0.001 |
| *Anxiety > Depression* | Putamen lh | -29, -2, 6 | 5.743 | 8.334, 2.555 | 0.039 | <0.01 |
| *Depression > Anxiety* | Paracentral lobule rh | 15, -27, 57 | 5.693 | 8.206, 2.889 | 0.038 | <0.01 |
| *Combined* + | Medial forebrain bundle rh | 10, -3, -9 | 7.28 | 12.776, 7.277 | 0.005 | <0.001 |
| *Combined* − | External capsule rh | 32, 3, 3 | -8.416 | 16.710, 10.952 | 0.012 | <0.001 |

**DTI MD**

| *Contrast* | Area | Peak (MNI) | *Z* | $-\log_{10}(p, q)$ | *β* | p_hFDR |
|---|---|---|---|---|---|---|
| *Anxiety* + | Parieto-occipital white matter lh (superior of calcarine sulcus) | -15, -52, 16 | 5.22 | 7.049, 1.899 | 0.009 | <0.05 |
| *Depression* + | Corticospinal tract lh above internal capsule | -24, -17, 20 | 6.72 | 11.042, 6.046 | 0.006 | <0.001 |
| *Anxiety* − | Inferior cerebral peduncle rh | 15, -37, -42 | -4.777 | 6.050, 0.740 | 0.035 | n.s. |
| *Depression* − | Fornix lh / third ventricle | -1, -19, 16 | -5.297 | 7.231, 1.706 | 0.051 | <0.05 |
| *Anxiety > Depression* | Fornix lh / third ventricle | -1, -18, 16 | 5.179 | 6.953, 1.446 | 0.089 | n.s. |
| *Depression > Anxiety* | Entorhinal cortex rh (posterior part) | 20, -25, -19 | 5.58 | 7.920, 2.887 | 0.126 | <0.01 |
| *Combined* + | Thalamus lh | -7, -15, 3 | 7.854 | 14.696, 9.314 | 0.006 | <0.001 |
| *Combined* − | Lateral ventricle rh (anterior horn) / WM boundary | 17, 31, 2 | -4.605 | 5.685, 0.371 | 0.015 | n.s. |

**Peak statistics for experiment 2:** Anxiety vs. Depressive Symptom Load

**DTI FA**

| *Contrast* | Area | Peak (MNI) | *Z* | $-\log_{10}$(p, q) | *β* | p_hFDR |
|---|---|---|---|---|---|---|
| *Anxiety +* | Fundus of callosal sulcus lh | -13, 9, 36 | 5.587 | 7.936, 2.163 | 0.021 | <0.05 |
| *Depression +* | Pons lh | -9, -30, -41 | 5.059 | 6.677, 1.108 | 0.034 | n.s. |
| *Anxiety −* | Lateral ventricle rh (anterior horn) / WM boundary | 15, 27, -6 | -4.73 | 5.951, 0.870 | 0.01 | n.s. |
| *Depression −* | Internal capsule lh | 16, 0, 14 | -6.969 | 11.797, 6.025 | 0.022 | <0.001 |
| *Anxiety > Depression* | Internal capsule lh | -14, 1, 11 | 5.704 | 8.232, 2.555 | 0.03 | <0.01 |
| *Depression > Anxiety* | Pons lh | -9, -30, -41 | 5.139 | 6.861, 1.105 | 0.063 | n.s. |
| *Combined +* | Amygdalo-striatal transition area lh posterior to anterior commissure | -28, -8, -12 | 6.277 | 9.763, 4.275 | 0.012 | <0.001 |
| *Combined −* | Forceps minor rh | 16, 33, 8 | -8.067 | 15.443, 9.868 | 0.017 | <0.001 |

**tfMRI zstat (shapes)**

| *Contrast* | Area | Peak (MNI) | *Z* | $-\log_{10}$(p, q) | *β* | p_hFDR |
|---|---|---|---|---|---|---|
| *Anxiety +* | Cerebellum lh (VIIIa) | -28, -40, -46 | 3.803 | 4.146, 0.000 | 0.021 | n.s. |
| *Depression +* | Basolateral amygdala rh | 18, 0, -24 | 4.839 | 6.186, 1.784 | 0.022 | <0.05 |
| *Anxiety −* | Temporo-parietal junction rh | 58, -49, 14 | -6.532 | 10.489, 5.905 | 0.048 | <0.001 |
| *Depression −* | Cerebellum lh (VIIIa) | -28, -40, -46 | -7.19 | 12.490, 7.529 | 0.04 | <0.001 |
| *Anxiety > Depression* | Cerebellum lh (VIIIa) | -28, -40, -46 | 6.008 | 9.029, 4.068 | 0.061 | <0.001 |
| *Depression > Anxiety* | Temporo-parietal junction rh | 58, -55, 22 | 5.248 | 7.114, 3.133 | 0.061 | <0.01 |
| *Combined +* | Basolateral amygdala rh | 18, -2, -24 | 3.484 | 3.607, 0.000 | 0.009 | n.s. |
| *Combined −* | Fronto-parietal operculum rh | 65, -13, 18 | -7.397 | 13.157, 8.668 | 0.03 | <0.001 |

**Peak statistics for experiment 2:** Anxiety vs. Depressive Symptom Load

#### tfMRI zstat (faces)

| *Contrast* | Area | Peak (MNI) | *Z* | $-\log_{10}(p, q)$ | *β* | p_hFDR |
|---|---|---|---|---|---|---|
| *Anxiety +* | Cerebellum lh (IX) | -14, -54, -46 | 4.168 | 4.813, 0.152 | 0.02 | n.s. |
| *Depression +* | Precuneus rh | 7, -66, 40 | 5.154 | 6.895, 2.493 | 0.034 | <0.01 |
| *Anxiety −* | Temporo-parietal junction rh | 58, -48, 13 | -6.17 | 9.467, 4.967 | 0.048 | <0.001 |
| *Depression −* | Cerebellum lh (VIIIa) | -28, -40, -46 | -6.87 | 11.494, 6.533 | 0.038 | <0.001 |
| *Anxiety > Depression* | Cerebellum lh (VIIIa) | -28, -40, -46 | 5.549 | 7.842, 2.882 | 0.055 | <0.01 |
| *Depression > Anxiety* | Superior temporal gyrus lh (posterior part) | 61, -43, 13 | 5.266 | 7.156, 3.116 | 0.076 | <0.01 |
| *Combined +* | Anterior putamen rh | 18, 12, -10 | 4.567 | 5.608, 0.647 | 0.013 | n.s. |
| *Combined −* | Parietal operculum rh (/ postcentral gyrus) | 65, -14, 18 | -7.728 | 14.263, 9.431 | 0.03 | <0.001 |

#### tfMRI zstat (faces−shapes)

| *Contrast* | Area | Peak (MNI) | *Z* | $-\log_{10}(p, q)$ | *β* | p_hFDR |
|---|---|---|---|---|---|---|
| *Anxiety +* | Supramarginal gyrus rh | 66, -32, 37 | 4.323 | 5.114, 1.244 | 0.027 | n.s. |
| *Depression +* | PACC rh | 5, -52, 25 | 4.402 | 5.271, 1.232 | 0.032 | n.s. |
| *Anxiety −* | Precuneus rh | 5, -55, 25 | -5.146 | 6.875, 2.256 | 0.039 | <0.05 |
| *Depression −* | vlPFC lh | -57, 14, 15 | -4.151 | 4.781, 0.758 | 0.024 | n.s. |
| *Anxiety > Depression* | Supramarginal gyrus rh | 64, -35, 44 | 4.252 | 4.975, 1.098 | 0.048 | n.s. |
| *Depression > Anxiety* | Precuneus rh | 5, -55, 26 | 5.058 | 6.675, 2.355 | 0.07 | <0.01 |
| *Combined +* | Lateral occipital cortex rh | 46, -81, 32 | 3.938 | 4.387, 0.563 | 0.013 | n.s. |
| *Combined −* | vlPFC rh | 53, 42, 10 | -4.729 | 5.947, 1.443 | 0.02 | n.s. |

**Peak statistics for experiment 2:** Anxiety vs. Depressive Symptom Load

**RSN d25-2 (DMN)**

| ***Contrast*** | **Area** | **Peak (MNI)** | ***Z*** | **$-\log_{10}$(p, q)** | ***β*** | **p_hFDR** |
|---:|---|---|---:|---|---:|---:|
| *Anxiety +* | Precuneus rh | 7, -65, 49 | 5.359 | 7.378, 2.832 | 0.05 | <0.01 |
| *Depression +* | dmPFC lh | -20, 20, 64 | 4.193 | 4.861, 0.572 | 0.024 | n.s. |
| *Anxiety −* | Angular gyrus rh | 56, -48, 48 | -4.985 | 6.510, 1.883 | 0.039 | <0.05 |
| *Depression −* | Cerebellum lh (VIIIb) | -26, -40, -52 | -4.501 | 5.472, 1.311 | 0.017 | n.s. |
| *Anxiety > Depression* | dmPFC rh | 22, 59, 31 | 4.676 | 5.834, 1.359 | 0.065 | n.s. |
| *Depression > Anxiety* | dmPFC rh | 7, 44, 39 | 4.498 | 5.464, 1.129 | 0.057 | n.s. |
| *Combined +* | Precuneus rh | 7, -65, 48 | 5.393 | 7.461, 2.835 | 0.029 | <0.01 |
| *Combined −* | Supramarginal gyrus lh | -63, -38, 38 | -4.342 | 5.151, 1.118 | 0.017 | n.s. |

**RSN d25-9 (SN)**

| ***Contrast*** | **Area** | **Peak (MNI)** | ***Z*** | **$-\log_{10}$(p, q)** | ***β*** | **p_hFDR** |
|---:|---|---|---:|---|---:|---:|
| *Anxiety +* | Cerebellum lh (VIIIa) | -32, -58, -60 | 4.776 | 6.049, 1.088 | 0.021 | n.s. |
| *Depression +* | Precuneus lh posterior of cingulate sulcus | -8, -53, 62 | 6.009 | 9.030, 4.224 | 0.039 | <0.001 |
| *Anxiety −* | vlPFC lh / lateral orbitofrontal cortex | -46, 42, -13 | -3.901 | 4.319, 0.226 | 0.022 | n.s. |
| *Depression −* | Paracentral lobule lh | -8, -22, 72 | -5.088 | 6.743, 2.466 | 0.042 | <0.01 |
| *Anxiety > Depression* | Cuneus lh posterior of parieto-occipital sulcus | -11, -83, 31 | 4.621 | 5.718, 1.488 | 0.052 | n.s. |
| *Depression > Anxiety* | Precuneus lh posterior of cingulate sulcus | -8, -53, 62 | 5.282 | 7.195, 2.235 | 0.063 | <0.05 |
| *Combined +* | Precuneus rh posterior of cingulate sulcus | 9, -50, 64 | 5.039 | 6.631, 1.938 | 0.019 | <0.05 |
| *Combined −* | Cerebellar vermis | 4, -64, -38 | -4.642 | 5.764, 0.943 | 0.011 | n.s. |

**Peak statistics for experiment 3:** EPHA3 SNP rs987748 (A:C)

| ***Sub-modality*** | **+/-** | **Area** | **Peak (MNI)** | ***Z*** | **-log$_{10}$(p/q)** | ***β*** | **p_hFDR** |
|---|---|---|---|---|---|---|---|
| *T1-VBM* | + | Anterior commissure | -2, 2, -4 | 14.409 | 46.647 / 41.614 | 0.019 | <0.001 |
| | - | Fusiform gyrus rh | 38, -20, -32 | -11.266 | 29.015 / 23.681 | 0.044 | <0.001 |
| *SWI-QSM* | + | Anterior commissure rh | 19, 1, -6 | 13.983 | 44.005 / 37.721 | 0.122 | <0.001 |
| | - | Parietal periventricular white matter lh | -20, -31, 28 | -9.616 | 21.465 / 15.181 | 0.027 | <0.001 |
| *SWI-T2** | + | Straight sinus | 1, -78, -5 | 8.760 | 18.012 / 11.928 | 0.047 | <0.001 |
| | - | Confluence of sinuses | 4, -84, -29 | -8.376 | 16.563 / 10.205 | 0.041 | <0.001 |
| *NODDI-ICVF* | + | Lateral occipital cortex lh (WM boundary) | -30, -81, 9 | 11.122 | 28.307 / 22.862 | 0.044 | <0.001 |
| | - | Anterior commissure / uncinate fasciculus rh | 26, -2, -10 | -19.821 | 87.006 / 81.227 | 0.062 | <0.001 |
| *NODDI-ISOVF* | + | Lateral occipital cortex rh | 28, -88, 21 | 9.879 | 22.590 / 16.811 | 0.086 | <0.001 |
| | - | Anterior commissure / uncinate fasciculus rh | 32, -3, -11 | -11.875 | 32.099 / 26.320 | 0.031 | <0.001 |
| *NODDI-OD* | + | Anterior commissure / uncinate fasciculus rh | 28, -4, -10 | 23.824 | 125.022 / 119.243 | 0.046 | <0.001 |
| | - | Uncinate fasciculus rh | 32, -11, -10 | -16.227 | 58.788 / 53.010 | 0.023 | <0.001 |
| *DTI-FA* | + | Arcuate fasciculus lh | -41, -43, 21 | 12.428 | 35.036 / 29.263 | 0.036 | <0.001 |
| | - | Anterior commissure / uncinate fasciculus rh | 27, -3, -10 | -22.880 | 115.431 / 109.658 | 0.091 | <0.001 |
| *DTI-MD* | + | Lateral occipital cortex rh (WM boundary) | 28, -88, 21 | 9.856 | 22.493 / 16.780 | 0.079 | <0.001 |
| | - | Parietal white matter rh | 18, -37, 36 | -10.595 | 25.805 / 20.026 | 0.012 | <0.001 |
| *DTI-L1* | + | External capsule rh | 33, -13, -10 | 12.060 | 33.066 / 27.287 | 0.015 | <0.001 |
| | - | Anterior commissure / uncinate fasciculus lh | -24, -2, -10 | -17.967 | 71.752 / 65.973 | 0.034 | <0.001 |
| *DTI-L2* | + | Anterior commissure / uncinate fasciculus rh | 27, -3, -11 | 17.061 | 64.843 / 59.064 | 0.036 | <0.001 |
| | - | Arcuate fasciculus lh | -44, -38, 3 | 13.303 | 39.953 / 34.174 | 0.023 | <0.001 |
| *tfMRI zstat (shapes)* | + | Inferior temporal gyrus lh (posteriorl part) | -49, -57, -20 | 7.744 | 14.317 / 9.685 | 0.036 | <0.001 |
| | - | Superior temporal gyrus lh (posterior part) | -60, -51, 14 | -8.744 | 17.950 / 12.990 | 0.046 | <0.001 |
| *tfMRI zstat (faces)* | + | Supramarginal gyrus rh | 66, -34, 33 | 7.004 | 11.905 / 7.281 | 0.034 | <0.001 |
| | - | Supramarginal gyrus rh | 65, -42, 19 | -7.586 | 13.781 / 9.021 | 0.039 | <0.001 |
| *tfMRI zstat (faces-shapes)* | + | Lateral occipital cortex lh | -42, -90, 9 | 8.186 | 15.869 / 11.118 | 0.047 | <0.001 |
| | - | Lateral occipital cortex rh | 34, -84, 29 | -6.308 | 9.849 / 5.536 | 0.032 | <0.001 |
| *RSN d25-2 (DMN)* | + | Cingulate gyrus lh (posterior) | -3, -29, 39 | 9.666 | 21.675 / 16.978 | 0.041 | <0.001 |
| | - | Inferior temporal gyrus lh (temporooccipital) | -57, -52, -17 | -7.714 | 14.216 / 9.374 | 0.037 | <0.001 |
| *RSN d25-6 (FPN)* | + | Precuneus lh | -7, -49, 41 | 9.574 | 21.290 / 16.531 | 0.050 | <0.001 |
| | - | Middle temporal gyrus lh | -63, -43, -4 | -11.169 | 28.541 / 23.761 | 0.072 | <0.001 |
| *RSN d25-18 (auditory)* | + | Supramarginal gyrus lh | -63, -41, 33 | 11.394 | 29.649 / 24.828 | 0.080 | <0.001 |
| | - | Superior temporal gyrus lh | -65, -39, 9 | -10.876 | 27.123 / 22.163 | 0.056 | <0.001 |

**Peak statistics for experiment 4:** Age-of-Diagnosis–Stratified Polygenic ASD Scores

**VBM**

| *Contrast* | Area | Peak (MNI) | *Z* | −$\log_{10}$(p, q) | *β* | p_hFDR |
|---|---|---|---|---|---|---|
| *Early ASD +* | Medulla oblongata rh | 4, -46, -50 | 5.048 | 6.652, 1.699 | 0.01 | n.s. |
| *Late ASD +* | Cerebellum lh (crus I) | -46, -48, -38 | 4.343 | 5.152, 0.738 | 0.023 | n.s. |
| *Early ASD −* | Cerebellum lh (VI) | -10, -80, -20 | -5.858 | 8.631, 3.469 | 0.029 | <0.01 |
| *Late ASD −* | Lateral occipital cortex lh | -52, -68, 12 | -4.896 | 6.311, 1.150 | 0.019 | n.s. |
| *Combined +* | Cerebellum rh (crus I) | 34, -54, -34 | 5.115 | 6.804, 2.258 | 0.015 | <0.05 |
| *Combined −* | Thalamus rh | 8, -16, -2 | -4.685 | 5.854, 1.119 | 0.002 | n.s. |
| *Early ASD > Late ASD* | Angular gyrus lh | -40, -76, 40 | 5.081 | 6.726, 1.796 | 0.021 | n.s. |
| *Late ASD > Early ASD* | Putamen rh | 30, 2, -6 | 5.28 | 7.190, 2.469 | 0.017 | <0.05 |

**SWI QSM**

| *Contrast* | Area | Peak (MNI) | *Z* | −$\log_{10}$(p, q) | *β* | p_hFDR |
|---|---|---|---|---|---|---|
| *Early ASD +* | Putamen lh (surface) | -32, -18, -1 | 5.257 | 7.136, 1.031 | 0.019 | n.s. |
| *Late ASD +* | Cerebellar midline | 2, -81, -41 | 4.71 | 5.907, -0.153 | 0.005 | n.s. |
| *Early ASD −* | Posterior insula rh | 39, -17, 11 | -5.404 | 7.488, 1.204 | 0.019 | n.s. |
| *Late ASD −* | Anterior commissure rh | 8, 3, -4 | -5.498 | 7.717, 1.739 | 0.048 | n.s. |
| *Combined +* | Putamen lh (surface) | -32, -19, -1 | 5.357 | 7.374, 1.204 | 0.016 | n.s. |
| *Combined −* | Anterior subiculum lh | -18, -16, -22 | -5.636 | 8.060, 1.776 | 0.027 | n.s. |
| *Early ASD > Late ASD* | Cerebellar WM rh | 8, -57, -32 | 5.106 | 6.783, 0.805 | 0.027 | n.s. |
| *Late ASD > Early ASD* | Cerebellum lh (VIIb) | -20, -68, -45 | 4.89 | 6.298, 0.529 | 0.018 | n.s. |

**Peak statistics for experiment 4:** Age-of-Diagnosis–Stratified Polygenic ASD Scores

**SWI T2***

| *Contrast* | Area | Peak (MNI) | *Z* | −log$_{10}$(p, q) | *β* | p_hFDR |
|---|---|---|---|---|---|---|
| *Early ASD +* | Outside brain (left cerebellum) | -39, -76, -58 | 5.22 | 7.049, 1.459 | 0.004 | n.s. |
| *Late ASD +* | Hypothalamus rh | 10, -3, -8 | 4.791 | 6.082, 0.492 | 0.014 | n.s. |
| *Early ASD −* | Cerebellum WM rh | 9, -56, -27 | -5.44 | 7.574, 1.928 | 0.021 | n.s. |
| *Late ASD −* | Transverse sinus lh | -54, -73, -23 | -4.444 | 5.355, 0.050 | 0.001 | n.s. |
| *Combined +* | Collateral sulcus between occipital gyri and fusiform cortex lh | -21, 10, -26 | 5.554 | 7.855, 2.032 | 0.022 | n.s. |
| *Combined −* | Fusiform cortex lh | -24, -68, -11 | -4.624 | 5.725, 0.401 | 0.015 | n.s. |
| *Early ASD > Late ASD* | Lateral ventricle lh (anterior horn) | -15, 29, 1 | 4.7 | 5.886, 0.224 | 0.028 | n.s. |
| *Late ASD > Early ASD* | Medial border of substantia nigra rh | 6, -10, -9 | 5.402 | 7.482, 1.366 | 0.024 | n.s. |

**NODDI ICVF**

| *Contrast* | Area | Peak (MNI) | *Z* | −log$_{10}$(p, q) | *β* | p_hFDR |
|---|---|---|---|---|---|---|
| *Early ASD +* | Pulvinar rh | 22, -28, -1 | 4.867 | 6.248, 1.066 | 0.011 | n.s. |
| *Late ASD +* | Claustrum rh | 25, 14, -12 | 5.661 | 8.124, 2.863 | 0.008 | <0.05 |
| *Early ASD −* | Posterior callosal fibers rh near cingulum bundle | 18, -20, 33 | -5.761 | 8.379, 3.339 | 0.019 | <0.01 |
| *Late ASD −* | External globus pallidus lh | -19, -2, 1 | -5.314 | 7.270, 2.207 | 0.03 | <0.05 |
| *Combined +* | Basal operculum rh | 40, 32, -4 | 5.647 | 8.088, 2.591 | 0.015 | <0.05 |
| *Combined −* | Corpus callosum midline | 0, -13, 23 | -4.235 | 4.943, -0.063 | 0.018 | n.s. |
| *Early ASD > Late ASD* | Cerebellar WM lh | -13, -57, -35 | 4.984 | 6.508, 1.043 | 0.037 | n.s. |
| *Late ASD > Early ASD* | Inferior fronto-occipital fasciculus lh | -40, -33, -7 | 6.173 | 9.474, 3.917 | 0.026 | <0.01 |

**Peak statistics for experiment 4:** Age-of-Diagnosis–Stratified Polygenic ASD Scores

### NODDI OD

| *Contrast* | Area | Peak (MNI) | *Z* | $-\log_{10}$(p, q) | *β* | p_hFDR |
|---|---|---|---|---|---|---|
| *Early ASD +* | Posterior cingulum bundle lh | -11, -37, 31 | 5.159 | 6.906, 1.648 | 0.006 | n.s. |
| *Late ASD +* | Uncinate fascicle rh (border) | 37, -10, -16 | 6.277 | 9.763, 3.984 | 0.007 | <0.01 |
| *Early ASD −* | Cerebellar WM rh | 27, -60, -40 | -5.657 | 8.113, 2.334 | 0.015 | <0.05 |
| *Late ASD −* | Substantia nigra lh | -10, -13, -9 | -5.074 | 6.711, 1.270 | 0.009 | n.s. |
| *Combined +* | Pons | 1, -37, -30 | 5.197 | 6.994, 1.869 | 0.007 | n.s. |
| *Combined −* | Cerebellum rh GM/WM boundary | 34, -55, -37 | -5.565 | 7.882, 2.401 | 0.013 | <0.05 |
| *Early ASD > Late ASD* | Internal globus pallidus rh | 17, -3, -5 | 5.675 | 8.158, 2.380 | 0.017 | <0.05 |
| *Late ASD > Early ASD* | Uncinate fascicle rh | 35, -4, -13 | 5.887 | 8.707, 3.609 | 0.013 | <0.01 |

### DTI FA

| *Contrast* | Area | Peak (MNI) | *Z* | $-\log_{10}$(p, q) | *β* | p_hFDR |
|---|---|---|---|---|---|---|
| *Early ASD +* | Cerebellar WM rh | 26, -61, -40 | 5.257 | 7.135, 1.593 | 0.02 | n.s. |
| *Late ASD +* | Cerebellar WM rh | 34, -51, -38 | 4.702 | 5.891, 0.719 | 0.021 | n.s. |
| *Early ASD −* | Centrum semiovale lh | -23, -25, 32 | -6.143 | 9.393, 4.280 | 0.018 | <0.01 |
| *Late ASD −* | Cerebellar WM rh | 17, -59, -32 | -4.923 | 6.371, 0.634 | 0.012 | n.s. |
| *Combined +* | Cerebellar WM rh | -34, -52, -38 | 5.499 | 7.719, 1.946 | 0.026 | <0.05 |
| *Combined −* | Insular WM rh | 34, -4, 11 | -5.047 | 6.648, 1.324 | 0.009 | n.s. |
| *Early ASD > Late ASD* | Middle cerebellar peduncle lh | -16, -38, -34 | 5.162 | 6.913, 1.530 | 0.019 | n.s. |
| *Late ASD > Early ASD* | Lateral amygdala WM rh / temporal horn of lateral ventricle | 31, -7, -18 | 5.579 | 7.917, 2.859 | 0.011 | <0.05 |

**Peak statistics for experiment 4:** Age-of-Diagnosis–Stratified Polygenic ASD Scores

**DTI MD**

| *Contrast* | Area | Peak (MNI) | *Z* | −$\log_{10}$(p, q) | *β* | p_hFDR |
|---|---|---|---|---|---|---|
| *Early ASD +* | dmPFC rh | 18, 10, 47 | 5.325 | 7.296, 2.019 | 0.004 | <0.05 |
| *Late ASD +* | dmPFC rh | 20, 14, 62 | 3.673 | 3.921, -0.061 | 0.042 | n.s. |
| *Early ASD −* | Lateral ventricle / mid corpus callosum rh | 6, 1, 33 | -5.191 | 6.980, 1.413 | 0.004 | n.s. |
| *Late ASD −* | Posterior callosal fibres lh | -19, -39, 29 | -4.94 | 6.408, 1.707 | 0.005 | n.s. |
| *Combined +* | dlPFC lh | -31, 15, 56 | 4.214 | 4.902, -0.060 | 0.051 | n.s. |
| *Combined −* | Cingulum bundle rh | 7, 8, 31 | -4.872 | 6.259, 1.023 | 0.004 | n.s. |
| *Early ASD > Late ASD* | Frontal lobe WM lh | -30, 44, 4 | 5.722 | 8.278, 3.093 | 0.009 | <0.05 |
| *Late ASD > Early ASD* | vlPFC lh | -47, 14, 22 | 4.484 | 5.436, 0.282 | 0.033 | n.s. |

**Task fMRI: Shapes (zstat 1)**

| *Contrast* | Area | Peak (MNI) | *Z* | −$\log_{10}$(p, q) | *β* | p_hFDR |
|---|---|---|---|---|---|---|
| *Early ASD +* | VTA | 2, -20, -18 | 4.528 | 5.526, 0.853 | 0.014 | n.s. |
| *Late ASD +* | dlPFC rh | 39, 46, 35 | 4.192 | 4.860, 0.359 | 0.02 | n.s. |
| *Early ASD −* | Lateral amygdala lh | -22, -4, -30 | -3.793 | 4.128, 0.000 | 0.013 | n.s. |
| *Late ASD −* | Cerebellum lh (crus I) | -34, -74, -34 | -3.631 | 3.850, 0.000 | 0.013 | n.s. |
| *Combined +* | dlPFC rh | 42, 43, 36 | 5.084 | 6.733, 2.253 | 0.023 | n.s. |
| *Combined −* | PCC rh | 5, -45, 33 | -3.727 | 4.013, 0.000 | 0.014 | n.s. |
| *Early ASD > Late ASD* | Cerebellum lh (crus I) | -12, -76, -30 | 3.957 | 4.420, 0.153 | 0.02 | n.s. |
| *Late ASD > Early ASD* | Pulvinar lh | -24, -30, -2 | 3.53 | 3.682, 0.000 | 0.014 | n.s. |

**Peak statistics for experiment 4:** Age-of-Diagnosis–Stratified Polygenic ASD Scores

**Task fMRI: Faces (zstat 2)**

| *Contrast* | Area | Peak (MNI) | *Z* | −$\log_{10}$(p, q) | *β* | p_hFDR |
|---|---|---|---|---|---|---|
| *Early ASD +* | Cerebellum rh (VIIb/VIIIa) | 24, -66, -44 | 3.528 | 3.679, 0.000 | 0.014 | n.s. |
| *Late ASD +* | Anterior insula rh / basal operculum rh | 42, 21, -7 | 3.885 | 4.291, 0.000 | 0.015 | n.s. |
| *Early ASD −* | PCC rh | 4, -44, 32 | -4.797 | 6.093, 1.642 | 0.018 | n.s. |
| *Late ASD −* | Lateral occipital cortex lh | -46, -83, 22 | -4.122 | 4.726, 0.464 | 0.023 | n.s. |
| *Combined +* | Putamen rh | 28, 10, 8 | 3.917 | 4.348, 0.000 | 0.012 | n.s. |
| *Combined −* | PCC rh | 5, -45, 33 | -5.532 | 7.801, 3.220 | 0.021 | <0.01 |
| *Early ASD > Late ASD* | Cerebellum rh (VIIIa) | 24, -60, -46 | 3.739 | 4.035, 0.000 | 0.02 | n.s. |
| *Late ASD > Early ASD* | Fusiform gyrus lh | -24, -52, -18 | 4.05 | 4.592, 0.057 | 0.022 | n.s. |

**Task fMRI: Faces−Shapes (zstat 5)**

| *Contrast* | Area | Peak (MNI) | *Z* | −$\log_{10}$(p, q) | *β* | p_hFDR |
|---|---|---|---|---|---|---|
| *Early ASD +* | Entorhinal cortex rh | 14, -10, -24 | 3.369 | 3.423, 0.000 | 0.014 | n.s. |
| *Late ASD +* | Orbitofrontal cortex rh | 16, 23, -24 | 3.175 | 3.126, 0.000 | 0.013 | n.s. |
| *Early ASD −* | Anterior thalamus lh | -8, -6, 8 | -4.766 | 6.027, 1.826 | 0.017 | n.s. |
| *Late ASD −* | Mesencephalon rh | 8, -32, -16 | -4.786 | 6.071, 1.361 | 0.017 | n.s. |
| *Combined +* | Pons rh | 6, -14, -22 | 3.113 | 3.033, 0.000 | 0.012 | n.s. |
| *Combined −* | Cerebellum lh (crus II) | -44, -52, -46 | -4.808 | 6.119, 2.313 | 0.02 | n.s. |
| *Early ASD > Late ASD* | Mid hippocampus rh | 32, -22, -18 | 3.193 | 3.153, 0.000 | 0.018 | n.s. |
| *Late ASD > Early ASD* | Precuneus rh | 6, -52, 14 | 3.805 | 4.150, 0.213 | 0.023 | n.s. |

**Peak statistics for experiment S1:** Smoking, Coffee and Alcohol

**VBM**

| *Contrast* | Area | Peak (MNI) | *Z* | $-\log_{10}(p, q)$ | *β* | p_hFDR |
|---|---|---|---|---|---|---|
| *Smoking* + | Thalamus lh (dorsolateral pulvinar) bordering on ventricle | -18, -32, 12 | 8.804 | 18.181, 12.860 | 0.018 | <0.001 |
| *Coffee* + | BNST / anterior commissure rh | 6, 2, -4 | 12.564 | 35.776, 30.442 | 0.015 | <0.001 |
| *Alcohol* + | Confluence of sinuses | -2, -84, -18 | 10.04 | 23.294, 17.960 | 0.048 | <0.001 |
| *Smoking* − | Uncinate fascicle rh (caudal to putamen) | 18, 18, -12 | -12.605 | 36.001, 30.963 | 0.011 | <0.001 |
| *Coffee* − | Cerebellum lh (V / white matter) | -8, -56, -24 | -9.292 | 20.119, 14.974 | 0.005 | <0.001 |
| *Alcohol* − | Hypothalamus rh | 10, -2, -8 | -17.49 | 68.072, 62.738 | 0.006 | <0.001 |
| *Combined* + | Ventral tegmental area (VTA) | 0, -18, -8 | 13.044 | 38.463, 33.129 | 0.01 | <0.001 |
| *Combined* − | Putamen rh | 28, -2, -4 | -17.57 | 68.679, 63.482 | 0.025 | <0.001 |
| *Smoking* > *Coffee* | Lateral ventricle lh | -22, -36, 12 | 8.143 | 15.715, 10.381 | 0.014 | <0.001 |
| *Smoking* > *Alcohol* | Lateral ventricle rh | 32, -44, 2 | 8.181 | 15.851, 10.517 | 0.023 | <0.001 |
| *Coffee* > *Alcohol* | Lateral hypothalamus lh / medial forebrain lh | -8, 0, -6 | 14.504 | 47.242, 42.070 | 0.013 | <0.001 |
| *Coffee* > *Smoking* | Anterior commissure rh | 8, 2, -6 | 13.602 | 41.709, 36.375 | 0.015 | <0.001 |
| *Alcohol* > *Smoking* | Transverse sinus rh | 46, -72, -20 | 7.208 | 12.546, 7.212 | 0.044 | <0.001 |
| *Alcohol* > *Coffee* | Dorsal thalamus (/ lateral ventricle) lh | -12, -26, 16 | 6.486 | 10.355, 5.021 | 0.016 | <0.001 |

**Peak statistics for experiment S1:** Smoking, Coffee and Alcohol

**SWI QSM**

| ***Contrast*** | **Area** | **Peak (MNI)** | ***Z*** | **$-\log_{10}(p, q)$** | ***β*** | **p_hFDR** |
|---|---|---|---|---|---|---|
| *Smoking +* | Putamen lh | -28, -16, 2 | 19.33 | 82.826, 76.542 | 0.175 | <0.001 |
| *Coffee +* | Habenula rh | 5, -25, 2 | 13.481 | 40.998, 35.015 | 0.091 | <0.001 |
| *Alcohol +* | Putamen rh | 29, -16, 4 | 19.018 | 80.217, 73.933 | 0.185 | <0.001 |
| *Smoking −* | Fronto-parietal operculum rh | 32, 6, 14 | -14.519 | 47.335, 41.051 | 0.03 | <0.001 |
| *Coffee −* | Lateral ventricle / caudate rh | 7, 8, 1 | -8.074 | 15.468, 9.184 | 0.037 | <0.001 |
| *Alcohol −* | External capsule / putaminal surface lh | -33, -7, 0 | -13.939 | 43.737, 37.453 | 0.053 | <0.001 |
| *Combined +* | Putamen lh | -28, -16, 4 | 27.413 | 165.020, 158.736 | 0.224 | <0.001 |
| *Combined −* | Internal capsule lh | -19, -6, 14 | -20.395 | 92.033, 85.749 | 0.053 | <0.001 |
| *Smoking > Coffee* | Putamen lh | -29, -17, 1 | 10.585 | 25.759, 19.475 | 0.142 | <0.001 |
| *Smoking > Alcohol* | External globus pallidus lh | -26, -11, -3 | 7.907 | 14.879, 8.595 | 0.07 | <0.001 |
| *Coffee > Alcohol* | External globus pallidus rh | 19, 4, 0 | 9.256 | 19.972, 13.688 | 0.1 | <0.001 |
| *Coffee > Smoking* | Posterior claustrum lh | -32, -6, 13 | 9.569 | 21.269, 15.242 | 0.033 | <0.001 |
| *Alcohol > Smoking* | Lateral ventricle lh (posterior horn) | -32, -42, -1 | 10.733 | 26.450, 20.166 | 0.064 | <0.001 |
| *Alcohol > Coffee* | Putamen lh | -29, -19, 4 | 10.533 | 25.515, 19.257 | 0.148 | <0.001 |

**Peak statistics for experiment S1:** Smoking, Coffee and Alcohol

**SWI T2***

| *Contrast* | Area | Peak (MNI) | *Z* | $-\log_{10}(p, q)$ | *β* | p_hFDR |
|---|---|---|---|---|---|---|
| *Smoking +* | Frontal midline / CSF | -1, 67, 32 | 11.153 | 28.460, 22.101 | 0.009 | <0.001 |
| *Coffee +* | CSF superior to dmPFC rh | 21, 22, 73 | 5.567 | 7.886, 2.109 | 0.003 | <0.01 |
| *Alcohol +* | CSF adjacent to dlPFC rh | 57, 9, 52 | 14.454 | 46.930, 40.571 | 0.01 | <0.001 |
| *Smoking −* | Putamen rh | 28, -13, 5 | -14.931 | 49.984, 43.889 | 0.06 | <0.001 |
| *Coffee −* | Habenula lh | -3, -26, 3 | -8.8 | 18.163, 11.804 | 0.027 | <0.001 |
| *Alcohol −* | Putamen lh | -29, -17, 3 | -15.839 | 56.079, 49.720 | 0.068 | <0.001 |
| *Combined +* | CSF adjacent to dlPFC lh | -49, 11, 59 | 16.619 | 61.599, 55.240 | 0.01 | <0.001 |
| *Combined −* | Putamen rh | 29, -15, 4 | -21.32 | 100.429, 94.070 | 0.08 | <0.001 |
| *Smoking > Coffee* | Frontal midline / CSF | -1, 67, 32 | 8.704 | 17.794, 11.435 | 0.011 | <0.001 |
| *Smoking > Alcohol* | Anterior horn of lateral ventricle lh | -4, 19, 6 | 6.387 | 10.072, 3.713 | 0.035 | <0.001 |
| *Coffee > Alcohol* | Putamen lh | -29, -17, 3 | 9.772 | 22.130, 15.841 | 0.06 | <0.001 |
| *Coffee > Smoking* | Putamen rh | 27, -11, 6 | 9.11 | 19.385, 13.206 | 0.051 | <0.001 |
| *Alcohol > Smoking* | Basal operculum / inferior frontal gyrus lh | -31, 17, -24 | 8.251 | 16.105, 10.228 | 0.067 | <0.001 |
| *Alcohol > Coffee* | Occipital gyri (superior parts) | -15, -98, 29 | 9.407 | 20.593, 14.455 | 0.054 | <0.001 |

**Peak statistics for experiment S1:** Smoking, Coffee and Alcohol

**DTI MD**

| *Contrast* | Area | Peak (MNI) | *Z* | −log10(p, q) | *β* | p_hFDR |
|---|---|---|---|---|---|---|
| *Smoking +* | Thalamus lh | -9, -20, 1 | 9.904 | 22.700, 16.921 | 0.008 | <0.001 |
| *Coffee +* | Fourth ventricle / superior cerebellar peduncle rh | 6, -42, -29 | 7.164 | 12.406, 6.752 | 0.023 | <0.001 |
| *Alcohol +* | Anterior corpus callosum rh | 6, 22, 15 | 14.766 | 48.915, 43.136 | 0.03 | <0.001 |
| *Smoking −* | Pons | 5, -24, -35 | -6.168 | 9.462, 4.256 | 0.028 | <0.001 |
| *Coffee −* | Callosal fibers rh in proximity of cingulum bundle | -9, -6, 30 | -5.925 | 8.807, 3.198 | 0.004 | <0.001 |
| *Alcohol −* | Pons rh | 13, -29, -27 | -8.201 | 15.924, 10.187 | 0.011 | <0.001 |
| *Combined +* | Fourth ventricle / superior cerebellar peduncle lh | -5, -41, -29 | 16.446 | 60.345, 54.566 | 0.055 | <0.001 |
| *Combined −* | Pons lh | -12, -31, -28 | -8.931 | 18.677, 13.285 | 0.012 | <0.001 |
| *Smoking > Coffee* | Insula rh GM/WM boundary | 37, 3, -15 | 7.623 | 13.907, 8.660 | 0.013 | <0.001 |
| *Smoking > Alcohol* | Cerebellar WM lh | -14, -68, -38 | 6.653 | 10.843, 5.325 | 0.007 | <0.001 |
| *Coffee > Alcohol* | Superior cerebellar peduncle lh | -2, -47, -24 | 6.201 | 9.551, 4.174 | 0.025 | <0.001 |
| *Coffee > Smoking* | Vermis GM/WM boundary | -2, -43, -24 | 5.75 | 8.352, 2.862 | 0.077 | <0.01 |
| *Alcohol > Smoking* | Lateral ventricle lh (posterior horn) | -31, -46, 3 | 7.206 | 12.542, 6.763 | 0.078 | <0.001 |
| *Alcohol > Coffee* | Anterior corpus callosum lh | -7, 22, 16 | 8.351 | 16.472, 10.973 | 0.018 | <0.001 |

**Peak statistics for experiment S1:** Smoking, Coffee and Alcohol

**DTI FA**

| *Contrast* | Area | Peak (MNI) | *Z* | $-\log_{10}(p, q)$ | *β* | p_hFDR |
|---|---|---|---|---|---|---|
| *Smoking +* | Putamen lh | -30, -13, 5 | 9.849 | 22.463, 16.732 | 0.033 | <0.001 |
| *Coffee +* | Thalamus lh | -6, -17, 6 | 5.503 | 7.729, 2.547 | 0.01 | <0.01 |
| *Alcohol +* | Central tegmental tract rh | 5, -35, -25 | 9.778 | 22.156, 16.465 | 0.025 | <0.001 |
| *Smoking −* | Hypothalamus lh | -7, -6, -5 | -11.308 | 29.224, 23.451 | 0.02 | <0.001 |
| *Coffee −* | Mid-posterior callosum (apical edge) | -2, -14, 27 | -7.2 | 12.522, 7.335 | 0.017 | <0.001 |
| *Alcohol −* | WM/CSF boundary lateral ventricle lh (temporal horn) | -36, -31, -7 | -12.068 | 33.110, 27.338 | 0.02 | <0.001 |
| *Combined +* | WM near superior cerebellar peduncle lh | -6, -37, -29 | 12.716 | 36.620, 30.847 | 0.028 | <0.001 |
| *Combined −* | Fornix rh | 3, 1, -2 | -14.832 | 49.342, 43.569 | 0.027 | <0.001 |
| *Smoking > Coffee* | Uncinate fascicle rh | 34, -1, -19 | 6.978 | 11.825, 6.257 | 0.022 | <0.001 |
| *Smoking > Alcohol* | Uncinate fascicle lh | -35, -2, -14 | 6.885 | 11.538, 6.282 | 0.037 | <0.001 |
| *Coffee > Alcohol* | Superior parietal lobule lh / parieto-occipital transition area | -21, -83, 42 | 7.399 | 13.165, 7.392 | 0.019 | <0.001 |
| *Coffee > Smoking* | Anterior thalamus/WM boundary | -7, -7, -4 | 7.388 | 13.127, 7.354 | 0.017 | <0.001 |
| *Alcohol > Smoking* | Cerebellar WM lh | -6, -54, -25 | 8.324 | 16.370, 10.706 | 0.026 | <0.001 |
| *Alcohol > Coffee* | Anterior putamen rh | 16, 14, -7 | 6.923 | 11.656, 5.883 | 0.016 | <0.001 |

**Peak statistics for experiment S1:** Smoking, Coffee and Alcohol

**DTI MO**

| *Contrast* | Area | Peak (MNI) | *Z* | $-\log_{10}$(p, q) | *β* | p_hFDR |
|---|---|---|---|---|---|---|
| *Smoking +* | Internal capsule rh (genu) | 14, 4, 8 | 9.58 | 21.312, 15.606 | 0.01 | <0.001 |
| *Coffee +* | Posterior callosal fibers rh | 20, -30, 30 | 6.647 | 10.825, 5.576 | 0.018 | <0.001 |
| *Alcohol +* | Posterior callosal fibers rh | 23, -36, 28 | 11.825 | 31.837, 26.344 | 0.039 | <0.001 |
| *Smoking −* | Posterior putamen rh /WM boundary | 28, -14, -2 | -10.86 | 27.049, 21.270 | 0.023 | <0.001 |
| *Coffee −* | Pulvinar rh | 13, -32, 0 | -7.113 | 12.247, 6.468 | 0.02 | <0.001 |
| *Alcohol −* | Cingulum bundle rh | 10, 29, 20 | -12.859 | 37.419, 31.640 | 0.029 | <0.001 |
| *Combined +* | WM superior to posterior horn of lateral ventricle rh | 22, -37, 28 | 14.324 | 46.109, 40.330 | 0.04 | <0.001 |
| *Combined −* | Posterior putamen rh /WM boundary | 28, -14, -2 | -13.412 | 40.589, 34.811 | 0.025 | <0.001 |
| *Smoking > Coffee* | Posterior internal capsule lh / pallidum lh | -23, -18, 1 | 8 | 15.206, 9.427 | 0.009 | <0.001 |
| *Smoking > Alcohol* | Uncinate fascicle lh | -36, -7, -15 | 7.805 | 14.526, 9.153 | 0.031 | <0.001 |
| *Coffee > Alcohol* | Cingulum bundle rh | 9, 37, 9 | 8.343 | 16.441, 10.663 | 0.032 | <0.001 |
| *Coffee > Smoking* | Pulvinar lh | -8, -24, 12 | 7.6 | 13.830, 8.052 | 0.038 | <0.001 |
| *Alcohol > Smoking* | Pulvinar lh | -8, -25, 11 | 7.345 | 12.986, 7.283 | 0.039 | <0.001 |
| *Alcohol > Coffee* | Basal forebrain rh | 12, -2, -9 | 7.241 | 12.651, 6.954 | 0.013 | <0.001 |

**Peak statistics for experiment S1:** Smoking, Coffee and Alcohol

**NODDI ICVF**

| *Contrast* | Area | Peak (MNI) | *Z* | $-\log_{10}(p, q)$ | $\beta$ | p_hFDR |
|---|---|---|---|---|---|---|
| *Smoking +* | Putamen rh | 31, -11, -3 | 14.722 | 48.633, 42.854 | 0.074 | <0.001 |
| *Coffee +* | Medial forebrain bundle lh | -9, -3, -8 | 8.944 | 18.727, 12.948 | 0.033 | <0.001 |
| *Alcohol +* | Putamen rh | 32, -9, 1 | 12.686 | 36.449, 31.090 | 0.058 | <0.001 |
| *Smoking −* | Precentral gyrus WM lh | -11, -26, 65 | -9.949 | 22.896, 17.240 | 0.023 | <0.001 |
| *Coffee −* | Precentral cortex lh | -28, -24, 64 | -5.996 | 8.994, 3.522 | 0.036 | <0.001 |
| *Alcohol −* | Putamen rh | 43, -15, 62 | -11.541 | 30.388, 24.761 | 0.118 | <0.001 |
| *Combined +* | Putamen rh | 31, -11, -3 | 19.234 | 82.014, 76.486 | 0.087 | <0.001 |
| *Combined −* | dlPFC lh | -33, 8, 61 | -14.855 | 49.488, 43.994 | 0.158 | <0.001 |
| *Smoking > Coffee* | Putamen lh | -29, -13, -1 | 9.355 | 20.380, 14.601 | 0.078 | <0.001 |
| *Smoking > Alcohol* | Superior temporal gyrus lh | -55, 0, -5 | 6.495 | 10.382, 4.603 | 0.017 | <0.001 |
| *Coffee > Alcohol* | Cingulum bundle rh | 7, 28, 22 | 8.966 | 18.814, 13.194 | 0.017 | <0.001 |
| *Coffee > Smoking* | Lateral amygdala rh | 28, -5, -16 | 7.61 | 13.862, 8.443 | 0.01 | <0.001 |
| *Alcohol > Smoking* | Middle longitudinal fasciculus lh | -38, -35, 1 | 6.916 | 11.634, 6.527 | 0.028 | <0.001 |
| *Alcohol > Coffee* | Putamen lh | -30, -12, 0 | 7.802 | 14.516, 9.246 | 0.066 | <0.001 |

**Peak statistics for experiment S1:** Smoking, Coffee and Alcohol

**NODDI OD**

| ***Contrast*** | **Area** | **Peak (MNI)** | ***Z*** | **$-\log_{10}(p, q)$** | **$\beta$** | **p_hFDR** |
|---|---|---|---|---|---|---|
| *Smoking +* | Substantia nigra lh | -10, -13, -10 | 11.185 | 28.615, 22.836 | 0.023 | <0.001 |
| *Coffee +* | Caudate nucleus rh | 11, 9, -5 | 8.666 | 17.651, 11.872 | 0.02 | <0.001 |
| *Alcohol +* | Anterior commissure rh | 5, 2, -4 | 12.501 | 35.434, 29.655 | 0.023 | <0.001 |
| *Smoking −* | Uncinate fascicle lh | -34, -4, -15 | -11.739 | 31.397, 25.766 | 0.016 | <0.001 |
| *Coffee −* | Thalamus lh | -7, -18, 1 | -6.216 | 9.595, 3.816 | 0.01 | <0.001 |
| *Alcohol −* | WM superior to posterior horn of lateral ventricle rh | 22, -39, 27 | -12.017 | 32.842, 27.063 | 0.017 | <0.001 |
| *Combined +* | Caudate nucleus lh (surface) | -12, 11, 2 | 14.964 | 50.198, 44.419 | 0.026 | <0.001 |
| *Combined −* | WM superior to posterior horn of lateral ventricle rh | 22, -39, 27 | -14.581 | 47.732, 41.953 | 0.019 | <0.001 |
| *Smoking > Coffee* | Putamen rh | 32, -8, -3 | 6.476 | 10.328, 4.549 | 0.02 | <0.001 |
| *Smoking > Alcohol* | Substantia nigra lh | -8, -17, -12 | 7.38 | 13.101, 7.766 | 0.016 | <0.001 |
| *Coffee > Alcohol* | Anterior callosal fibers rh | 13, 32, 8 | 7.371 | 13.073, 7.294 | 0.004 | <0.001 |
| *Coffee > Smoking* | Uncinate fascicle lh | -34, -3, -15 | 8.853 | 18.370, 12.591 | 0.017 | <0.001 |
| *Alcohol > Smoking* | Fronto-basal WM / Forceps minor lh | -14, 32, -8 | 8.364 | 16.518, 10.939 | 0.025 | <0.001 |
| *Alcohol > Coffee* | Putamen lh | -31, -7, 0 | 6.935 | 11.692, 6.057 | 0.029 | <0.001 |

**Peak statistics for experiment S1:** Smoking, Coffee and Alcohol

**Warpfield Jacobian**

| *Contrast* | Area | Peak (MNI) | *Z* | $-\log_{10}(p, q)$ | $\beta$ | p_hFDR |
|---|---|---|---|---|---|---|
| *Smoking +* | Anterior capsula interna lh | -17, 5, 10 | 6.796 | 11.270, 6.342 | 0.014 | <0.001 |
| *Coffee +* | Cerebellum rh (VIIIa) | 23, -67, -58 | 5.929 | 8.816, 4.373 | 0.013 | <0.001 |
| *Alcohol +* | Occipital gyri lh | -7, -87, -16 | 9.68 | 21.735, 16.163 | 0.037 | <0.001 |
| *Smoking −* | Frontal midline (dmPFC) | 0, 54, 45 | -9.826 | 22.362, 17.053 | 0.041 | <0.001 |
| *Coffee −* | Posterior capsula interna rh | 15, -5, -3 | -8.69 | 17.741, 11.866 | 0.015 | <0.001 |
| *Alcohol −* | Outside brain (dlPFC) | 56, 4, 61 | -15.013 | 50.523, 45.025 | 0.055 | <0.001 |
| *Combined +* | Lateral ventricle rh (anterior horn) | 4, 4, 8 | 11.123 | 28.315, 22.988 | 0.078 | <0.001 |
| *Combined −* | Outside brain (dlPFC) | 53, 7, 61 | -15.458 | 53.476, 48.063 | 0.053 | <0.001 |
| *Smoking > Coffee* | Central sulcus of insula rh | 43, 1, -4 | 6.086 | 9.237, 4.123 | 0.026 | <0.001 |
| *Smoking > Alcohol* | Outside brain (supramarginal gyrus lh) | -50, -48, 68 | 7.55 | 13.662, 8.414 | 0.049 | <0.001 |
| *Coffee > Alcohol* | Outside brain (superior parietal lobule lh) | -35, -57, 77 | 10.161 | 23.829, 18.281 | 0.049 | <0.001 |
| *Coffee > Smoking* | Precuneus rh | 13, -75, 43 | 7.065 | 12.095, 6.992 | 0.063 | <0.001 |
| *Alcohol > Smoking* | Spinal cord lh | -4, -47, -72 | 8.259 | 16.134, 10.362 | 0.029 | <0.001 |
| *Alcohol > Coffee* | Spinal cord lh | -4, -48, -72 | 5.248 | 7.114, 1.964 | 0.018 | <0.05 |

**Peak statistics for experiment S1:** Smoking, Coffee and Alcohol

**RSN d25-2**

| ***Contrast*** | **Area** | **Peak (MNI)** | ***Z*** | **$-\log_{10}(p, q)$** | ***β*** | **p_hFDR** |
|---|---|---|---|---|---|---|
| *Smoking +* | Angular gyrus / superior parietal lobule rh | 41, -58, 51 | 4.82 | 6.143, 1.868 | 0.023 | <0.05 |
| *Coffee +* | Precuneus lh | -13, -74, 55 | 3.747 | 4.048, 0.000 | 0.02 | n.s. |
| *Alcohol +* | Middle temporal gyrus rh / temporal pole | 47, 16, -37 | 6.94 | 11.708, 6.748 | 0.033 | <0.001 |
| *Smoking −* | Angular gyrus rh | 60, -49, 24 | -4.627 | 5.731, 1.336 | 0.025 | n.s. |
| *Coffee −* | Lateral occipito-temporal sulcus rh (between fusiform and inferior temporal gyrus) | 53, -52, -21 | -5.02 | 6.589, 2.254 | 0.018 | <0.05 |
| *Alcohol −* | Parieto-occipital transition area lh | -22, -73, 51 | -3.564 | 3.739, 0.000 | 0.017 | n.s. |
| *Combined +* | Supramarginal gyrus lh | -59, -48, 42 | 5.489 | 7.693, 3.148 | 0.026 | <0.01 |
| *Combined −* | vmPFC rh | 11, 49, 44 | -4.656 | 5.793, 1.108 | 0.027 | n.s. |
| *Smoking > Coffee* | vlPFC rh | 58, 19, 11 | 5.124 | 6.825, 2.228 | 0.03 | <0.05 |
| *Smoking > Alcohol* | vlPFC lh | -49, 47, 5 | 4.062 | 4.614, 0.005 | 0.031 | n.s. |
| *Coffee > Alcohol* | vlPFC lh | -50, 45, -3 | 3.52 | 3.666, 0.000 | 0.027 | n.s. |
| *Coffee > Smoking* | Angular gyrus rh | 60, -50, 26 | 4.537 | 5.545, 1.226 | 0.037 | n.s. |
| *Alcohol > Smoking* | mPFC lh | -12, 63, 22 | 5.633 | 8.052, 3.417 | 0.065 | <0.001 |
| *Alcohol > Coffee* | Precuneus rh | 7, -63, 33 | 6.478 | 10.332, 5.893 | 0.057 | <0.001 |

**Peak statistics for experiment S1:** Smoking, Coffee and Alcohol

**RSN d25-6**

| ***Contrast*** | **Area** | **Peak (MNI)** | ***Z*** | **$-\log_{10}(p, q)$** | ***β*** | **p_hFDR** |
|---|---|---|---|---|---|---|
| *Smoking +* | Cerebellum rh (I-IV) | 18, -36, -24 | 4.72 | 5.928, 1.418 | 0.013 | <0.05 |
| *Coffee +* | Cerebellum rh (crus II) | 36, -78, -50 | 4.54 | 5.551, 0.664 | 0.016 | n.s. |
| *Alcohol +* | Lateral orbitofrontal cortex lh | -45, 44, -15 | 6.778 | 11.215, 6.329 | 0.038 | <0.001 |
| *Smoking −* | Precuneus lh | -9, -71, 31 | -4.777 | 6.052, 1.275 | 0.023 | n.s. |
| *Coffee −* | Angular gyrus lh | -50, -68, 26 | -5.59 | 7.945, 3.328 | 0.038 | <0.001 |
| *Alcohol −* | ACC lh | -5, 42, 4 | -6.709 | 11.008, 6.048 | 0.024 | <0.001 |
| *Combined +* | vlPFC rh | 60, 4, 7 | 5.131 | 6.841, 2.498 | 0.017 | <0.01 |
| *Combined −* | vmPFC / ACC lh | -6, 44, 10 | -5.144 | 6.871, 2.366 | 0.017 | <0.01 |
| *Smoking > Coffee* | Angular gyrus lh | -51, -68, 25 | 6.166 | 9.455, 4.974 | 0.061 | <0.001 |
| *Smoking > Alcohol* | vmPFC / ACC lh | -6, 43, 3 | 5.694 | 8.207, 3.517 | 0.032 | <0.001 |
| *Coffee > Alcohol* | Parieto-occipital transition area rh | 27, -88, 36 | 5.262 | 7.146, 2.337 | 0.033 | <0.01 |
| *Coffee > Smoking* | dlPFC rh | 56, 5, 47 | 4.512 | 5.494, 1.128 | 0.03 | n.s. |
| *Alcohol > Smoking* | Precuneus rh | 9, -70, 42 | 5.512 | 7.752, 3.151 | 0.045 | <0.001 |
| *Alcohol > Coffee* | Angular gyrus lh | -50, -68, 26 | 7.155 | 12.377, 7.756 | 0.072 | <0.001 |

**Peak statistics for experiment S1:** Smoking, Coffee and Alcohol

**RSN d25-9**

| *Contrast* | Area | Peak (MNI) | *Z* | −log$_{10}$(p, q) | *β* | p_hFDR |
|---|---|---|---|---|---|---|
| *Smoking* + | Precentral gyrus lh | -34, -20, 69 | 6.023 | 9.067, 4.492 | 0.028 | <0.001 |
| *Coffee* + | Angular gyrus rh | 58, -54, 42 | 4.624 | 5.726, 1.038 | 0.022 | n.s. |
| *Alcohol* + | SMA rh | 7, -5, 65 | 7.094 | 12.186, 7.333 | 0.037 | <0.001 |
| *Smoking* − | dlPFC rh | 35, 44, 40 | -4.625 | 5.728, 1.490 | 0.023 | <0.05 |
| *Coffee* − | Postcentral gyrus rh | 46, -28, 65 | -4.39 | 5.246, 1.366 | 0.021 | n.s. |
| *Alcohol* − | dmPFC lh | -17, 10, 70 | -4.777 | 6.052, 1.947 | 0.021 | <0.05 |
| *Combined* + | Superior temporal gyrus lh | -51, -17, 0 | 4.971 | 6.477, 2.367 | 0.016 | <0.01 |
| *Combined* − | Caudate nucleus rh | 12, -4, 18 | -5.28 | 7.190, 2.625 | 0.014 | <0.01 |
| *Smoking* > *Coffee* | Precentral gyrus lh | -40, -16, 64 | 5.583 | 7.928, 3.194 | 0.038 | <0.001 |
| *Smoking* > *Alcohol* | Precentral gyrus lh | -36, -20, 68 | 6.574 | 10.612, 6.130 | 0.048 | <0.001 |
| *Coffee* > *Alcohol* | SMA lh | -18, 7, 71 | 4.647 | 5.774, 1.174 | 0.03 | n.s. |
| *Coffee* > *Smoking* | Paracentral lobule lh | -6, -31, 66 | 4.78 | 6.057, 1.774 | 0.034 | <0.05 |
| *Alcohol* > *Smoking* | SMA rh | 7, -4, 66 | 6.192 | 9.527, 4.626 | 0.049 | <0.001 |
| *Alcohol* > *Coffee* | Precentral gyrus lh | 44, -14, 57 | 6.216 | 9.595, 5.093 | 0.038 | <0.001 |

**Peak statistics for experiment S1:** Smoking, Coffee and Alcohol

**RSN d25-11**

| ***Contrast*** | **Area** | **Peak (MNI)** | ***Z*** | **$-\log_{10}(p, q)$** | ***β*** | **p_hFDR** |
|---|---|---|---|---|---|---|
| *Smoking* + | Posterior cingulate cortex (PCC) lh | -3, -39, 32 | 4.635 | 5.748, 0.971 | 0.016 | n.s. |
| *Coffee* + | Putamen lh (surface) | -32, 0, 0 | 3.815 | 4.168, 0.000 | 0.01 | n.s. |
| *Alcohol* + | dlPFC lh | -47, 34, 24 | 8.644 | 17.566, 12.917 | 0.056 | <0.001 |
| *Smoking* − | Middle temporal gyrus lh | -57, -1, -29 | -5.967 | 8.919, 3.958 | 0.023 | <0.001 |
| *Coffee* − | Caudate nucleus lh | -8, 2, 12 | -5.001 | 6.545, 1.873 | 0.015 | <0.05 |
| *Alcohol* − | Parieto-occipital sulcus rh | 14, -77, 41 | -5.369 | 7.402, 2.634 | 0.025 | <0.01 |
| *Combined* + | Middle temporal gyrus lh (posterior part) | -58, -58, 7 | 5.873 | 8.670, 3.900 | 0.024 | <0.001 |
| *Combined* − | Middle temporal gyrus rh | 62, -9, -27 | -5.184 | 6.965, 2.564 | 0.019 | <0.01 |
| *Smoking > Coffee* | Occipital gyri rh | 50, -78, 16 | 4.519 | 5.508, 1.007 | 0.029 | n.s. |
| *Smoking > Alcohol* | Supramarginal gyrus lh | -62, -48, 32 | 4.938 | 6.404, 1.752 | 0.039 | <0.05 |
| *Coffee > Alcohol* | dmPFC lh | -13, 38, 55 | 4.841 | 6.189, 1.733 | 0.037 | <0.05 |
| *Coffee > Smoking* | Middle temporal gyrus lh | -57, -1, -29 | 4.48 | 5.428, 0.930 | 0.024 | n.s. |
| *Alcohol > Smoking* | dlPFC lh | -48, 34, 23 | 6.792 | 11.257, 6.808 | 0.066 | <0.001 |
| *Alcohol > Coffee* | dlPFC lh | -47, 33, 26 | 7.704 | 14.180, 9.477 | 0.072 | <0.001 |
