## Supplementary material for "PANDORA: Population Archive of Neuroimaging Data Organized for Rapid Analysis": Full Benchmarks

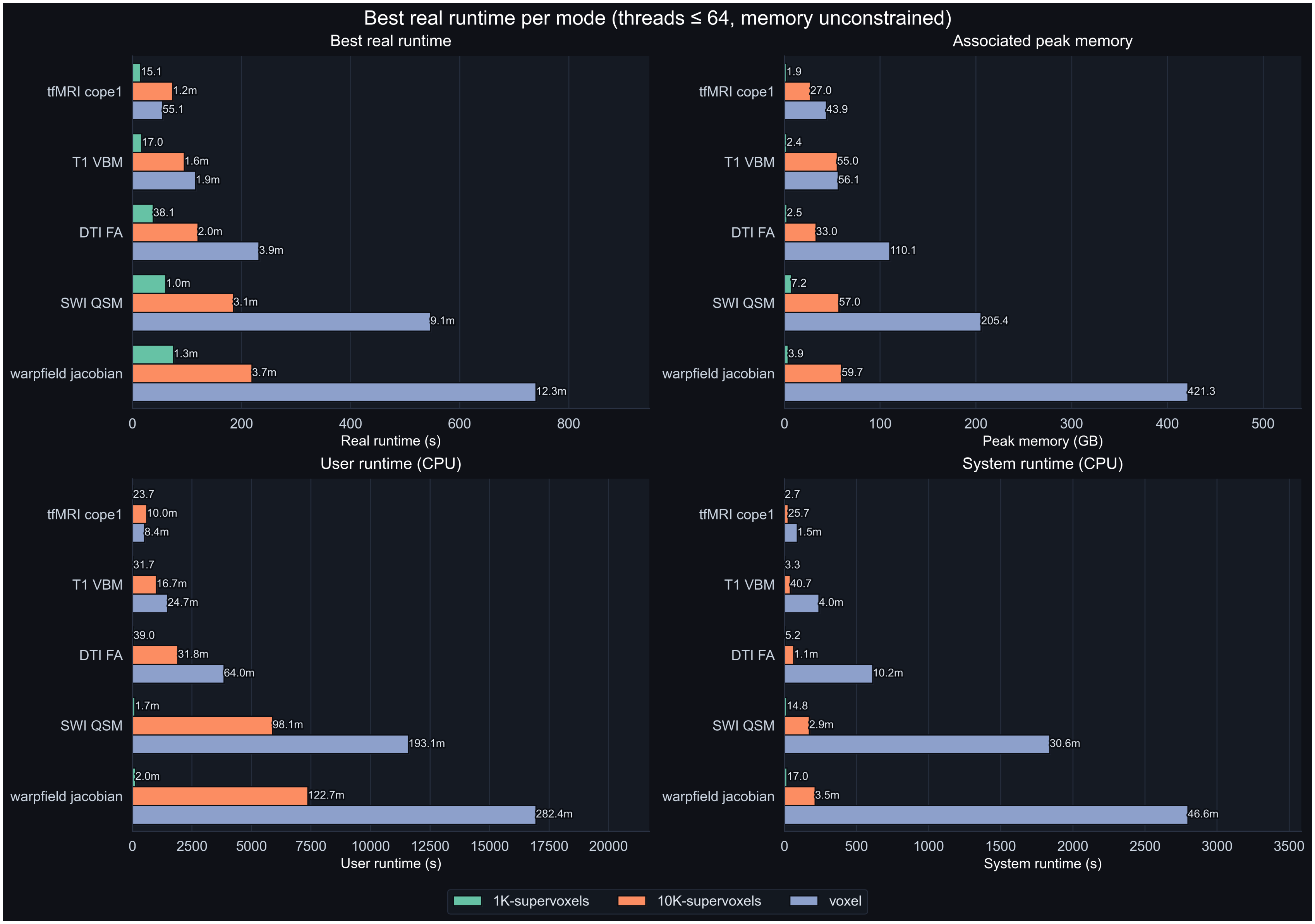


***Figure S7. fsl_glm benchmarks. Best real runtime per mode unrestricted.*** *1K-supervoxel analysis is clearly faster and more memory efficient than the other options. The advantage of 10K-supervoxels over full voxels becomes evident for sub-modalities with higher voxel counts. Lower panels show runtime portioned into single-threaded user- and system runtime. While compute is non-system dominated, system runtime reflects the massive overall reduction in I/O dependent processes.*

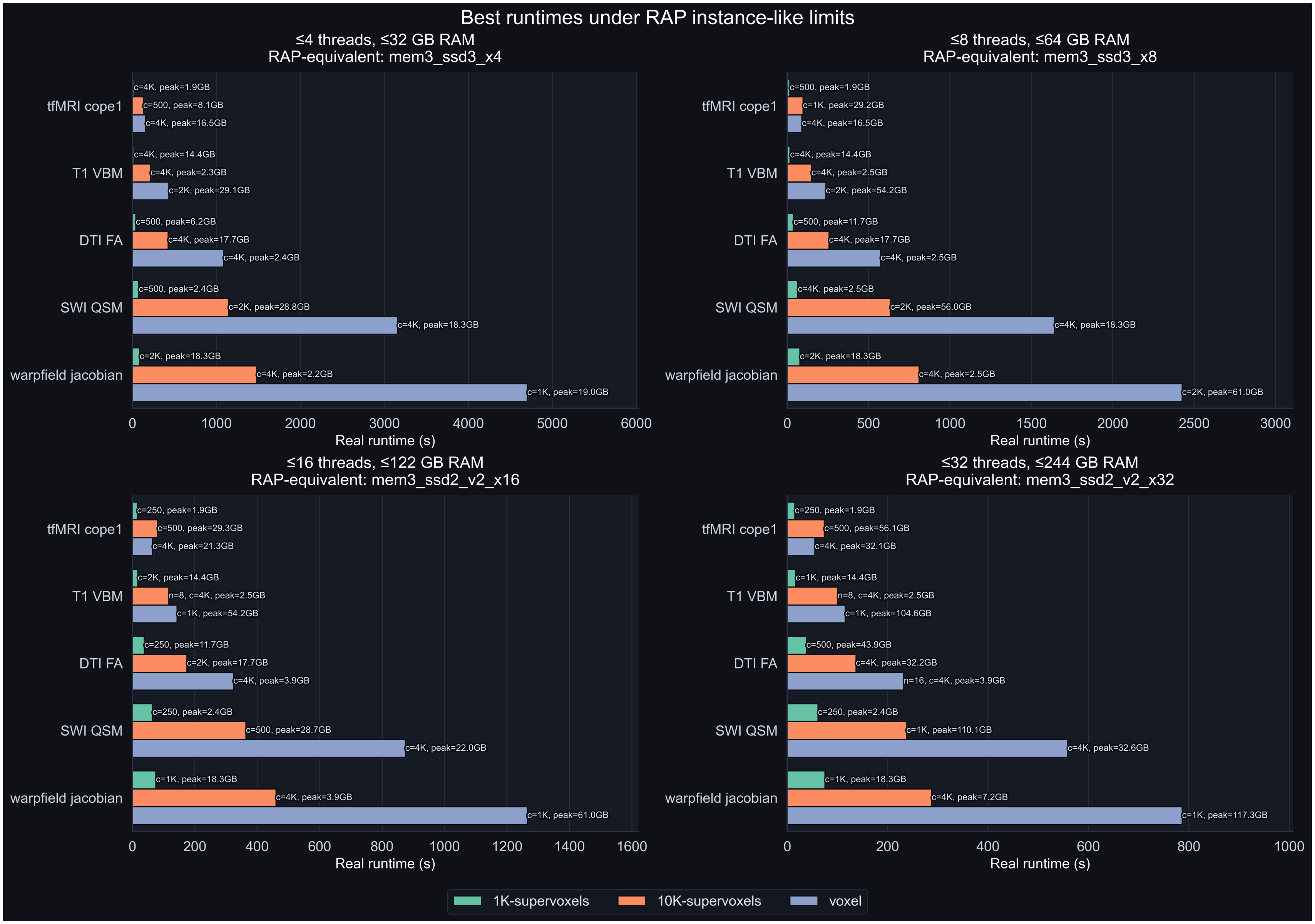


***Figure S8. fsl_glm benchmarks.*** *Best real runtime using UKB-RAP instance-like limits I.*

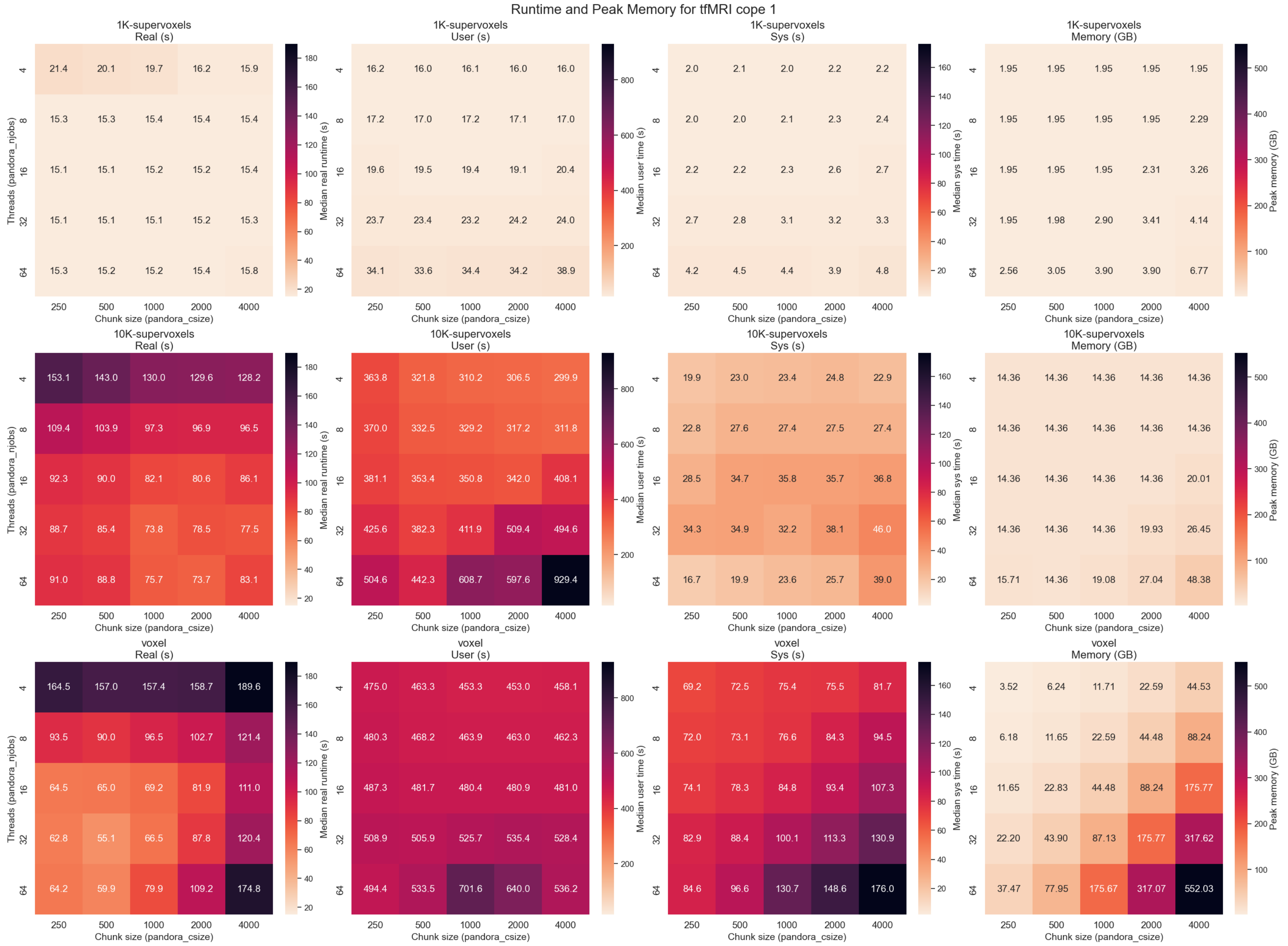


***Figure S9.1. fsl_glm benchmarks (tfMRI cope 1).*** *Real, system and user runtimes (columns 1-3) and associated peak memory consumption (column 4).*

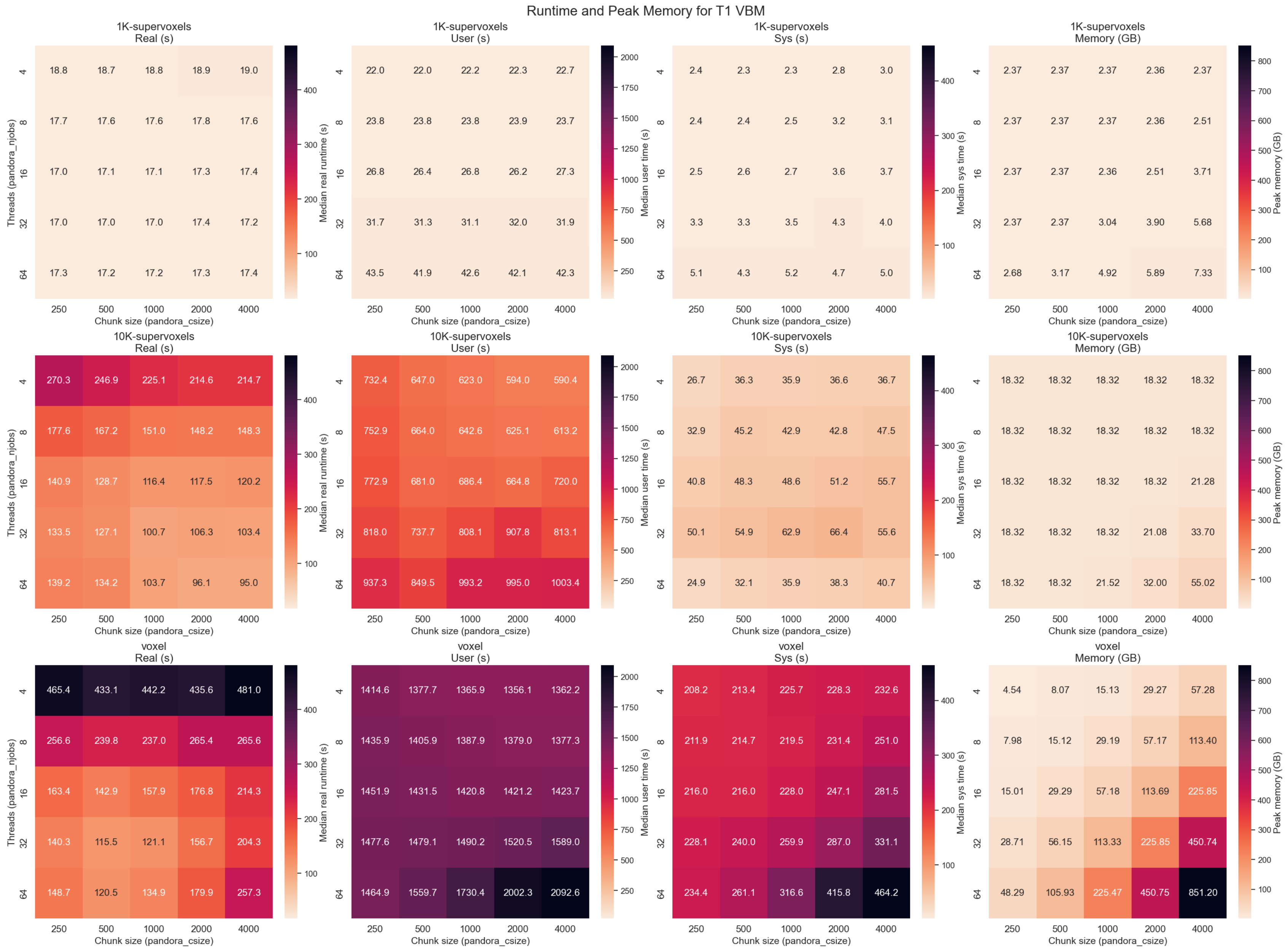


***Figure S9.2. fsl_glm benchmarks (T1 VBM).*** *Real, system and user runtimes (columns 1-3) and associated peak memory consumption (column 4).*

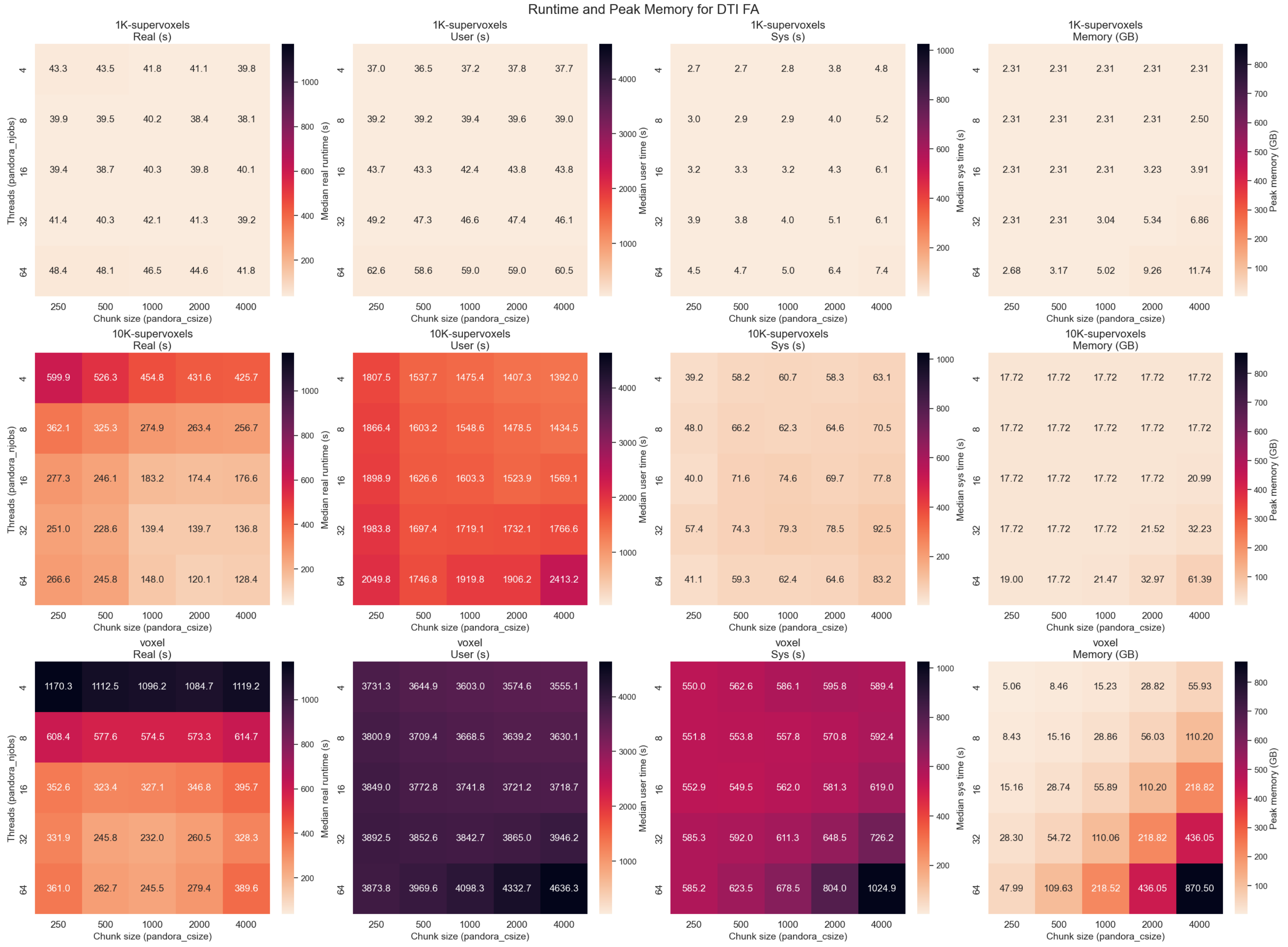


***Figure S9.3. fsl_glm benchmarks (DTI FA).*** *Real, system and user runtimes (columns 1-3) and associated peak memory consumption (column 4).*

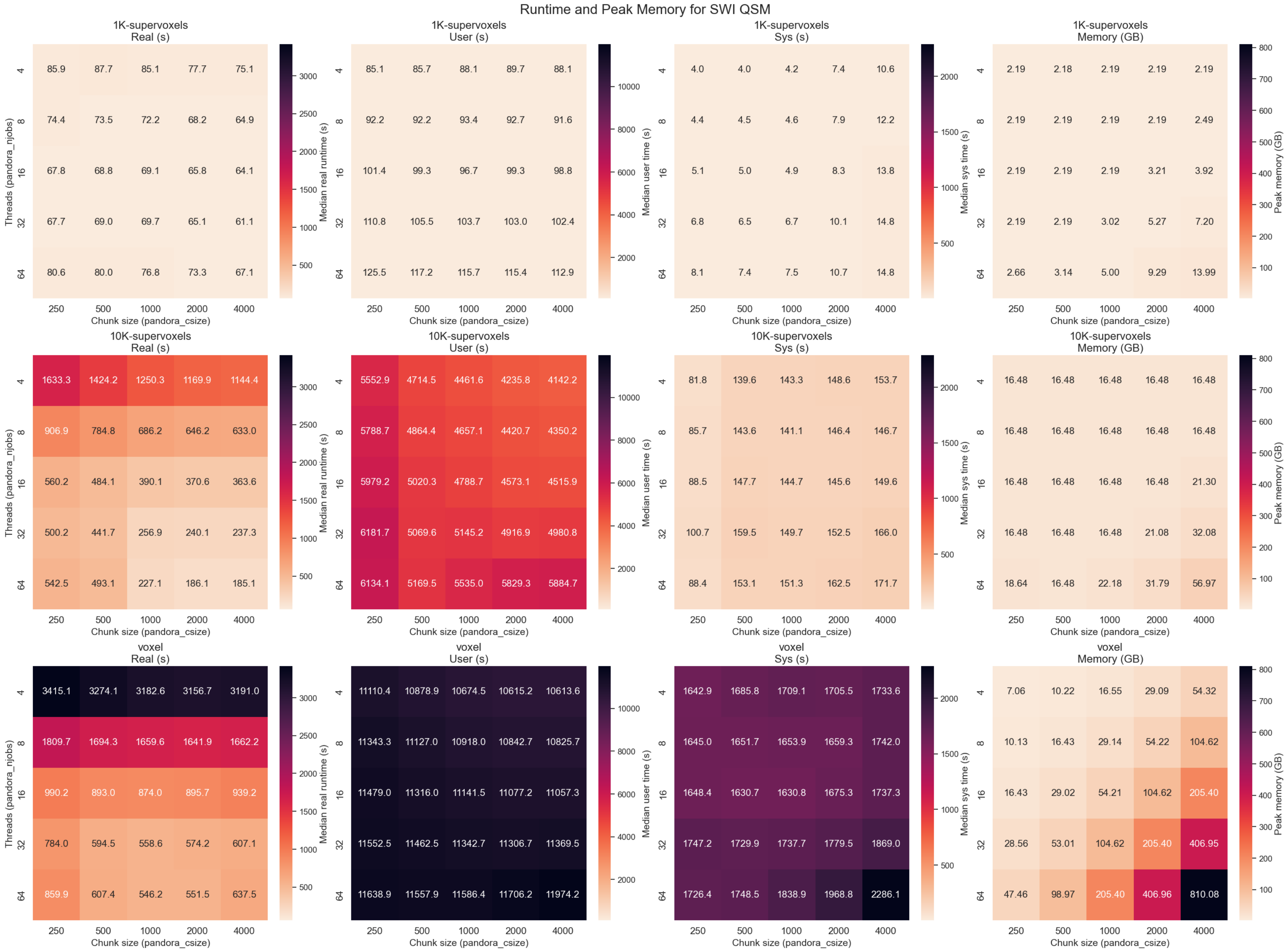


***Figure S9.4. fsl_glm benchmarks (SWI QSM).*** *Real, system and user runtimes (columns 1-3) and associated peak memory consumption (column 4).*

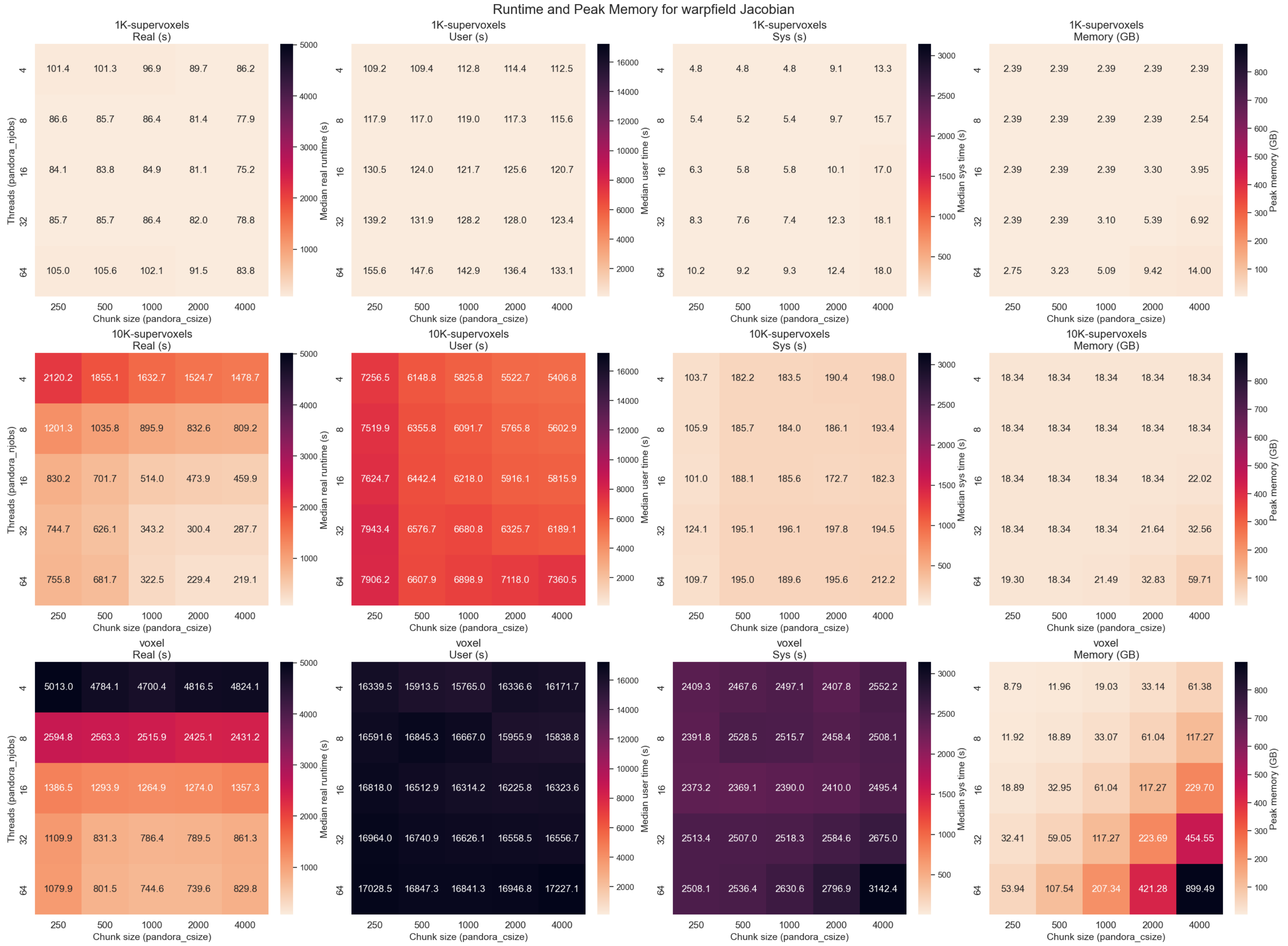


***Figure S9.5. fsl_glm benchmarks (warpfield Jacobian).*** *Real, system and user runtimes (columns 1-3) and associated peak memory consumption (column 4).*
